# In-silico, interactomic and clinical validation based approach for screening and identification of miR biomarkers involved in Oral submucous fibrosis to Oral squamous cell carcinoma transition

**DOI:** 10.1101/2020.10.05.20206904

**Authors:** Shweta Ukey, Jeewan Ram Vishnoi, Chinmayee Choudhury, Purvi Purohit, Shailendra Dwivedi, Ankit jain, Ankita Chugh, Poonam Elhence, Puneet Pareek, Sanjeev Misra, Praveen Sharma

**Affiliations:** All India Institute of Medical Sciences, Jodhpur, Rajasthan, India; Department of Experimental Medicine and Biotechnology, Post Graduate Institute of Medical Education and Research, Chandigarh., India; All India Institute of Medical Sciences, Gorakhpur, Uttar Pradesh, India

**Author notes:** Corresponding Author Dr. Praveen Sharma, Professor and Head, Department of Biochemistry, All India Institute of Medical Sciences, Jodhpur, Rajasthan, India, 342005. SU, JV, AC, AJ, PP, PE, PP and PS thank AIIMS, Jodhpur for financial support and research facilities, SD thank AIIMS, Gorakhpur for financial support, CC thank DST-INSPIRE research grant and Department of Experimental Medicine and Biotechnology, Post Graduate Institute of Medical Education and Research, Chandigarh. SU thank CSIR for financial support in form of CSIR-Senior Research fellowship.

**Keywords:** Oral Submucous Fibrosis, Oral Squamous Cell Carcinoma, microRNA’s, Interatomics, Risk Stratification Biomarkers

## Abstract

Oral Squamous Cell Carcinoma (OSCC) is common preventable disease when diagnosed early, but mostly its progression follows transition from oral potentially malignant disorders (OPMDs) like Oral Submucous Fibrosis (OSF). However, it is difficult to predict possibilities of progression in these premalignant lesions hence, identification of molecular biomarkers would have major clinical impact in early diagnosis and better prognosis. In this context microRNA’s(miR’s) provide better opportunities in malignancy prediction and demarcation in OSF to OSCC transition as they perform key regulatory roles in many tumorigenic processes. Here, we computationally screened differentially expressed miR’s of OSCC and OSF from public databases followed by construction of protein interaction networks and enrichment analyses. The relevant miR’s were validated using qPCR of total 93 samplesincluding 34 OSCC, 30 OSF and 29 control blood and tissue samples. We identified significant down regulation of miR-133a-3p in OSCC compared to controls and interesting up-regulation compared to OSCC and control. miR-9-5p was up-regulated in OSF as well as OSCC and down-regulated in OSF compared to OSCC. Therefore, these two miR’s may serve as risk stratification biomarkers with validation in larger categorical datasets.

## 1. INTRODUCTION

OSCC is the commonest type of Head and neck cancer(HNSCC). OSCC has a low five-year survival rate because of late diagnosis and frequent recurrence[1]. Its progression is mostly seen from a group of precursor lesions called OPMDs[2]. Many retrospectives and epidemiological studies on OSCC suggests that OPMDs such as OSF, Oral lichen planus (OLP), oral leukoplakia and human papillomavirus (HPV)have a significant contribution to OSCC development [3,4]. OSF has a high malignant transformation rate which is up to 15% and which differs with ethnicity, region, habits and culture [5,6]. Chronic inflammation of oral mucosa, excessive collagen deposition in the connective tissue, local inflammation of lamina propia are the major pathological characteristics of OSF[7]. OSF being a precursor to oral cancer comprises of various epithelial alterations which vary from atrophy with hypoplasia to hyperplasia and dysplasia[8]. This shift in epithelial compliance leads to carcinomatous processes such as epithelial-mesenchymal transition (EMT) [9,10]. In addition to EMT other pathological symptoms for malignant transformation of OSF are inflammatory cytokines and growth factor, cell cycle regulation, hypoxia and angiogenesis[11]. All these mechanisms participating in multistep carcinogenesis and progression are regulated genetically and epigenetically.

Protein coding genes and their contribution in carcinogenesis are always being explored but noncoding genome especiallymiR’s and its involvement in the initiation, progression, metastasis, chemoresistance and recurrence remain relatively unexplored so far[12]. miR’s are endogenous, 18-24 nucleotide non-coding RNAs, they post-transcriptionally regulate mRNA expression through binding to semi-complimentary sequences on 3’ untranslated regions (3’UTR) of target mRNA resulting translational inhibition or mRNA degradation[13].

In past decade studies have established miR’s as key regulators of the oncogenic potential of cells, and also they are predicted to regulate the expression of atleast 60% of human genes[13,14]; therefore the aberrant expression of miR’s in OSF can’t be ignored in malignant transformation of OSCC.

Many studies are done to explore the role of miR’s in OSCC tumors, especially in invasion, migration angiogenesis and their related pathways[15–18]. Scapoli *et al*. have investigated miR’s role in metastasis of OSCC[19], miR profiling is also done in OSCC tissue and blood for investigating its correlation with clinico-pathological profile of tumors and prognosis to investigate if miR’s could serve the purpose of biomarkers[2,17,18,20]

There are few studies with analyses of change in miR expression associated with OSF pathogenesis[21–23].A recent systematic review by El-Sakka*et al*. have extensively explored miR studies, and their correlation with OPMDs, their findings inthe majority of included studies have nospecific relation between differentially expressed miR’s in OSCC with OPMDs[2]. Some studies have shown the correlation between the expression of miR’s with OPMDs such as a study by Brito *et al*.done with both formalin-fixed tissue and blood showed significant upregulation of miR-21, miR181b and miR345in OSCC compared to oral leukoplakia[24]. Similarly in study by Harrandah *et al*. of formalin-fixed, paraffin-embedded samples from progressive premalignant lesions and OSCC tumorsobtained from the same site of tumor they found miR-21 up-regulated, miR-494 downregulated, and miR-375 downregulated, in OSCC tissue compared to premalignant lesions[21]. Heravi *et al* showed downregulation of miR-29a (p<0.05), miR-146a (p<0.05), miR-223 and up-regulation of miR-27b (p<0.05)in OLP compared to OSCC[25]. Yi Lu*et al*. investigated the functional role of miR-200 in buccal mucosal fibroblasts and concluded its association with ZEB1 as target and involvement with areca nut associated OSF pathogenesis[26]. Thus there are a limited number of reports which talk about the transition of OSF to OSCC. Therefore, identification of early-stage molecular signatures that predicts tumorigenesis is need of the hour.

With the development of multiple disease-specific, validated and predicted datasets, the in-silico analysis of various target genes, regulatory micro RNAs and their interactomes has become easier[27]. Data available in such public datasets can be used to screen, compare and analyze information, which otherwise would be very difficult to collect from individual studies. Here we used data from publically available datasets of miR’s specific for OSCC/ HNSCC and OSF and cancer-specific datasets so as to perform in-silico analysis of common miR’s and their interactome. Further, Protein-protein interaction (PPI) of these genes were performed using STRING and scrutiny of the obtained networks were done, and the miR’s targets associated to pathways related to OSF and OSCC pathogenesis were selected, and their expression was validated using Real-time PCR in patient cohort of OSCC and OSF. A systematic work flow is shown in **Figure 1**.

**Figure 1.**
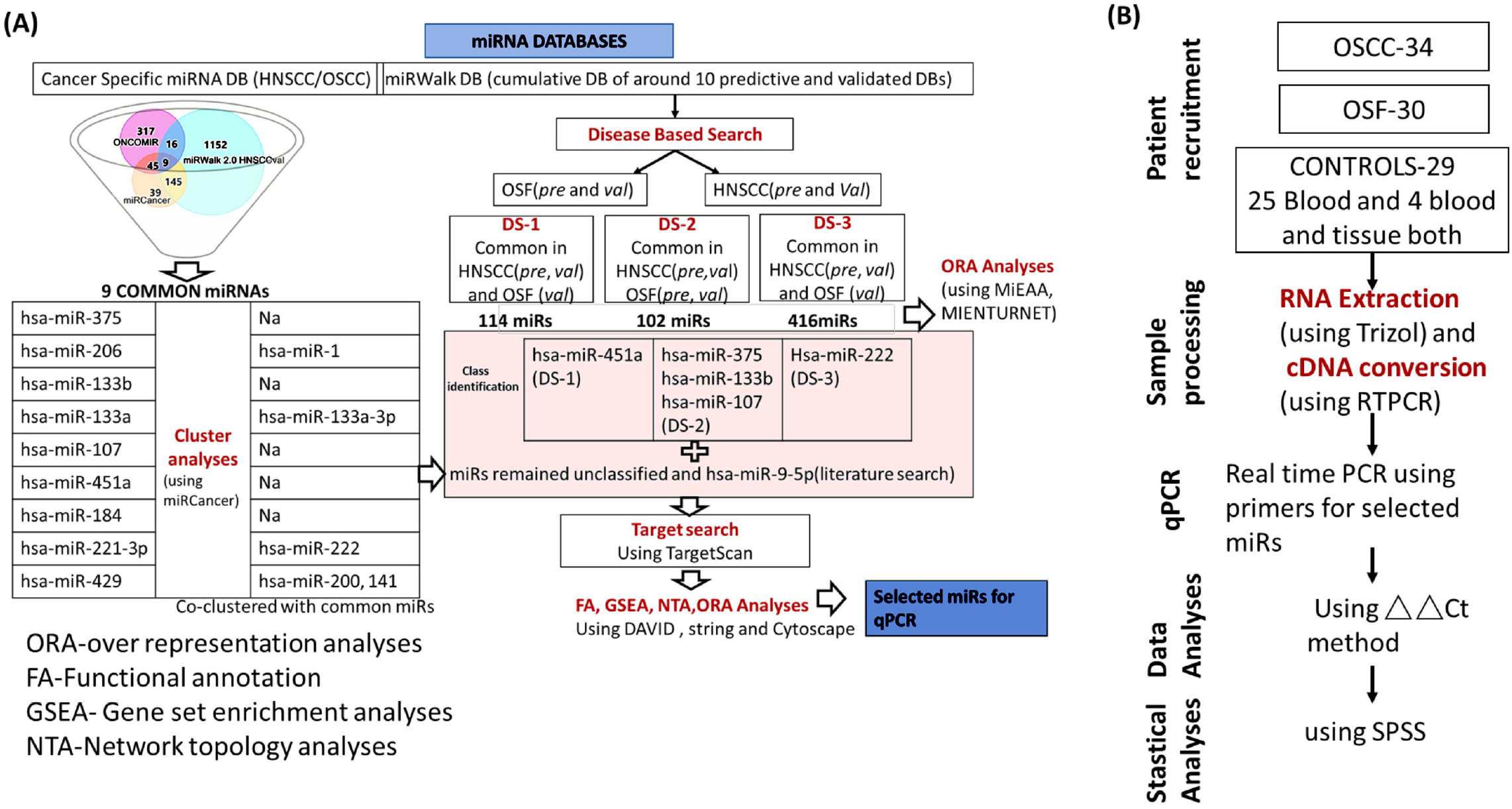
Workflow for (A) Database search and bioinformatics and interatomic analyses for miR’s (B) Patient recruitment and qPCR

Current study therefore provides for the first time both in-silico analysis of OSF / OSCC regulating miR’s as well as the validation of the target miR’s in clinical samples.

## 2. MATERIAL AND METHODS

### 2.1 Database Search

The database search was performed in two steps, in first step we collected miR’s data from miRWalk database[28], which is a cumulative common database of more than 10 (TargetScan, miRDB, miRBase, miRTarBase, TarPmiR PicTar, miRanda etc.) predictive and validated miR Databases. Both predictive and validated data modules were exploited for disease associated miR’s for OSF and HNSCC as OSCC was not available in the disease list of most of the databases and according to American Cancer Society OSCC is the most common and prevalent HNSCC, with tumor sites of tongue, tonsils, gums, floor of mouth, oropharynx with left out sites of Lyrnx, lymphoid tissue and nasopharynx in HNSCC. Secondly, a cancer specific Database search was performedto obtain most common and differentially expressed miR’s in HNSCC/OSCC from renowned experimentally validated datasets ONCOMIR[29], miRCancer[30] and validated miR’s for HNSCC from miRWalk. We sorted commonly found validated miR’s which are differentially expressed with more than 1.5 fold change in cancer specific databases for HNSCC/OSCC further, miR enrichment analyses was done using miEEA[31], the miR’s of relevance were further subjected to Targetscan database[32] to predict miR target genes.

### 2.2 Cluster analyses

Cluster analyses was done using miRCancer sequence analyses platform selecting batch of oral cancer, HNSCC, Oral squamous cell carcinoma and oropharangeal carcinoma in cancer type, keeping default parameters with intra distance at 6 points with a 1 point gap and substitution.

### 2.3 Over Representation analyses

We performed over representation analyses for the three datasets obtained to explore whether these show any significant representation pattern associated to OSF pathogenesis or any set of miR’s within the list are showing involvement in any carcinomatous pathways using miEEA[31]and MIENTURNET[33] which are online miR enrichment tools.

### 2.4 Protein-Protein interaction networks and molecular function enrichment

i. Total 16 miR’s wereobtained in common from cancer specific databasesalong with their mature iso-strands and their co-clustered miR’s and miR-9 was selectedbased on literature search. Targets of each miR’s were searched using TargetScan and cutoff set according to weighted context ++ scores[32]to - 0.5 in order to minimize false positive association of miR’s with their targets and to obtain manageable number of targets.
ii. Obtained targets for each selected miR were subjected to STRING[34], we established networks using target genes for each miR’s taking confidence score cutoff greater than 0.7 which is considered as high confidence in string and the molecular functions were scrutinized and interpreted using Cytoscape[35].
iii. DAVID functional annotation tool[36] was used to further explore the enrichment of the target genes of miR’s found to be related to pathologies associated to OSF and OSCC

### 2.5 Clinical Samples

Samples used in this study were tissue and blood samples of OSCC and OSF patients collected from Department of Onco-surgery and Department of Dentistry, All India institute of medical sciences (Jodhpur, India) with informed patient consent from July 2018 to February 2020. Control blood samples were collected from healthy volunteers consenting to participate in the study and control tissue samples were collected from patients undergoing dental procedures in Department of Dentistry with informed consent.

Patient’s biochemical parameters such as LFT, KFT, Blood sugar, Hemoglobin were collected from pre-anesthetic check-up done immediate before surgeryalong with demographic characters such as age, sex and volunteer’s biochemical parameters were asked and noted from their recent reports. Patient and control smoking, drinking, smoking and chewing habits were noted depending upon their habit frequency and time. The patients with HPV infection, were excluded from the study from all the three groups. All the samples were histologically verified in Department of Pathology, AIIMS Jodhpur. This study is approved by Institutional Review Board of AIIMS, Jodhpur.

### 2.6 RNA isolation

Total RNA was isolated from all OSCC, OSF and Control tissue and blood samples using TRIzol reagent (Himedia) according to the manufacturer’s instructions. Quality and quantity of total RNA was analyzed spectrometry and samples with 260/280 ratio of 1.9±0.5 were taken for further analyses.

### 2.7 Quantitative real-time PCR(qPCR)

SYBR-based qPCR was used to quantify mature miR expression using the miSript PCR system by QIAGEN. This three component system includes a reverse transcription kit with an optimized blend of poly(A) polymerase and reverse transcriptase for cDNA synthesis and a SYBR-Green PCR kit. miR’s specific primers were custom designed. The qPCR reaction mix was made according to the manufactures instructions and the conditions followed were: Initial activation for 15 min at 95^0^Cfollowed by 40 cycles of 94^0^C for 15sec, 55^0^C for 30 sec and 70^0^C for 30 sec using a Bio-Rad CFX96^DX^ Real-Time quantitative PCR machine. The relative miR expression level was normalized to that of U6 using 2^-ΔΔCt^ cycle threshold method[37].

### 2.8 Statistical Analyses

In the internal test cohort, the univariate analysis was performed for all covariates including age, gender, TNM stage, histologic grade. Normality tests such as Kolmogorov-Smirnov and Shapiro-Wilk were performed to check the distribution of log2FC values for each miRexpression data to confirm data as parametric or non-parametric and accordingly for the use of tests forsignificance, p value less than 0.05 was considered as not normally distributed data and non-significant p values in normality test are considered as normally distributed data. miR datasets for three groups i.e. OSCC, OSF, Control were normally distributed datasets analyzed using one way analyses of variance(ANOVA) and Post Hoc tests like Bonferroni and Tukeys were also performed, similarly analyses between two individual groups with parametric data were performed using students T-test. Construction of receiver operator curve (ROC) was done using SPSS for analyzing sensitivity and specificity. Statistical analyses were performed using SPSS 25.0 (SPSS Inc., Chicago, IL, USA).

### 3. RESULTS

### 3.1 Data mining analyses

miRWalk predicted and validated datasets Venn diagrams for HNSCC/OSCC and OSF are shown in Figure 2(A) and 2(B), these show that around 80 % of HNSCC miR’s which are predicted to be differentially expressed whereas only 17% of OSF predicted are validated so we expect there is a large scope to explore OSF miR’s in tumorogenesis. Four sets of miR lists HNSCCvalidated(*val*), HNSCCpredicted(*pre*), OSF(*val*) and OSF(*pre*) were compared and three datasets were made with common miR’s named as **DS-1**(HNSCC*val*, HNSCC*pre*, OSF*pre*), **DS-2**(HNSCC*val*, HNSCC*pre*, OSF*val* and OSF*pre*) and **DS-3**(HNSCC*val*, HNSCC*pre*, OSF*val*). There were 114, 102 and 416 miR’s obtained common in DS-1, DS-2 and DS-3 respectively(**supplementary Table 1**)and this list of miRs was subjected to miR enrichment analyses.

**Figure 2.**
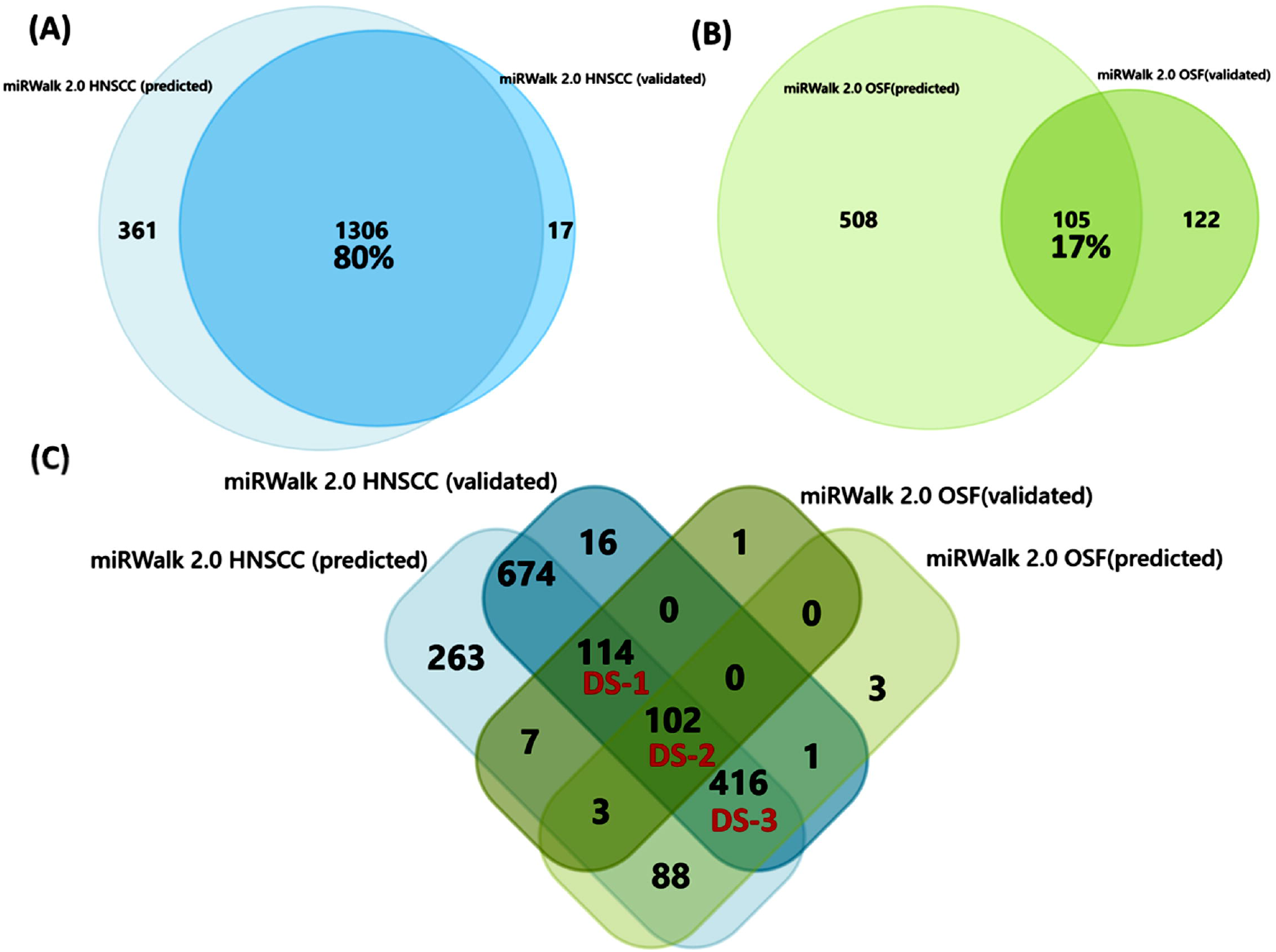
miRWalk database search for disease associated miR’s in HNSCC and OSF validated and predicted target module.(A) and (B) are Venn diagram showing predicted and validated miR’s in HNSCC and OSF respectively. (C) is venn diagram showing coinciding miR’s between the four groups.DS-1 are common miR’s between (HNSCC*val*, HNSCC*pre* and OSF*val*), DS-2 are (HNSCC*val*, HNSCC*pre*, OSF*val* and OSF*pre*) and DS-3 are (HNSCC*val*, HNSCC*pre* and OSF*pre*)

Cancer specific database search(**supplementary table 2**) providednine common differentially expressed miRs in HNSCC/OSCC. The clustering analyses (**supplementary Table 3**) providednine common miRs that were co-clustered with additional six miRs. We obtained 15 miRs and checked if they are present in any of DS’s. hsa-miR-451a(DS-1) hsa-miR-375, hsa-miR-133b, hsa-miR-107 (DS-2) and hsa-miR-222 (DS-3) were found. All these 15 miRs along with miR-9 both strands were subjected to target search using TargetScan followed by their PPI analyses to predict if these miR targets were associated with OSF pathogenesis.

### 3.2 miRNA enrichment and annotation analyses

miR’s listed as DS-1, DS-2, DS-3 were analyzed separately using two different miR enrichment tools miEAA and MIENTURNET to explore if the list of miRs in each dataset is enriched or associated with any particular pathways involved in the pathogenesis of OSF to OSCC. Our analyses results do not provide any crude correlation of OSF pathogenesis with the miR’s listed in each DS’s as there were many miR’s and each DS have many targets involved in multiple pathways. While performing the Enrichment and Over Representation Analyses from the three datasets many of the miR’s were not even identified by the enrichment tools, the reason may be most of these miRs are predicted in the databases according to their complementarity with the genes involved in various diseases using different algorithmsand if their differential expression is validated experimentally also, but their associated targets are still unexplored. The list of, miRs unidentified are given in **supplementary Table 1**

**Table 1.**
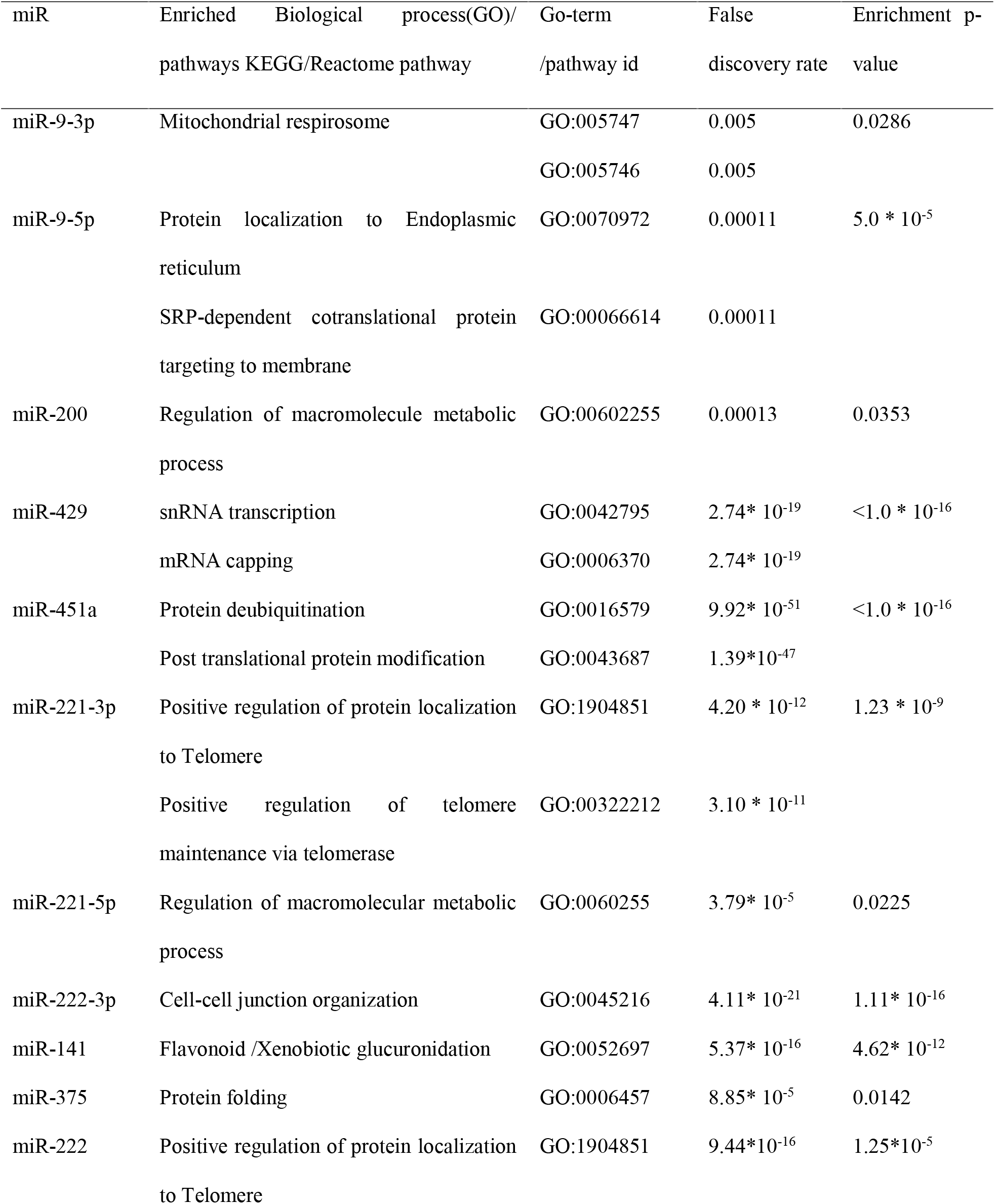
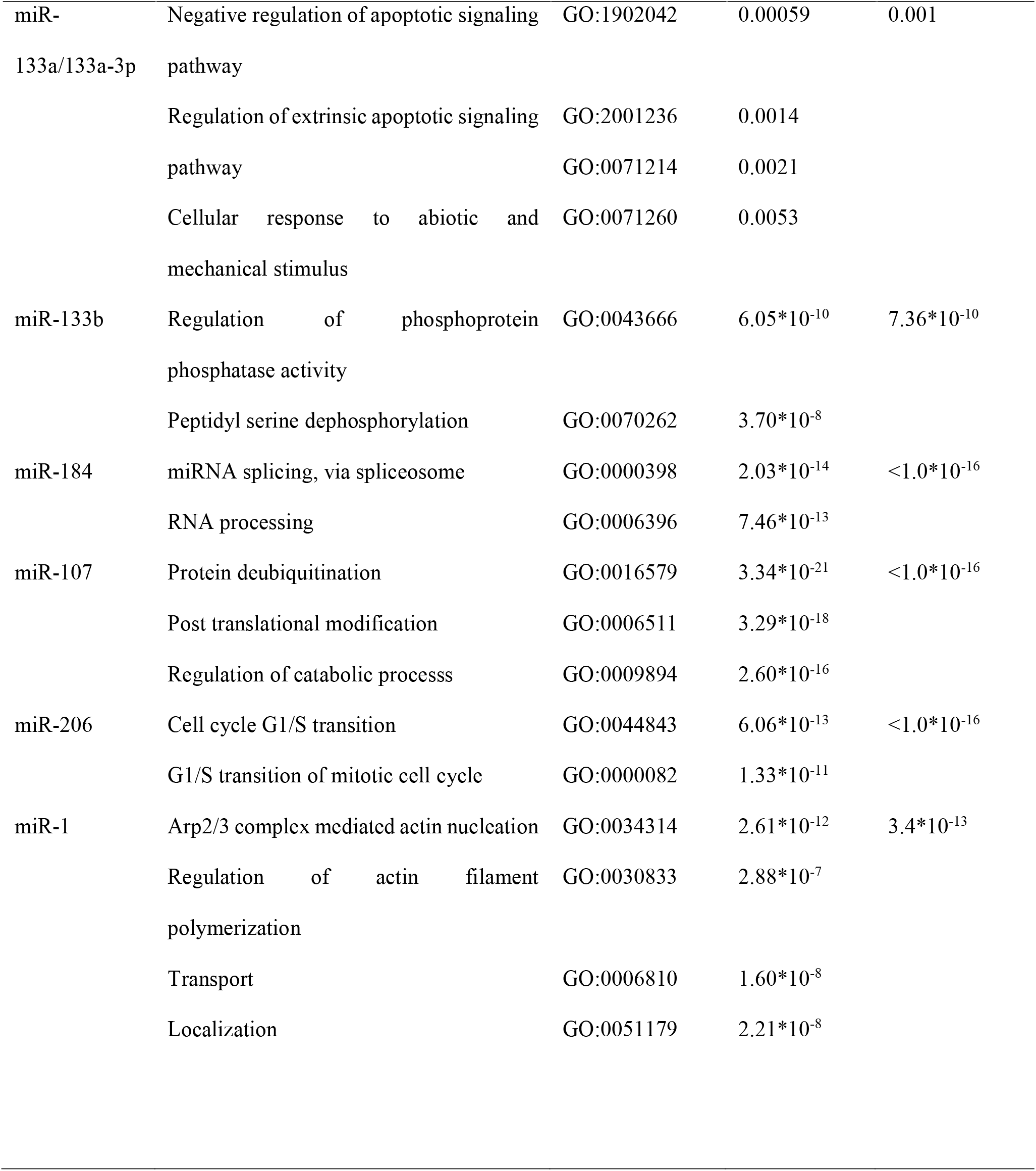
STRING statistical analyses of enriched target genes for each miR listed.

The ORA analyses of DS-3 miR listin Disease(MNDR Database) category showed high under-representation associated with many cancer types including squamous cell carcinoma and OSCC with a highly significant p and adjusted p values shown in **supplementary Table 4** Underrepresented or depleted miR’s involved in signaling may be equally important as over represented ones if they are strongly significant because if these show over representation and differential expression it affects the normal functioning associated with these cancer types. Probably some of these under represented miRs have significant involvement in cancer associated processes if over represented but, still we cannot predict any relevance to OSCC transition from OSF. Therefore, for further studies we took the common miR’s which were differentially expressed in cancer specific databases.

### 3.3 STRING network and molecular function analyses

The TargetScan predicted miR targets with total context++ score less than 0.5 are mentioned in **supplementary Table 4** These genes were subjected to PPI using STRING, (STRING enrichment statistics are mentioned in **Table1)** which mentioned most enriched (pink) and successive enriched (cyan) targets of each miR (shown in **Figure 3)** and **supplement figure 1** with their gene ontology term, their enrichment score and false discovery rate. miR-133a/miR-133a-3p target proteins were enriched in negative regulation of apoptotic signaling pathway, regulation of extrinsic apoptotic signaling and cellular response to abiotic stress and mechanical stimulus, miR-133-b enriched in phosphoprotein phosphatase activity and peptidyl serine dephosphorylation which is associated to apoptotic signaling. There are studies which show down regulation of miR-133a [38–40]andmiR-133b [38]but no significant expression profiling with OPMDs. miR-107, miR-451a targets were enriched in protein deubiquitination and post translational modifications, miR-184 in miR splicing via spliceosome and RNA processing, that are the most common type of regulations in cancers. There are studies which mentioned miR-451a as pro-apoptotic which inhibits aggressiveness features in squamous lung cancer[41], miR-107 regulates proliferation and migration in OSCC[42], miR-184 associated to proliferation and Cisplatin resistance in OSCC[43] miR-221-3p/miR-222 belongs to same class and were enriched in positive regulation of protein localization to telomere and telomere maintenance via telomerase which is very important pathway for cancer pathogenesis. Various studies have shown that overexpression of miR-221-3p/222-3p increases growth and tumorigenesis in OSCC [44] but their significance to OPMDs are not mentioned to the best of our knowledge.miR-9 was selected in this study due to its controversial expression profile in many cancer types such as down regulated in Breast cancer, Renal cell carcinoma, gastric cell carcinoma and HNSCC) on the other hand is found to be up-regulated in hepatocellular, gliomas and colorectal cancers[45]. In a study by Kim *et al*, compression induced miR-9 downregulation and reversal was seen in breast cancer tissues [46], this suggest that mechano-transduction is followed in miR-9 which may be associated to OSF also, therefore we decided to explore miR-9 in OSCC and OSF tissues. In our enrichment analyses miR-9-5p targets are enriched for protein localization to endoplasmic reticulum and SRP-dependent co-translational protein targeting to membrane and miR-9-3p is enriched in mitochondrial respirosome. Protein targeting plays key role in cancer signaling such as evading growth repressors, angiogenesis, and metastasis signaling therefore contribution of miR-9-5p contribution can’t be ignored. Also, there is a recent study which found miR-9 enriched in HPV infected HNSCC patients and downregulates PPARD to induce macrophage polarization[47]. miR-1 is enriched in ARP2/3 complex mediated actin nucleation and regulation of actin filament polymerization also in transport and localization. miR-200, 221-5p targets were enriched in macromolecule metabolic process which is abroad area for investigation and miR-200/ZEB1 axis is already proven for epithelial mesenchymal transition in OSCC and OSF[48], hence already included as biomarker in OSF tissue.miR-429 found to be enriched in snRNA transcription, mRNA capping also it is co-clustered with miR200 and miR-141 and called as miR-200 family and reported to target ZEB1 in OSCC[49]. miR206 targets are found to be enriched for cell cycle G1/S phase transition with a high PPI enrichment p value (<1.0*10^−16^) it is having genes associated to cyclin dependent protein holoenzyme complex, transferase complex and catalytic complexes which are surely dysregulated in cancers. Two out of four studies showed downregulation of miR-206 in OSCC compared to normal one study showed upregulation and one did not specified the correlation but when compared OPMDs to normal tissue none of them showed any significant difference [2] therefore we did not selected it for further validation by qPCR. On the basis of above analyses we selected miR-9-5p, miR-221-3p and miR-133a-3p to explore them further using DAVID 6.8 which is a functional annotation (FA) tool followed by qPCR on clinical samples.

**Figure 3.**
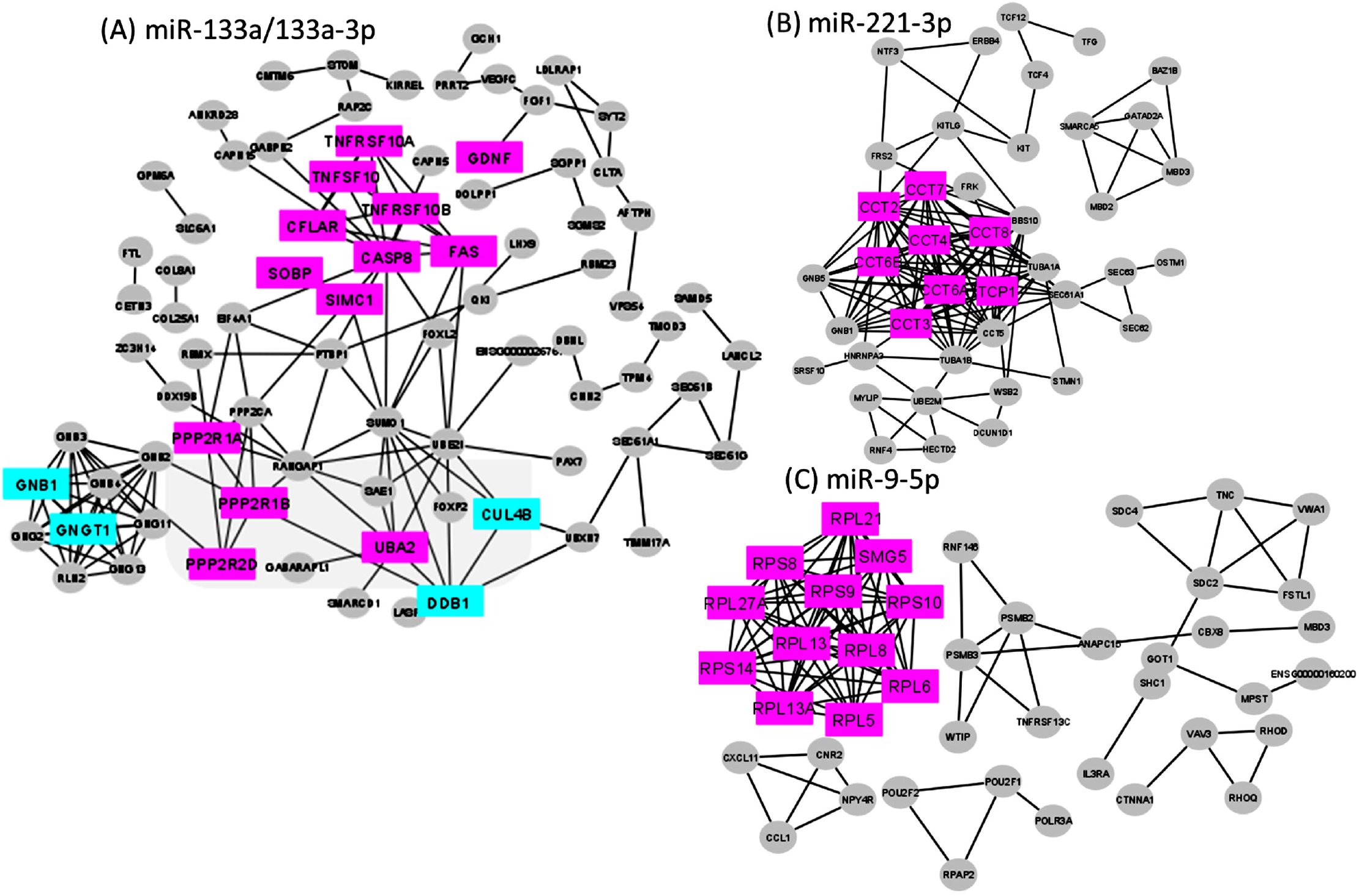
STRING networks showing most enriched (pink) and successive enriched gene(cyan) functions using STRING and CYTOSCAPE for three selected miRs.

### 3.4 Functional Annotation analyses

The FA results are summarized in **Figure 4** those are self-exploratory and suggests that there may be some association of our selected miRs in OSF pathogenesis due to their significant involvement in apoptosis, cell signaling, cell to cell adhesion, and their presence in thecell components such as intracellular membrane, plasma membrane and cell–cell adherens. Considering these results, we further proceeded with the qPCR of these miR’s with clinical samples

**Figure 4.**
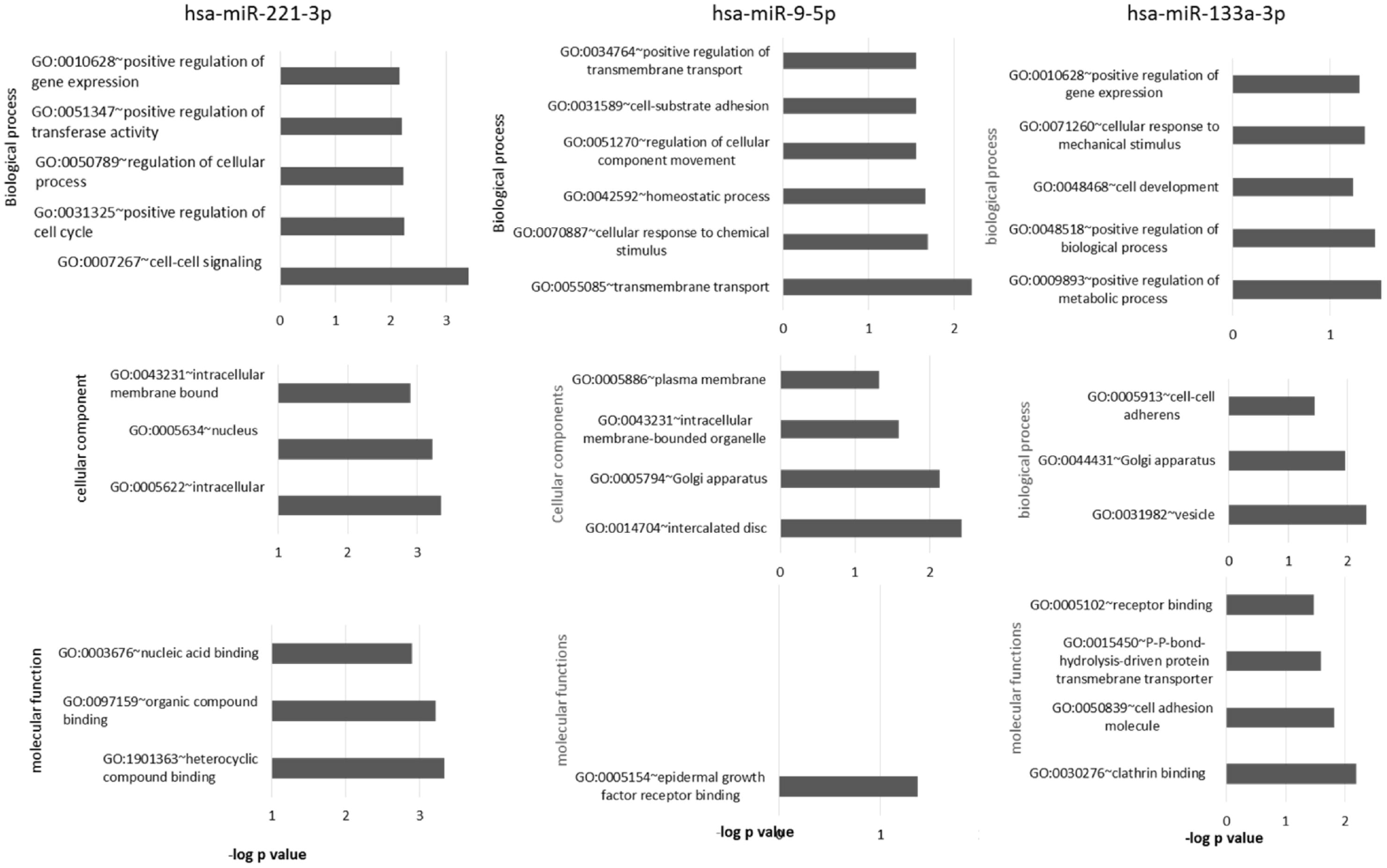
Function annotations of the three miR’s selected after STRING PPI and detailed biological processes, cellular component and molecular function available from DAVID 6.8 functional annotation tool.

### 3.5 miR expression analyses using qPCR

The patient cohort contained 34 consecutively collected tissue and blood samples of OSCC, 30 OSF and 29 control samples the patient and control clinicopathological parameters are mentioned in **Table 2A**. The differential expression of the miRs compared to controls in OSF and OSCC groups are mentioned in **Table 2B**. As our data was normally distributed statistical tests performed for significance wasone way ANOVA. The log2FC values between two groups are also mentioned with their significance using independent T test and post hoc tests like LSD, Bonferroni and Tukeys tests. The OSF and OSCC study participants were age matched and there was no significant difference in the mean age. However there was significant difference in the mean age for OSCC and Controls and OSF and controls, as we could only get young participants in control group. Chewing habits were present in all the participants of OSF and OSCC groups with more than 5 years of frequent habit but only 14 percent of controls were having chewing habits and that to less than 5 years at lower frequency.

**Table 2.**
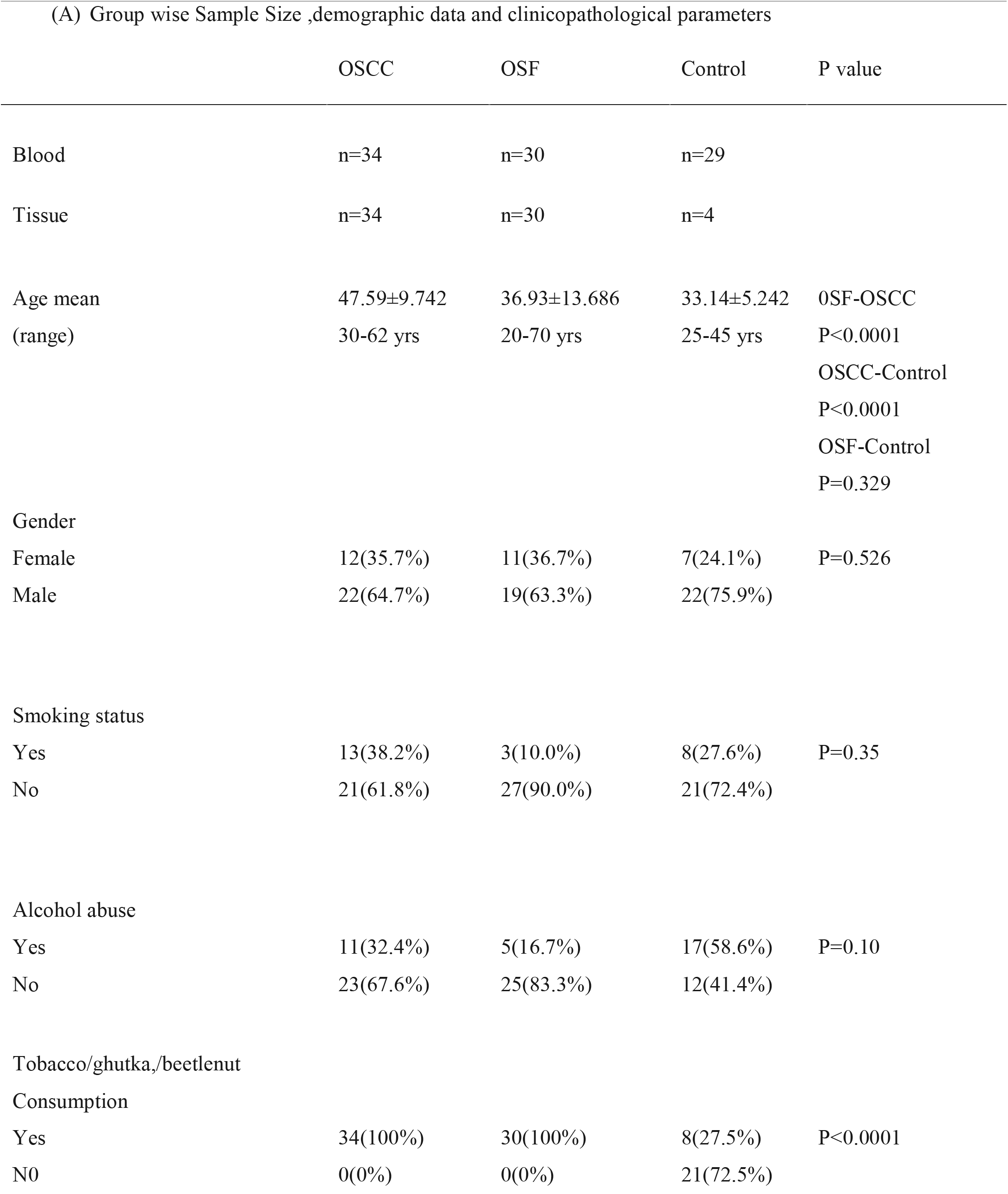
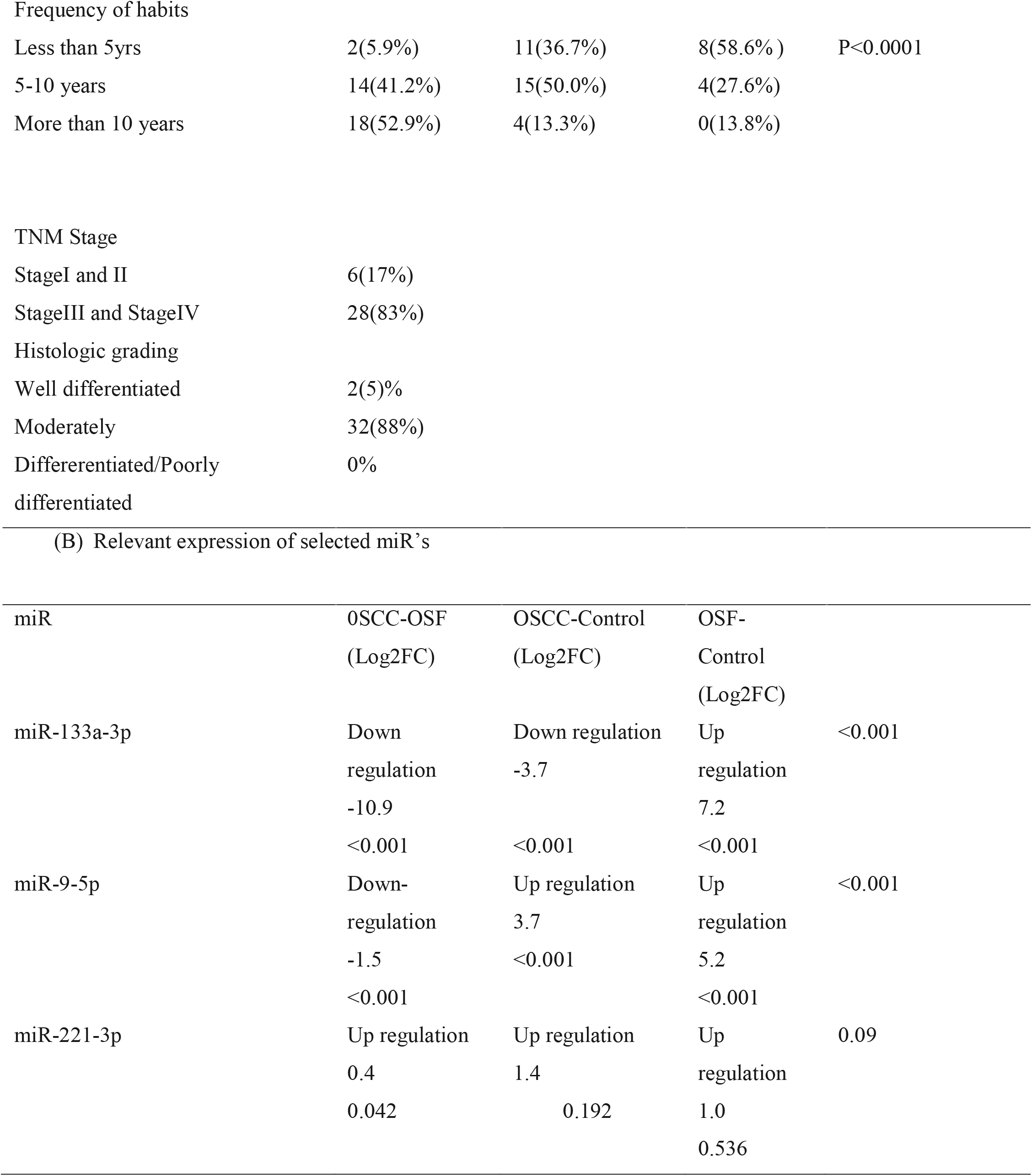
Group wise Sample size, demographic data and clinico pathological parameters (B) hsa-miR-133-3p, hsa-miR-221-5p and has-miR-9-5p expression in OSCC and OSF data compared to control from RT-qPCR

| (A) Group wise Sample Size ,demographic data and clinicopathological parameters |  |  |  |  |
| --- | --- | --- | --- | --- |
|  | OSCC | OSF | Control | P value |
| Blood | n=34 | n=30 | n=29 |  |
| Tissue | n=34 | n=30 | n=4 |  |
| Age mean | 47.59±9.742 | 36.93±13.686 | 33.14±5.242 | OSF-OSCC |
| (range) | 30-62 yrs | 20-70 yrs | 25-45 yrs | P<0.0001 |
|  |  |  |  | OSCC-Control |
|  |  |  |  | P<0.0001 |
|  |  |  |  | OSF-Control |
|  |  |  |  | P=0.329 |
| Gender |  |  |  |  |
| Female | 12(35.7%) | 11(36.7%) | 7(24.1%) | P=0.526 |
| Male | 22(64.7%) | 19(63.3%) | 22(75.9%) |  |
| Smoking status |  |  |  |  |
| Yes | 13(38.2%) | 3(10.0%) | 8(27.6%) | P=0.35 |
| No | 21(61.8%) | 27(90.0%) | 21(72.4%) |  |
| Alcohol abuse |  |  |  |  |
| Yes | 11(32.4%) | 5(16.7%) | 17(58.6%) | P=0.10 |
| No | 23(67.6%) | 25(83.3%) | 12(41.4%) |  |
| Tobacco/ghutka,/beetlenut |  |  |  |  |
| Consumption |  |  |  |  |
| Yes | 34(100%) | 30(100%) | 8(27.5%) | P<0.0001 |
| NO | 0(0%) | 0(0%) | 21(72.5%) |  |
| Frequency of habits |  |  |  |  |
| Less than 5yrs | 2(5.9%) | 11(36.7%) | 8(58.6%) | P<0.0001 |
| 5-10 years | 14(41.2%) | 15(50.0%) | 4(27.6%) |  |
| More than 10 years | 18(52.9%) | 4(13.3%) | 0(13.8%) |  |

|  |  |
| --- | --- |
| TNM Stage |  |
| StageI and II | 6(17%) |
| StageIII and StageIV | 28(83%) |
| Histologic grading |  |
| Well differentiated | 2(5)% |
| Moderately | 32(88%) |
| Differerentiated/Poorly differentiated | 0% |

(B) Relevant expression of selected miR's
| miR | OSCC-OSF<br>(Log2FC) | OSCC-Control<br>(Log2FC) | OSF-Control<br>(Log2FC) |  |
| --- | --- | --- | --- | --- |
| miR-133a-3p | Down<br>regulation<br>-10.9<br><0.001 | Down regulation<br>-3.7<br><0.001 | Up<br>regulation<br>7.2<br><0.001 | <0.001 |
| miR-9-5p | Down-<br>regulation<br>-1.5<br><0.001 | Up regulation<br>3.7<br><0.001 | Up<br>regulation<br>5.2<br><0.001 | <0.001 |
| miR-221-3p | Up regulation<br>0.4<br>0.042 | Up regulation<br>1.4<br>0.192 | Up<br>regulation<br>1.0<br>0.536 | 0.09 |

In the clinical samples, miR-9 was found to be up-regulated in OSF and OSCC compared to controls and between OSF and OSCC it was up-regulated in OSF with a logFC of 5.1, 3.6 and 1.5 respectively. miR-221 expression was found to be up-regulated in OSF and OSCC both compared to control but the values were not significant but a significant upregulation was seen in OSCC than controls with a Log FC of 1.4. miR-133-3p in our study was found to be significantly up-regulated in OSF and interestingly was down regulated in OSCC that to with a logFC value of 3.7 and 7.2 with significant p-value less than 0.001 respectively. The fold change expression are shown in **figure 5**

**Figure 5.**
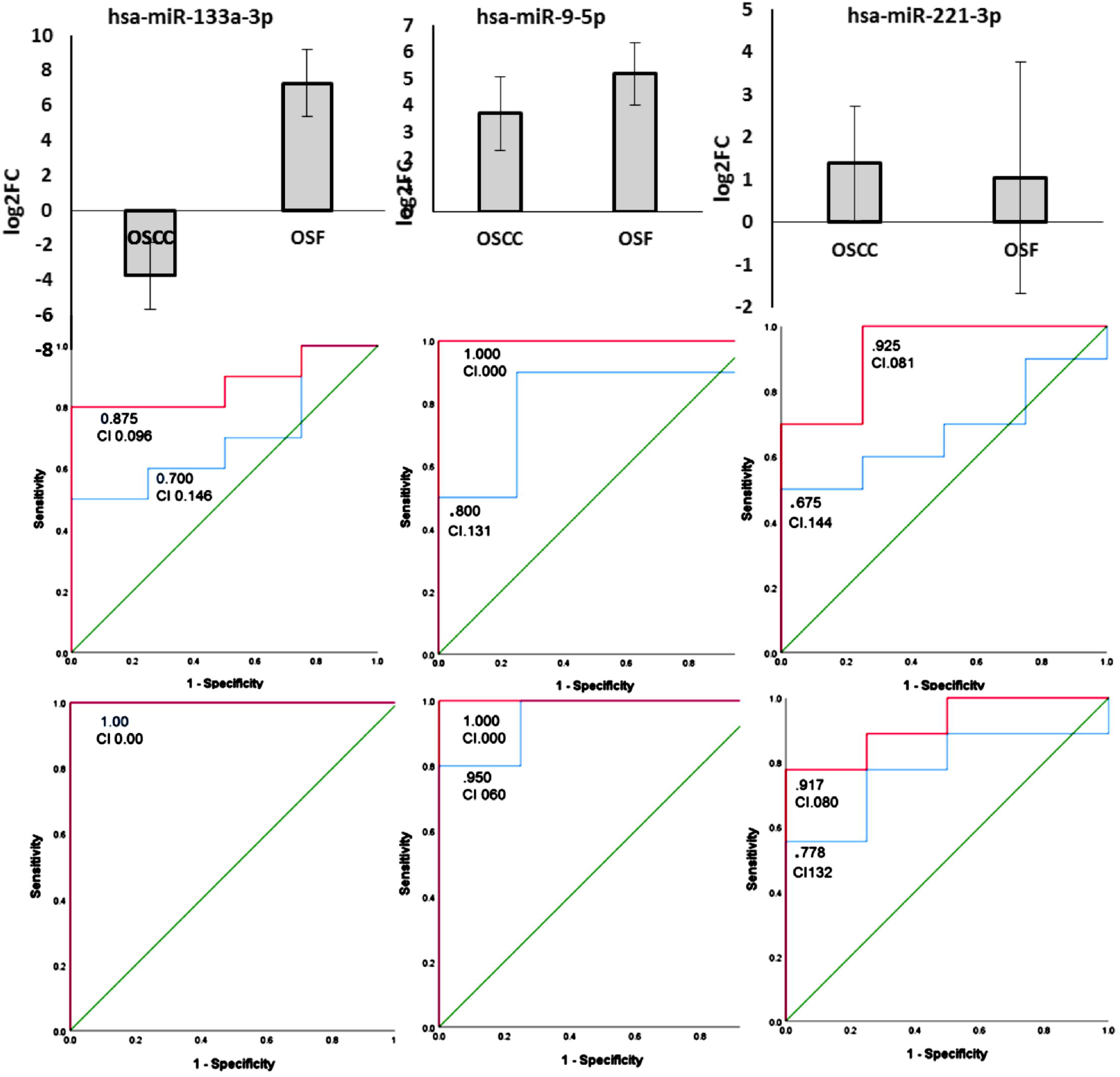
Log2Fc plots for OSF and OSCC expression compared to normal (top), ROC plot for OSCC vs Control (middle) and OSF Vs control (bottom) for blood and tissue ΔCt values for (A) miR-133a-3p (B) miR-9-5p (C) miR-221-3p

We have taken controls blood samples as only four control tissue samples were available during the study period so we compared the ΔCt values of tissues available with their respective blood for controls participants in each miR expression. They appeared to us within the error limits shown in **supplementary figure 3**. Further we compared ΔCt values of randomly chosen 10 blood and tissue samples for OSCC and OSF also but there were also no significant difference between the ΔCt values of blood and tissue values therefore, taking control blood samples for calculating fold change in miRs can be justified. We also constructed ROC for ΔCt valuesof OSCC and OSF compared to control for comparing specificity and sensitivity for each miR’s shown in **figure 5**. We obtained a comparable specificity and sensitivity for each miR between blood and tissue for each miR. So, the results look promising to be considered as biomarkers for diagnosing and demarcation of OSF to OSCC transition.

## DISCUSSION

OSCC is a preventable disease if diagnosed at earlier stages and OPMD’s transition is important role player towards the tumourogenesis of OSCC, in this article we tried to explore the altered expression of miR’s in OSF and OSCC expecting they can be used as risk stratification biomarkers. The role of miRs predicted in pathogenesis are discussed in earlier segments of the article. The qPCR expression data of selected miRs gives preliminary results about the involvement of these miRs in OSCC pathogenesis. miR-9 which is having a dynamic expression profile through different cancer types was found to be up-regulated in our study compared to controls that may be justified as miR-9 upregulation is involved in angiogenesis, metastasis in various cancer types [45] and also it was found to be up-regulated in OSF compared to OSCC which is an interesting outcome and probably supports the mechano-transduction of miR-9[46]as miR-9 is said to be down regulated due to mechanical compression in the tissue and may be OSF has a higher mechanical stress than OSCC due to very high extra cellular matrix (ECM) deposition. However, mechanistic studies using cell lines and animal models may be useful in validating the course of pathogenesis. miR-133 was significantly downregulated in OSCC similar to previous studies by Chattopadhay *et al*[39]. They also observed an upregulation in oral leukoplakia but they could not find significant difference between the groups. In our study groupwe observed a significant upregulation of miR-133-a-3p in OSF compared to OSCC and Controls with a Log Fc value of 10.9 and 7.2 with p value <0.001 respectively. A significant role of miR-133a-3p in OSF to OSCC pathogenesis may be present this may be because collagen associated genes COL1A1, COL25A1 are direct targets of mir133a-3p shown in string PPI networks even if they are not enriched. Previous studies have shown that upregulation of these genes increases the collagen production which is mainly involved in OSF pathogenesis[50]mir-133a-3p acts as positive regulator of apoptosis so its downregulation in OSCC may be justified hence it can act as double marker for OSF to OSCC pathogenesis.

The sample Ct data for controls and OSF were scattered for miR-221. A categorical comparison may be helpful to see the difference in expression compared to age, sex etc. to obtain a personalized diagnostic tests in OSF but unfortunately we could not do that because of small sample size and the differences in mean age between the OSF and control groups. The targets of mir-221 obtained from our enrichment study were mostly chaperonins family members such as TCP-1, CCT-3, CCT6A are involved in the folding of various proteins like telomere maintenance protein, ubiquitin pathway proteins, actin-tubulin proteins and alternate transcriptional splice variants[51], also miR-221 is reported to be elevated in many cancers such as hepatocellular, gastric, breast and lung squamous cell carcinoma with targets such as SOD2, MMP1 genes which are also pre dominantly studied in OSCC [52]. Involvement of miR-221 in multiple cancer associated processes make it a putative therapeutic target. Therefore, we suggest miR-221-3p need to be explored more with a larger categorical data in OSCC and OSF. The current study suggests that transition from OSF to OSCC may be regulated by complex miR’s interaction with their target genes and specifically the significant expression of miR-133a-3p, miR9-5p makes them useful as biomarkers for early diagnosis of OSCC in OSF patients, as is also reinforced by the sensitivity and specificity observations of the current study in **Figure 5**. These maybe explored further using experimental validation for the target genes associated to uncover the overall miR role in pathogenesis of OSF to OSCC for early diagnosis and hence better prognosis. The dysregulated miRs in OSFmay be used as signature for predicting OSCC and this group of OSF patients may be followed up and should be advised for regular examinations. Large experimental dataset platforms such as Microarray datasets, TCGA database should be exploited to uncover the genome and epigenome wide associations involved for pathogenic pathways in OSC. The miRs predicted should be validated for their target identification experimentally so as to identify their uniquebiomarkers and putative drug targets in OSFto OSCC pathogenesis for early diagnosis and cure.

## Supporting information

complete suppli

## Data Availability

supplimentary data

## ABBREVATIONS

OSCC: Oral squamous cell carcinoma
HNSCC: Head and neck cancer
OPMDs: Oral potentially malignant disorders
OSF: Oral submucous fibrosis
EMT: Epithelial mesenchymal transition
HPV: Human papilloma virus
miR’s: microRNA’s

