## Supplementary material for "In-silico, interactomic and clinical validation based approach for screening and identification of miR biomarkers involved in Oral submucous fibrosis to Oral squamous cell carcinoma transition": complete suppli: Suppliment for AACR .docx

**Supplementary data**

| **Content** | **Description** | **Page#** |
| --- | --- | --- |
| **Table S1** | DS1, DS2 and DS3 miRs listed and miRs identified (ID) and not identified(un-ID) by miEEA and MINURNET in enrichment analyses | **2-8** |
| **Table S2** | miRs with differential expression in cancer specific databases miRCancer, ONCOMIR, and HNSCCval | **9-13** |
| **Table S3** | Cluster analyses using miRCancer | **13-27** |
| **Table S4** | Over representation analyses of three datasets in brief. | **27-28** |
| **Fig S1** | Results of miR enrichment analyses | **29** |
| **Table S5** | Targets obtained from Target scan with their cutoff values | **29-38** |
| **Fig S2**  **Fig S3** | PPI Network using STRING and CYTOSCAPE  Scatter plot for ∆ct value of tissue and blood | **39**  **40** |

**Table S1** DS1, DS2 and DS3 miRs listed and miRs identified (ID) and not identified(un-ID) by miEEA and MINURNET in enrichment analyses

| DS-1(ID) | DS-1(un-ID) | DS-2(ID) | all DS-2(un-ID) | DS-3(ID) | all DS-3(un-ID) |
| --- | --- | --- | --- | --- | --- |
| hsa-miR-5692a | hsa-miR-1252 | hsa-miR-3686 | hsa-miR-6755 | hsa-miR-4419b | hsa-miR-877 |
| hsa-miR-4464 | hsa-miR-3913 | hsa-miR-1305 | hsa-miR-135b | hsa-miR-3929 | hsa-miR-4747 |
| hsa-miR-4311 | hsa-miR-4703 | hsa-miR-4282 | hsa-miR-6815 | hsa-miR-4478 | hsa-miR-5196 |
| hsa-miR-4490 | hsa-miR-4716 | hsa-miR-3135b | hsa-miR-6884 | hsa-miR-548av | hsa-miR-548a |
| hsa-miR-8055 | hsa-miR-4778 | hsa-miR-3128 | hsa-miR-33a | hsa-miR-3125 | hsa-miR-188 |
| hsa-miR-4643 | hsa-miR-545 | hsa-miR-3609 | hsa-miR-1910 | hsa-miR-3612 | hsa-miR-3156 |
| hsa-miR-562 | hsa-miR-26b | hsa-miR-3662 | hsa-miR-20a | hsa-miR-3685 | hsa-miR-4446 |
| hsa-miR-1323 | hsa-miR-4797 | hsa-miR-577 | hsa-miR-26a | hsa-miR-3714 | hsa-miR-520a |
| hsa-miR-6074 | hsa-miR-138 | hsa-miR-1976 | hsa-miR-485 | hsa-miR-587 | hsa-miR-525 |
| hsa-miR-4291 | hsa-miR-149 | hsa-miR-4729 | hsa-miR-6739 | hsa-miR-650 | hsa-miR-548g |
| hsa-miR-3671 | hsa-miR-7109 | hsa-miR-548p | hsa-miR-30a | hsa-miR-3188 | hsa-miR-5584 |
| hsa-miR-466 | hsa-miR-21 | hsa-miR-759 | hsa-miR-4668 | hsa-miR-4698 | hsa-miR-580 |
| hsa-miR-3122 | hsa-miR-32 | hsa-miR-940 | hsa-miR-494 | hsa-miR-3658 | hsa-miR-7159 |
| hsa-miR-5094 | hsa-miR-335 | hsa-miR-4748 | hsa-miR-506 | hsa-miR-922 | hsa-miR-744 |
| hsa-miR-600 | hsa-miR-3607 | hsa-miR-646 | hsa-miR-548c | hsa-miR-3646 | hsa-miR-3944 |
| hsa-miR-3652 | hsa-miR-548o | hsa-miR-4297 | hsa-miR-892c | hsa-miR-4692 | hsa-miR-371b |
| hsa-miR-5695 | hsa-miR-153 | hsa-miR-484 | hsa-miR-30c | hsa-miR-8063 | hsa-miR-7112 |
| hsa-miR-3153 | hsa-miR-10a | hsa-miR-936 | hsa-miR-889 | hsa-miR-320b | hsa-miR-190a |
| hsa-miR-636 | hsa-miR-627 | hsa-miR-5688 | hsa-miR-4659a | hsa-miR-320c | hsa-miR-3922 |
| hsa-miR-4470 | hsa-miR-151a | hsa-miR-107 | hsa-miR-4659b | hsa-miR-320d | hsa-miR-6768 |
| hsa-miR-4487 | hsa-miR-5589 | hsa-miR-558 | hsa-miR-6875 | hsa-miR-3689e | hsa-miR-33b |
| hsa-miR-4418 | hsa-miR-15a | hsa-miR-4430 | hsa-miR-6511a | hsa-miR-3689f | hsa-miR-3664 |
| hsa-miR-2053 | hsa-miR-15b | hsa-miR-4679 | hsa-miR-2682 | hsa-miR-4429 | hsa-miR-3680 |
| hsa-miR-5681b | hsa-miR-195 |  | hsa-miR-4735 | hsa-miR-4506 | hsa-miR-651 |
| hsa-miR-5697 | hsa-miR-181c |  | hsa-miR-6808 | hsa-miR-5681a | hsa-miR-6809 |
| hsa-miR-4310 | hsa-miR-155 |  | hsa-miR-6890 | hsa-miR-5692b | hsa-miR-942 |
| hsa-miR-3935 | hsa-miR-127 |  | hsa-miR-6893 | hsa-miR-5692c | hsa-miR-1207 |
| hsa-miR-451a | hsa-miR-4691 |  | hsa-miR-16 | hsa-miR-761 | hsa-miR-27a |
| hsa-miR-1322 | hsa-miR-1226 |  | hsa-miR-3529 | hsa-miR-4459 | hsa-miR-3190 |
| hsa-miR-4463 | hsa-miR-1301 |  | hsa-miR-5003 | hsa-miR-1321 | hsa-miR-330 |
| hsa-miR-607 | hsa-miR-192 |  | hsa-miR-625 | hsa-miR-4441 | hsa-miR-3606 |
| hsa-miR-3148 | hsa-miR-376a |  | hsa-miR-4789 | hsa-miR-4526 | hsa-miR-5088 |
| hsa-miR-4469 | hsa-miR-376b |  | hsa-miR-6765 | hsa-miR-548aa | hsa-miR-6878 |
| hsa-miR-4499 | hsa-miR-495 |  | hsa-miR-519d | hsa-miR-5191 | hsa-miR-1271 |
| hsa-miR-2113 | hsa-miR-210 |  | hsa-miR-3613 | hsa-miR-4261 | hsa-miR-1343 |
| hsa-miR-935 | hsa-miR-22 |  | hsa-miR-497 | hsa-miR-527 | hsa-miR-222 |
| 2hsa-miR-3668 | hsa-miR-3622b |  | hsa-miR-3160 | hsa-miR-635 | hsa-miR-23b |
|  | hsa-miR-7157 |  | hsa-miR-92a | hsa-miR-4419a | hsa-miR-3192 |
|  | hsa-miR-6749 |  | hsa-miR-17 | hsa-miR-6133 | hsa-miR-3194 |
|  | hsa-miR-152 |  | hsa-miR-186 | hsa-miR-4510 | hsa-miR-320a |
|  | hsa-miR-3622a |  | hsa-miR-20b | hsa-miR-6127 | hsa-miR-323a |
|  | hsa-miR-18a |  | hsa-miR-299 | hsa-miR-6129 | hsa-miR-3689a |
|  | hsa-miR-449c |  | hsa-miR-582 | hsa-miR-6130 | hsa-miR-3689b |
|  | hsa-miR-6807 |  | hsa-miR-6504 | hsa-miR-6072 | hsa-miR-371a |
|  | hsa-miR-140 |  | hsa-miR-93 | hsa-miR-5787 | hsa-miR-450b |
|  | hsa-miR-25 |  | hsa-miR-1225 | hsa-miR-3689d | hsa-miR-4645 |
|  | hsa-miR-302a |  | hsa-miR-6784 | hsa-miR-3907 | hsa-miR-4727 |
|  | hsa-miR-576 |  | hsa-miR-4524b | hsa-miR-4514 | hsa-miR-4753 |
|  | hsa-miR-3074 |  | hsa-miR-34b | hsa-miR-1193 | hsa-miR-4999 |
|  | hsa-miR-101 |  | hsa-miR-542 | hsa-miR-1254 | hsa-miR-548az |
|  | hsa-miR-18b |  | hsa-miR-329 | hsa-miR-4438 | hsa-miR-548e |
|  | hsa-miR-363 |  | hsa-miR-4524a | hsa-miR-4530 | hsa-miR-548t |
|  | hsa-miR-367 |  | hsa-miR-519c | hsa-miR-5683 | hsa-miR-550a |
|  | hsa-miR-589 |  | hsa-miR-106b | hsa-miR-3660 | hsa-miR-6730 |
|  | hsa-miR-6831 |  | hsa-miR-7 | hsa-miR-3914 | hsa-miR-6814 |
|  | hsa-miR-4677 |  | hsa-miR-6513 | hsa-miR-6081 | hsa-miR-1298 |
|  | hsa-miR-4670 |  | hsa-miR-106a | hsa-miR-1184 | hsa-miR-4717 |
|  | hsa-miR-450a |  | hsa-miR-6733 | hsa-miR-3123 | hsa-miR-4731 |
|  | hsa-miR-6792 |  | hsa-miR-1229 | hsa-miR-4682 | hsa-miR-4733 |
|  | hsa-miR-7b |  | hsa-miR-3691 | hsa-miR-520h | hsa-miR-6780a |
|  | hsa-miR-6806 |  | hsa-miR-1468 | hsa-miR-622 | hsa-miR-7152 |
|  | hsa-miR-4662a |  | hsa-miR-3940 | hsa-miR-8062 | hsa-miR-203b |
|  | hsa-miR-4799 |  | hsa-miR-887 | hsa-miR-4472 | hsa-miR-296 |
|  | hsa-miR-1911 |  | hsa-miR-6838 | hsa-miR-4316 | hsa-miR-374a |
|  | hsa-miR-503 |  | hsa-miR-103a | hsa-miR-4447 | hsa-miR-4695 |
|  | hsa-miR-425 |  | hsa-miR-3121 | hsa-miR-4660 | hsa-miR-4756 |
|  | hsa-miR-548ah |  | hsa-miR-218 | hsa-miR-7977 | hsa-miR-4768 |
|  | hsa-miR-199a |  | hsa-miR-424 | hsa-miR-1324 | hsa-miR-548ap |
|  | hsa-miR-199b |  | hsa-miR-1233 | hsa-miR-6165 | hsa-miR-6515 |
|  | hsa-miR-34a |  | hsa-miR-3927 | hsa-miR-1182 | hsa-miR-6721 |
|  | hsa-miR-150 |  | hsa-miR-3129 | hsa-miR-4257 | hsa-miR-7160 |
|  | hsa-miR-5581 |  | hsa-miR-144 | hsa-miR-5693 | hsa-miR-548aj |
|  | hsa-miR-215 |  | hsa-miR-504 | hsa-miR-3164 | hsa-miR-548f |
|  | hsa-miR-375 |  | hsa-miR-708 | hsa-miR-657 | hsa-miR-548x |
|  |  |  | hsa-miR-509 | hsa-miR-4505 | hsa-miR-524 |
|  |  |  | hsa-miR-135a | hsa-miR-4736 | hsa-miR-4709 |
|  |  |  | hsa-miR-5089 | hsa-miR-3143 | hsa-miR-539 |
|  |  |  |  | hsa-miR-6077 | hsa-miR-605 |
|  |  |  |  | hsa-miR-6124 | hsa-miR-6777 |
|  |  |  |  | hsa-miR-1293 | hsa-miR-200b |
|  |  |  |  | hsa-miR-4803 | hsa-miR-27b |
|  |  |  |  | hsa-miR-4268 | hsa-miR-30d |
|  |  |  |  | hsa-miR-3138 | hsa-miR-30e |
|  |  |  |  | hsa-miR-3659 | hsa-miR-410 |
|  |  |  |  | hsa-miR-4267 | hsa-miR-4482 |
|  |  |  |  | hsa-miR-7641 | hsa-miR-514a |
|  |  |  |  | hsa-miR-4434 | hsa-miR-518a |
|  |  |  |  | hsa-miR-5703 | hsa-miR-548au |
|  |  |  |  | hsa-miR-4443 | hsa-miR-548d |
|  |  |  |  | hsa-miR-4476 | hsa-miR-548h |
|  |  |  |  | hsa-miR-8069 | hsa-miR-556 |
|  |  |  |  | hsa-miR-133b | hsa-miR-6774 |
|  |  |  |  | hsa-miR-4752 | hsa-miR-6873 |
|  |  |  |  | hsa-miR-548ac | hsa-miR-30b |
|  |  |  |  | hsa-miR-548z | hsa-miR-3140 |
|  |  |  |  | hsa-miR-4302 | hsa-miR-338 |
|  |  |  |  | hsa-miR-4477a | hsa-miR-4777 |
|  |  |  |  | hsa-miR-6134 | hsa-miR-6508 |
|  |  |  |  | hsa-miR-3672 | hsa-miR-6813 |
|  |  |  |  | hsa-miR-3909 | hsa-miR-423 |
|  |  |  |  | hsa-miR-4252 | hsa-miR-6734 |
|  |  |  |  | hsa-miR-3674 | hsa-miR-767 |
|  |  |  |  | hsa-miR-4739 | hsa-miR-1250 |
|  |  |  |  | hsa-miR-4641 | hsa-miR-146b |
|  |  |  |  | hsa-miR-4513 | hsa-miR-5001 |
|  |  |  |  | hsa-miR-5096 | hsa-miR-619 |
|  |  |  |  | hsa-miR-764 | hsa-miR-6775 |
|  |  |  |  | hsa-miR-3924 | hsa-miR-874 |
|  |  |  |  | hsa-miR-3118 | hsa-miR-1277 |
|  |  |  |  | hsa-miR-4706 | hsa-miR-204 |
|  |  |  |  | hsa-miR-4662b | hsa-miR-3173 |
|  |  |  |  | hsa-miR-4535 | hsa-miR-362 |
|  |  |  |  | hsa-miR-3975 | hsa-miR-383 |
|  |  |  |  | hsa-miR-4318 | hsa-miR-4728 |
|  |  |  |  | hsa-miR-4325 | hsa-miR-500b |
|  |  |  |  | hsa-miR-198 | hsa-miR-5187 |
|  |  |  |  | hsa-miR-3135a | hsa-miR-6715b |
|  |  |  |  | hsa-miR-1264 | hsa-miR-6735 |
|  |  |  |  | hsa-miR-1283 | hsa-miR-6738 |
|  |  |  |  | hsa-miR-3171 | hsa-miR-6743 |
|  |  |  |  | hsa-miR-4424 | hsa-miR-6785 |
|  |  |  |  | hsa-miR-4465 | hsa-miR-6799 |
|  |  |  |  | hsa-miR-4705 | hsa-miR-6829 |
|  |  |  |  | hsa-miR-3147 | hsa-miR-6848 |
|  |  |  |  | hsa-miR-5092 | hsa-miR-6870 |
|  |  |  |  | hsa-miR-8079 | hsa-miR-6882 |
|  |  |  |  | hsa-miR-4480 | hsa-miR-4722 |
|  |  |  |  | hsa-miR-4501 | hsa-miR-6780b |
|  |  |  |  | hsa-miR-3133 | hsa-miR-10b |
|  |  |  |  |  | hsa-miR-1914 |
|  |  |  |  |  | hsa-miR-211 |
|  |  |  |  |  | hsa-miR-3663 |
|  |  |  |  |  | hsa-miR-7854 |
|  |  |  |  |  | hsa-miR-205 |
|  |  |  |  |  | hsa-miR-433 |
|  |  |  |  |  | hsa-miR-491 |
|  |  |  |  |  | hsa-miR-5189 |
|  |  |  |  |  | hsa-miR-6716 |
|  |  |  |  |  | hsa-miR-483 |
|  |  |  |  |  | hsa-miR-200a |
|  |  |  |  |  | hsa-miR-3120 |
|  |  |  |  |  | hsa-miR-361 |
|  |  |  |  |  | hsa-miR-4755 |
|  |  |  |  |  | hsa-miR-6510 |
|  |  |  |  |  | hsa-miR-6794 |
|  |  |  |  |  | hsa-miR-6818 |
|  |  |  |  |  | hsa-miR-6852 |
|  |  |  |  |  | hsa-miR-148a |
|  |  |  |  |  | hsa-miR-579 |
|  |  |  |  |  | hsa-miR-134 |
|  |  |  |  |  | hsa-miR-1912 |
|  |  |  |  |  | hsa-miR-4690 |
|  |  |  |  |  | hsa-miR-6876 |
|  |  |  |  |  | hsa-miR-7845 |
|  |  |  |  |  | hsa-miR-508 |
|  |  |  |  |  | hsa-miR-520g |
|  |  |  |  |  | hsa-miR-340 |
|  |  |  |  |  | hsa-miR-4639 |
|  |  |  |  |  | hsa-miR-4671 |
|  |  |  |  |  | hsa-miR-512 |
|  |  |  |  |  | hsa-miR-6756 |
|  |  |  |  |  | hsa-miR-4474 |
|  |  |  |  |  | hsa-miR-6867 |
|  |  |  |  |  | hsa-miR-3189 |
|  |  |  |  |  | hsa-miR-6847 |
|  |  |  |  |  | hsa-miR-7162 |
|  |  |  |  |  | hsa-miR-380 |
|  |  |  |  |  | hsa-miR-5004 |
|  |  |  |  |  | hsa-miR-574 |
|  |  |  |  |  | hsa-miR-6754 |
|  |  |  |  |  | hsa-miR-5010 |
|  |  |  |  |  | hsa-miR-34c |
|  |  |  |  |  | hsa-miR-3619 |
|  |  |  |  |  | hsa-miR-449b |
|  |  |  |  |  | hsa-miR-4761 |
|  |  |  |  |  | hsa-miR-3679 |
|  |  |  |  |  | hsa-miR-3127 |
|  |  |  |  |  | hsa-miR-130a |
|  |  |  |  |  | hsa-miR-4666a |
|  |  |  |  |  | hsa-miR-516a |
|  |  |  |  |  | hsa-miR-516b |
|  |  |  |  |  | hsa-miR-519a |
|  |  |  |  |  | hsa-miR-519b |
|  |  |  |  |  | hsa-miR-1255b |
|  |  |  |  |  | hsa-miR-642a |
|  |  |  |  |  | hsa-miR-4758 |
|  |  |  |  |  | hsa-miR-5699 |
|  |  |  |  |  | hsa-miR-6840 |
|  |  |  |  |  | hsa-miR-6877 |
|  |  |  |  |  | hsa-miR-126 |
|  |  |  |  |  | hsa-miR-377 |
|  |  |  |  |  | hsa-miR-4720 |
|  |  |  |  |  | hsa-miR-6732 |
|  |  |  |  |  | hsa-miR-6805 |
|  |  |  |  |  | hsa-miR-4766 |
|  |  |  |  |  | hsa-miR-624 |
|  |  |  |  |  | hsa-miR-6835 |
|  |  |  |  |  | hsa-miR-143 |
|  |  |  |  |  | hsa-miR-6499 |
|  |  |  |  |  | hsa-miR-7849 |
|  |  |  |  |  | hsa-miR-7156 |
|  |  |  |  |  | hsa-miR-1238 |
|  |  |  |  |  | hsa-miR-3688 |
|  |  |  |  |  | hsa-miR-372 |
|  |  |  |  |  | hsa-miR-4796 |
|  |  |  |  |  | hsa-miR-520b |
|  |  |  |  |  | hsa-miR-520c |
|  |  |  |  |  | hsa-miR-520e |
|  |  |  |  |  | hsa-miR-6868 |
|  |  |  |  |  | hsa-miR-6894 |
|  |  |  |  |  | hsa-miR-769 |
|  |  |  |  |  | hsa-miR-4800 |
|  |  |  |  |  | hsa-miR-513a |
|  |  |  |  |  | hsa-miR-664b |
|  |  |  |  |  | hsa-miR-331 |
|  |  |  |  |  | hsa-miR-541 |
|  |  |  |  |  | hsa-miR-3187 |
|  |  |  |  |  | hsa-miR-4723 |
|  |  |  |  |  | hsa-miR-6769b |
|  |  |  |  |  | hsa-miR-3591 |
|  |  |  |  |  | hsa-miR-412 |
|  |  |  |  |  | hsa-miR-6791 |
|  |  |  |  |  | hsa-miR-6871 |
|  |  |  |  |  | hsa-miR-6855 |
|  |  |  |  |  | hsa-miR-1178 |
|  |  |  |  |  | hsa-miR-4676 |
|  |  |  |  |  | hsa-miR-6512 |
|  |  |  |  |  | hsa-miR-4536 |
|  |  |  |  |  | hsa-miR-5582 |
|  |  |  |  |  | hsa-miR-6821 |
|  |  |  |  |  | hsa-miR-183 |
|  |  |  |  |  | hsa-miR-4763 |
|  |  |  |  |  | hsa-miR-6720 |
|  |  |  |  |  | hsa-miR-3616 |
|  |  |  |  |  | hsa-miR-520f |
|  |  |  |  |  | hsa-miR-552 |
|  |  |  |  |  | hsa-miR-19a |
|  |  |  |  |  | hsa-miR-19b |
|  |  |  |  |  | hsa-miR-6500 |
|  |  |  |  |  | hsa-miR-6507 |
|  |  |  |  |  | hsa-miR-154 |
|  |  |  |  |  | hsa-miR-3151 |
|  |  |  |  |  | hsa-miR-4700 |
|  |  |  |  |  | hsa-miR-6727 |
|  |  |  |  |  | hsa-miR-6776 |
|  |  |  |  |  | hsa-miR-7114 |
|  |  |  |  |  | hsa-miR-191 |
|  |  |  |  |  | hsa-miR-6726 |
|  |  |  |  |  | hsa-miR-551b |
|  |  |  |  |  | hsa-miR-4713 |
|  |  |  |  |  | hsa-miR-486 |
|  |  |  |  |  | hsa-miR-2467 |
|  |  |  |  |  | hsa-miR-4684 |
|  |  |  |  |  | hsa-miR-660 |
|  |  |  |  |  | hsa-miR-208a |
|  |  |  |  |  | hsa-miR-6820 |
|  |  |  |  |  | hsa-miR-301a |
|  |  |  |  |  | hsa-miR-888 |
|  |  |  |  |  | hsa-miR-1296 |
|  |  |  |  |  | hsa-miR-4697 |
|  |  |  |  |  | hsa-miR-6802 |
|  |  |  |  |  | hsa-miR-1288 |
|  |  |  |  |  | hsa-miR-3682 |
|  |  |  |  |  | hsa-miR-7155 |
|  |  |  |  |  | hsa-miR-6753 |
|  |  |  |  |  | hsa-miR-31 |
|  |  |  |  |  | hsa-miR-124 |
|  |  |  |  |  | hsa-miR-6722 |
|  |  |  |  |  | hsa-miR-28 |
|  |  |  |  |  | hsa-miR-6747 |
|  |  |  |  |  | hsa-miR-3144 |
|  |  |  |  |  | hsa-miR-4680 |
|  |  |  |  |  | hsa-miR-4633 |
|  |  |  |  |  | hsa-miR-1180 |
|  |  |  |  |  | hsa-miR-454 |
|  |  |  |  |  | hsa-miR-6895 |
|  |  |  |  |  | hsa-miR-1287 |
|  |  |  |  |  | hsa-miR-4757 |
|  |  |  |  |  | hsa-miR-487b |
|  |  |  |  |  | hsa-miR-3130 |
|  |  |  |  |  | hsa-miR-7851 |

**Table S2** miRs with differential expression in cancer specific databases miRCancer, ONCOMIR, and HNSCCval with FC more than 1.5 and p value less than 0.05.

| ONCOMIR | miRCancer | HNSCC val |  |  |
| --- | --- | --- | --- | --- |
| hsa-miR-33a | hsa-let-7d | hsa-miR-6790 |  |  |
| hsa-miR-34b-5p | hsa-miR-1 | hsa-miR-139 |  |  |
| hsa-miR-1468-5p | hsa-miR-133a | hsa-miR-378a |  |  |
| hsa-miR-188-5p | hsa-miR-205 | hsa-miR-1277 |  |  |
| hsa-miR-501-5p | hsa-miR-21 | hsa-miR-153 |  |  |
| hsa-miR-615-3p | hsa-miR-100 | hsa-miR-548m |  |  |
| hsa-miR-877-5p | hsa-miR-125b | hsa-miR-6808 |  |  |
| hsa-miR-3150b-3p | hsa-miR-373 | hsa-miR-29b |  |  |
| hsa-miR-655-3p | hsa-miR-138 | hsa-miR-17 |  |  |
| hsa-miR-18a-3p | hsa-miR-24 | hsa-miR-6810 |  |  |
| hsa-miR-483-3p | hsa-miR-106b | hsa-miR-4789 |  |  |
| hsa-miR-3687 | hsa-miR-25 | hsa-miR-26b |  |  |
| hsa-miR-99a-3p | hsa-miR-375 | hsa-miR-302d |  |  |
| hsa-miR-526b-5p | hsa-miR-93 | hsa-miR-891b |  |  |
| hsa-miR-519a-5p | hsa-miR-16 | hsa-miR-30a |  |  |
| hsa-miR-512-3p | hsa-miR-30a-5p | hsa-miR-4721 |  |  |
| hsa-miR-508-5p | hsa-miR-106a | hsa-miR-497 |  |  |
| hsa-miR-671-3p | hsa-miR-148a | hsa-miR-4516 |  |  |
| hsa-miR-421 | hsa-miR-218 | hsa-miR-548c |  |  |
| hsa-miR-150-3p | hsa-miR-210 | hsa-miR-6825 |  |  |
| hsa-miR-215-5p | hsa-miR-124 | hsa-miR-548aq |  |  |
| hsa-miR-431-5p | hsa-miR-92a | hsa-miR-518c |  |  |
| hsa-miR-1271-5p | hsa-miR-181a | hsa-miR-3665 |  |  |
| hsa-miR-138-5p | hsa-miR-200c | hsa-miR-16 |  |  |
| hsa-miR-592 | hsa-miR-221 | hsa-miR-548ag |  |  |
| hsa-miR-154-5p | hsa-miR-222 | hsa-miR-548as |  |  |
| hsa-miR-323b-3p | hsa-miR-17 | hsa-miR-21 |  |  |
| hsa-miR-3648 | hsa-miR-18a | hsa-miR-1266 |  |  |
| hsa-miR-516a-5p | hsa-miR-19a | hsa-miR-612 |  |  |
| hsa-miR-550a-5p | hsa-miR-19b | hsa-miR-20b |  |  |
| hsa-miR-338-5p | hsa-miR-20a | hsa-miR-548u |  |  |
| hsa-miR-224-3p | hsa-miR-203 | hsa-miR-7b |  |  |
| hsa-miR-323a-3p | hsa-miR-133a | hsa-miR-889 |  |  |
| hsa-miR-383-5p | hsa-miR-133b | hsa-miR-5584 |  |  |
| hsa-miR-101-5p | hsa-miR-145 | hsa-miR-6768 |  |  |
| hsa-miR-330-3p | hsa-miR-29c | hsa-miR-503 |  |  |
| hsa-miR-219a-1-3p | hsa-let-7b | hsa-miR-6749 |  |  |
| hsa-miR-3065-5p | hsa-let-7c | hsa-miR-1225 |  |  |
| hsa-miR-296-5p | hsa-let-7e | hsa-miR-3681 |  |  |
| hsa-miR-26a-2-3p | hsa-let-7g | hsa-miR-4729 |  |  |
| ONCOMIR | miRCancer | HNSCC val |  |  |
| hsa-miR-369-5p | hsa-miR-31 | hsa-miR-15a |  |  |
| hsa-miR-3127-5p | hsa-miR-206 | hsa-miR-3201 |  |  |
| hsa-miR-33b-5p | hsa-miR-107 | hsa-miR-130b |  |  |
| hsa-miR-1266-5p | hsa-miR-34a | hsa-miR-3192 |  |  |
| hsa-miR-424-3p | hsa-miR-214 | hsa-miR-155 |  |  |
| hsa-miR-485-3p | hsa-miR-126 | hsa-miR-4796 |  |  |
| hsa-let-7f-1-3p | hsa-miR-518b | hsa-miR-373 |  |  |
| hsa-miR-940 | hsa-miR-150 | hsa-miR-205 |  |  |
| hsa-miR-937-3p | hsa-miR-196b | hsa-miR-3648 |  |  |
| hsa-miR-651-5p | hsa-miR-22 | hsa-miR-100 |  |  |
| hsa-miR-326 | hsa-miR-370 | hsa-miR-514a |  |  |
| hsa-miR-34b-3p | hsa-miR-363 | hsa-miR-130a |  |  |
| hsa-miR-500a-5p | hsa-miR-149 | hsa-miR-93 |  |  |
| hsa-miR-500b-5p | hsa-miR-143 | hsa-miR-196a |  |  |
| hsa-miR-7-5p | hsa-miR-99a | hsa-miR-27a |  |  |
| hsa-miR-487b-3p | hsa-miR-874 | hsa-miR-7c |  |  |
| hsa-miR-539-5p | hsa-miR-134 | hsa-miR-4464 |  |  |
| hsa-miR-891a-5p | hsa-miR-27a* | hsa-miR-5581 |  |  |
| hsa-miR-653-5p | hsa-miR-181b | hsa-miR-3165 |  |  |
| hsa-let-7a-2-3p | hsa-miR-345 | hsa-miR-191 |  |  |
| hsa-miR-577 | hsa-miR-185 | hsa-miR-3689b |  |  |
| hsa-miR-370-3p | hsa-miR-195 | hsa-miR-4310 |  |  |
| hsa-miR-1296-5p | hsa-miR-200b | hsa-miR-6731 |  |  |
| hsa-miR-3653 | hsa-miR-29a | hsa-miR-222 |  |  |
| hsa-miR-299-5p | hsa-miR-9 | hsa-miR-5681b |  |  |
| hsa-miR-493-3p | hsa-miR-27a | hsa-miR-603 |  |  |
| hsa-miR-29a-5p | hsa-miR-129-5p | hsa-miR-340 |  |  |
| hsa-miR-9-3p | hsa-let-7a | hsa-miR-5006 |  |  |
| hsa-miR-340-3p | hsa-miR-146a | hsa-miR-942 |  |  |
| hsa-miR-629-3p | hsa-miR-146b | hsa-miR-195 |  |  |
| hsa-miR-3677-3p | hsa-miR-655 | hsa-miR-190b |  |  |
| hsa-miR-505-5p | hsa-miR-223 | hsa-miR-106a |  |  |
| hsa-miR-16-1-3p | hsa-miR-302b | hsa-miR-367 |  |  |
| hsa-miR-590-3p | hsa-miR-155 | hsa-miR-519d |  |  |
| hsa-miR-125a-3p | hsa-miR-504 | hsa-miR-186 |  |  |
| hsa-miR-376c-3p | hsa-miR-499a-5p | hsa-miR-3688 |  |  |
| hsa-miR-4326 | hsa-miR-1250 | hsa-miR-6779 |  |  |
| hsa-miR-671-5p | hsa-miR-136 | hsa-miR-3607 |  |  |
| hsa-miR-3130-5p | hsa-miR-147 | hsa-miR-215 |  |  |
| hsa-miR-92a-1-5p | hsa-miR-220a | hsa-miR-873 |  |  |
| hsa-miR-223-5p | hsa-miR-27b | hsa-miR-6778 |  |  |
| hsa-miR-1293 | hsa-miR-323-5p | hsa-miR-125a |  |  |
| hsa-miR-425-3p | hsa-miR-503 | hsa-miR-134 |  |  |
| hsa-miR-495-3p | hsa-miR-632 | hsa-miR-4282 |  |  |
| hsa-miR-335-5p | hsa-miR-646 | hsa-miR-34b |  |  |
| hsa-miR-3614-5p | hsa-miR-668 | hsa-miR-4525 |  |  |
| hsa-miR-185-3p | hsa-miR-877 | hsa-miR-4795 |  |  |
| hsa-miR-29b-1-5p | hsa-miR-300 | hsa-miR-32 |  |  |
| hsa-miR-320b | hsa-miR-451a | hsa-miR-136 |  |  |
| hsa-miR-342-5p | hsa-miR-208 | hsa-miR-1193 |  |  |
| hsa-miR-369-3p | hsa-miR-338 | hsa-miR-135b |  |  |
| hsa-miR-214-3p | hsa-miR-183 | hsa-miR-484 |  |  |
| hsa-miR-3615 | hsa-miR-335 | hsa-miR-571 |  |  |
| hsa-miR-221-5p | hsa-miR-184 | hsa-miR-204 |  |  |
| hsa-miR-204-5p | hsa-miR-329 | hsa-miR-628 |  |  |
| hsa-miR-95-3p | hsa-miR-410 | hsa-miR-520d |  |  |
| hsa-miR-23a-5p | hsa-miR-101 | hsa-miR-4662a |  |  |
| hsa-miR-432-5p | hsa-miR-32 | hsa-miR-7151 |  |  |
| hsa-miR-212-3p | hsa-miR-494 | hsa-miR-29a |  |  |
| hsa-miR-502-3p | hsa-miR-196a | hsa-miR-106b |  |  |
| hsa-miR-409-5p | hsa-miR-130b | hsa-miR-146a |  |  |
| hsa-miR-20a-3p | hsa-miR-429 | hsa-miR-7110 |  |  |
| hsa-miR-1301-3p | hsa-miR-26a | hsa-miR-4429 |  |  |
| hsa-miR-766-3p | hsa-miR-26b | hsa-miR-98 |  |  |
| hsa-miR-411-5p | hsa-miR-372 | hsa-miR-19a |  |  |
| hsa-let-7e-3p | hsa-miR-1179 | hsa-miR-558 |  |  |
| hsa-miR-431-3p | hsa-miR-128 | hsa-miR-1273f |  |  |
| hsa-miR-2355-3p | hsa-miR-1290 | hsa-miR-384 |  |  |
| hsa-miR-454-3p | hsa-miR-942 | hsa-miR-34c |  |  |
| hsa-miR-493-5p | hsa-miR-29b | hsa-miR-570 |  |  |
| hsa-miR-10b-3p | hsa-miR-1294 | hsa-miR-586 |  |  |
| hsa-miR-136-3p | hsa-miR-506 | hsa-miR-3613 |  |  |
| hsa-miR-450a-5p | hsa-miR-216a | hsa-miR-34a |  |  |
| hsa-miR-362-5p | hsa-miR-433 | hsa-miR-6504 |  |  |
| hsa-miR-378c | hsa-miR-1271 | hsa-miR-3149 |  |  |
| hsa-miR-503-5p | hsa-miR-330-3p | hsa-miR-548aw |  |  |
| hsa-miR-628-5p | hsa-miR-483-5p | hsa-miR-4637 |  |  |
| hsa-miR-129-5p | hsa-miR-224 | hsa-miR-526b |  |  |
| hsa-miR-301a-3p | hsa-miR-99b-3p | hsa-miR-103a |  |  |
| hsa-miR-19b-1-5p | hsa-miR-340 | hsa-miR-328 |  |  |
| hsa-miR-1976 | hsa-miR-192 | hsa-miR-5682 |  |  |
| hsa-miR-339-3p | hsa-miR-188 | hsa-miR-379 |  |  |
| hsa-miR-942-5p | hsa-miR-613 | hsa-miR-4778 |  |  |
| hsa-miR-497-5p | hsa-miR-92b | hsa-miR-4799 |  |  |
| hsa-miR-452-3p | hsa-miR-451 | hsa-miR-130b |  |  |
| hsa-miR-758-3p | hsa-miR-101-3p | hsa-miR-449a |  |  |
| hsa-miR-184 | hsa-miR-186 | hsa-miR-4685 |  |  |
| hsa-miR-193a-3p | hsa-miR-27a-3p | hsa-miR-5001 |  |  |
| hsa-miR-139-3p | hsa-miR-301a-3p | hsa-miR-324 |  |  |
| hsa-miR-29b-2-5p | hsa-miR-425 | hsa-miR-3942 |  |  |
| hsa-miR-93-3p | hsa-miR-140-5p | hsa-miR-1287 |  |  |
| hsa-miR-29c-5p | hsa-miR-718 | hsa-miR-199b |  |  |
| hsa-miR-576-5p | hsa-miR-483-3p | hsa-miR-149 |  |  |
| hsa-miR-410-3p | hsa-miR-365a-3p | hsa-miR-4276 |  |  |
| hsa-miR-181d-5p | hsa-miR-144 | hsa-miR-92b |  |  |
| hsa-miR-217 | hsa-miR-216b | hsa-miR-6891 |  |  |
| hsa-miR-345-5p | hsa-miR-202 | hsa-miR-4507 |  |  |
| hsa-miR-193b-5p | hsa-let-7f | hsa-miR-6871 |  |  |
| hsa-miR-3613-5p | hsa-miR-142-3p | hsa-miR-6833 |  |  |
| hsa-miR-1247-5p | hsa-miR-422a | hsa-miR-3659 |  |  |
| hsa-miR-511-5p | hsa-miR-153 | hsa-miR-4747 |  |  |
| hsa-miR-450b-5p | hsa-miR-495 | hsa-miR-363 |  |  |
| hsa-miR-590-5p | hsa-miR-187* | hsa-miR-6857 |  |  |
| hsa-miR-144-3p | hsa-miR-143-3p | hsa-miR-494 |  |  |
| hsa-miR-582-5p | hsa-miR-644a | hsa-miR-4694 |  |  |
| hsa-miR-16-2-3p | hsa-miR-194 | hsa-miR-5192 |  |  |
| hsa-miR-340-5p | hsa-miR-622 | hsa-miR-4755 |  |  |
| hsa-miR-130b-5p | hsa-miR-497 | hsa-miR-376c |  |  |
| hsa-miR-363-3p | hsa-miR-187 | hsa-miR-330 |  |  |
| hsa-miR-328-3p | hsa-miR-630 | hsa-miR-4803 |  |  |
| hsa-miR-34c-3p | hsa-miR-588 | hsa-miR-3921 |  |  |
| hsa-miR-1287-5p | hsa-miR-181d | hsa-miR-4458 |  |  |
| hsa-miR-26b-3p | hsa-miR-23b | hsa-miR-877 |  |  |
| hsa-miR-889-3p | hsa-miR-33b | hsa-miR-764 |  |  |
| hsa-miR-382-5p | hsa-miR-424-5p | hsa-miR-769 |  |  |
| hsa-miR-744-3p | hsa-miR-141-3p | hsa-miR-3065 |  |  |
| hsa-miR-24-1-5p | hsa-miR-127 | hsa-miR-6842 |  |  |
| hsa-miR-30c-2-3p | hsa-miR-1288 | hsa-miR-8062 |  |  |
| hsa-miR-598-3p | hsa-miR-182 | hsa-miR-548aj |  |  |
| hsa-miR-664a-3p | hsa-miR-486-5p | hsa-miR-148b |  |  |
| hsa-miR-222-5p | hsa-miR-675 | hsa-miR-548j |  |  |
| hsa-miR-769-5p | hsa-miR-192-5p | hsa-miR-6847 |  |  |
| hsa-miR-7-1-3p | hsa-miR-28-3p | hsa-miR-6751 |  |  |
| hsa-miR-874-3p | hsa-miR-20b-5p | hsa-miR-664b |  |  |
| hsa-miR-214-5p | hsa-miR-296-5p | hsa-miR-6738 |  |  |
| hsa-miR-96-5p | hsa-miR-223-3p | hsa-miR-18b |  |  |
| hsa-miR-331-3p | hsa-miR-193a-3p | hsa-miR-4668 |  |  |
| hsa-miR-409-3p | hsa-miR-638 | hsa-miR-181a |  |  |
| hsa-miR-145-3p | hsa-miR-200b-3p | hsa-miR-1912 |  |  |
| hsa-miR-106a-5p | hsa-miR-498 | hsa-miR-492 |  |  |
| hsa-miR-195-5p | hsa-miR-377 | hsa-miR-5571 |  |  |
| hsa-miR-337-3p | hsa-miR-542-3p | hsa-miR-3125 |  |  |
| hsa-miR-136-5p | hsa-miR-21-3p | hsa-miR-4305 |  |  |
| hsa-miR-1180-3p | hsa-miR-448 | hsa-miR-3609 |  |  |
| hsa-let-7g-3p | hsa-miR-584 | hsa-miR-145 |  |  |
| hsa-miR-130b-3p | hsa-miR-125a | hsa-miR-190a |  |  |
| hsa-miR-324-3p | hsa-miR-99b | hsa-miR-425 |  |  |
| hsa-miR-34c-5p | hsa-miR-138-5p | hsa-miR-124 |  |  |
| hsa-miR-324-5p | hsa-miR-372-3p | hsa-miR-6840 |  |  |
| hsa-miR-125b-2-3p | hsa-miR-381 | hsa-miR-138 |  |  |
| hsa-miR-654-3p | hsa-miR-297 | hsa-miR-615 |  |  |
| hsa-miR-18a-5p | hsa-miR-455-3p | hsa-miR-5692c |  |  |
| hsa-miR-652-3p | hsa-miR-150-5p | hsa-miR-365b |  |  |
| hsa-miR-509-3p | hsa-miR-150-3p | hsa-miR-369 |  |  |
| hsa-miR-200b-5p | hsa-miR-432-3p | hsa-miR-92a |  |  |
| hsa-miR-532-3p | hsa-miR-139-5p | hsa-miR-122 |  |  |
| hsa-miR-744-5p | hsa-miR-145-5p | hsa-miR-29c |  |  |
| hsa-miR-20b-5p | hsa-miR-125b-5p | hsa-miR-577 |  |  |
| hsa-miR-3607-3p | hsa-miR-191 | hsa-miR-1976 |  |  |
| hsa-miR-32-5p | hsa-miR-21-5p | hsa-miR-383 |  |  |
| hsa-miR-181c-5p | hsa-miR-548k | hsa-miR-3163 |  |  |
| hsa-let-7b-3p | hsa-miR-129 | hsa-miR-4719 |  |  |
| hsa-miR-140-5p | hsa-miR-195-5p | hsa-miR-6883 |  |  |
| hsa-miR-374a-5p | hsa-miR-30c | hsa-miR-6867 |  |  |
| hsa-miR-181c-3p | hsa-miR-1470 | hsa-miR-137 |  |  |
| hsa-miR-218-5p | hsa-miR-204 | hsa-miR-4476 |  |  |
| hsa-miR-143-5p | hsa-miR-137 | hsa-miR-885 |  |  |
| hsa-miR-514a-3p | hsa-miR-371 | hsa-miR-7109 |  |  |
| hsa-miR-505-3p | hsa-miR-4497 | hsa-miR-449b |  |  |
| hsa-miR-19a-3p | hsa-miR-125a-5p | hsa-miR-548s |  |  |
| hsa-miR-330-5p | hsa-miR-765 | hsa-miR-23c |  |  |
| hsa-miR-33a-5p | hsa-miR-135b | hsa-miR-7a |  |  |
| hsa-miR-501-3p | hsa-miR-1297 | hsa-miR-8084 |  |  |
| hsa-miR-139-5p | hsa-miR-516b | hsa-miR-18a |  |  |
| hsa-miR-22-5p | hsa-miR-876-5p | hsa-miR-624 |  |  |
| hsa-miR-339-5p | hsa-miR-7 | hsa-miR-7641 |  |  |
| hsa-let-7a-3p | hsa-miR-365 | hsa-miR-6129 |  |  |
| hsa-miR-148a-5p | hsa-miR-182-5p | hsa-miR-5580 |  |  |
| hsa-miR-3065-3p | hsa-miR-211 | hsa-miR-24 |  |  |
| hsa-miR-660-5p | hsa-miR-6775-3p | hsa-miR-6764 |  |  |
| hsa-miR-27b-5p | hsa-miR-424 | hsa-miR-126 |  |  |
| hsa-miR-187-3p | hsa-miR-10a | hsa-miR-101 |  |  |
| hsa-miR-767-5p | hsa-miR-10b-3p | hsa-miR-6864 |  |  |
| hsa-miR-15b-3p | hsa-miR-455-5p | hsa-miR-218 |  |  |
| hsa-miR-374b-5p | hsa-miR-384 | hsa-miR-7977 |  |  |
| hsa-miR-135b-5p | hsa-miR-625 | hsa-miR-548n |  |  |
| hsa-miR-24-2-5p | hsa-miR-338-5p | hsa-miR-583 |  |  |
| hsa-miR-98-5p | hsa-miR-141 | hsa-miR-718 |  |  |
| hsa-miR-423-5p | hsa-miR-23a-3p | hsa-miR-593 |  |  |
| hsa-miR-99b-3p | hsa-miR-342-3p | hsa-miR-548a |  |  |
| hsa-miR-27a-5p | hsa-miR-610 | hsa-miR-6510 |  |  |
| hsa-miR-200c-5p | hsa-miR-106a* | hsa-miR-4306 |  |  |
| hsa-miR-455-5p | hsa-miR-625-3p | hsa-miR-221-3p |  |  |
| hsa-miR-574-3p | hsa-miR-1258 | hsa-miR-222 |  |  |
| hsa-miR-133b | hsa-miR-219-5p | hsa-miR-548d |  |  |
| hsa-miR-508-3p | hsa-miR-601 | hsa-miR-6874 |  |  |
| hsa-miR-589-5p | hsa-miR-548d-3p | hsa-miR-4445 |  |  |
| hsa-miR-31-3p | hsa-miR-29a-3p | hsa-miR-5586 |  |  |
| hsa-miR-365a-3p | hsa-miR-373-3p | hsa-miR-375 |  |  |
| hsa-miR-105-5p | hsa-miR-372-5p | hsa-miR-4279 |  |  |
| hsa-miR-381-3p | hsa-miR-27b-5p | hsa-miR-3606 |  |  |
| hsa-miR-335-3p | hsa-miR-450a | hsa-miR-1909 |  |  |
| hsa-miR-107 | hsa-miR-96-5p | hsa-miR-744 |  |  |
| hsa-miR-484 | hsa-miR-221-3p | hsa-miR-519c |  |  |
| hsa-miR-675-3p | hsa-miR-200a | hsa-miR-329 |  |  |
| hsa-miR-429 |  | hsa-miR-548y |  |  |
| hsa-miR-132-3p |  | hsa-miR-133a-3p |  |  |
| hsa-miR-423-3p |  | hsa-miR-20a |  |  |
| hsa-miR-424-5p |  | hsa-miR-208b |  |  |
| hsa-miR-222-3p |  | hsa-miR-15b |  |  |
| hsa-let-7i-3p |  | hsa-miR-152 |  |  |
| hsa-miR-1269a |  | hsa-miR-197 |  |  |
| hsa-miR-31-5p |  | hsa-miR-1468 |  |  |
| hsa-miR-34a-5p |  | hsa-miR-200c |  |  |
| hsa-miR-142-5p |  | hsa-miR-3124 |  |  |
| hsa-miR-194-5p |  | hsa-miR-133b |  |  |
| hsa-miR-629-5p |  | hsa-miR-4487 |  |  |
| hsa-miR-378a-5p |  | hsa-miR-4715 |  |  |
| hsa-miR-144-5p |  | hsa-miR-6832 |  |  |
| hsa-miR-126-5p |  | hsa-miR-6817 |  |  |
| hsa-miR-224-5p |  | hsa-miR-6715b |  |  |
| hsa-miR-199b-5p |  | hsa-miR-4733 |  |  |
| hsa-miR-19b-3p |  | hsa-miR-4632 |  |  |
| hsa-miR-127-5p |  | hsa-miR-6757 |  |  |
| hsa-miR-149-5p |  | hsa-miR-1234 |  |  |
| hsa-miR-193b-3p |  | hsa-miR-338 |  |  |
| hsa-miR-361-3p |  | hsa-miR-3652 |  |  |
| hsa-miR-128-3p |  | hsa-miR-4763 |  |  |
| hsa-miR-584-5p |  | hsa-miR-515 |  |  |
| hsa-miR-342-3p |  | hsa-miR-3662 |  |  |
| hsa-miR-15a-5p |  | hsa-miR-4292 |  |  |
| hsa-miR-625-3p |  | hsa-miR-193b |  |  |
| hsa-miR-486-5p |  | hsa-miR-184 |  |  |
| hsa-miR-146a-5p |  | hsa-miR-2115 |  |  |
| hsa-let-7d-5p |  | hsa-miR-6785 |  |  |
| hsa-miR-181a-3p |  | hsa-miR-524 |  |  |
| hsa-miR-196b-5p |  | hsa-miR-607 |  |  |
| hsa-miR-186-5p |  | hsa-miR-192 |  |  |
| hsa-miR-944 |  | hsa-miR-4660 |  |  |
| hsa-miR-361-5p |  | hsa-miR-365a |  |  |
| hsa-miR-542-3p |  | hsa-miR-7703 |  |  |
| hsa-miR-500a-3p |  | hsa-miR-548au |  |  |
| hsa-miR-452-5p |  | hsa-miR-7i |  |  |
| hsa-miR-708-3p |  | hsa-miR-302c |  |  |
| hsa-miR-200a-3p |  | hsa-miR-4703 |  |  |
| hsa-miR-193a-5p |  | hsa-miR-3934 |  |  |
| hsa-miR-425-5p |  | hsa-miR-6165 |  |  |
| hsa-let-7d-3p |  | hsa-miR-483 |  |  |
| hsa-miR-30b-5p |  | hsa-miR-30b |  |  |
| hsa-miR-15b-5p |  | hsa-miR-1908 |  |  |
| hsa-miR-197-3p |  | hsa-miR-335 |  |  |
| hsa-miR-133a |  | hsa-miR-125b |  |  |
| hsa-miR-181b-5p |  | hsa-miR-6859 |  |  |
| hsa-miR-192-5p |  | hsa-miR-147a |  |  |
| hsa-miR-125a-5p |  | hsa-miR-508 |  |  |
| hsa-miR-106b-5p |  | hsa-miR-1178 |  |  |
| hsa-miR-582-3p |  | hsa-miR-6845 |  |  |
| hsa-miR-134-5p |  | hsa-miR-4650 |  |  |
| hsa-miR-1 |  | hsa-miR-4524a |  |  |
| hsa-miR-338-3p |  | hsa-miR-7108 |  |  |
| hsa-miR-191-5p |  | hsa-miR-5703 |  |  |
| hsa-miR-30c-5p |  | hsa-miR-107 |  |  |
| hsa-miR-17-3p |  | hsa-miR-548z |  |  |
| hsa-miR-152-3p |  | hsa-miR-3680 |  |  |
| hsa-miR-221-3p |  | hsa-miR-421 |  |  |
| hsa-miR-455-3p |  | hsa-miR-6735 |  |  |
| hsa-miR-99a-5p |  | hsa-miR-598 |  |  |
| hsa-miR-106b-3p |  | hsa-miR-4261 |  |  |
| hsa-miR-125b-5p |  | hsa-miR-3690 |  |  |
| hsa-miR-20a-5p |  | hsa-miR-301a |  |  |
| hsa-let-7g-5p |  | hsa-miR-141 |  |  |
| hsa-let-7i-5p |  | hsa-miR-4797 |  |  |
| hsa-miR-29b-3p |  | hsa-miR-4696 |  |  |
| hsa-miR-127-3p |  | hsa-miR-19b |  |  |
| hsa-miR-26b-5p |  | hsa-miR-5187 |  |  |
| hsa-miR-16-5p |  | hsa-miR-374a |  |  |
| hsa-miR-17-5p |  | hsa-miR-4506 |  |  |
| hsa-miR-223-3p |  | hsa-miR-3161 |  |  |
| hsa-miR-532-5p |  | hsa-miR-7 |  |  |
| hsa-miR-320a |  | hsa-miR-4272 |  |  |
| hsa-miR-199a-5p |  | hsa-miR-708 |  |  |
| hsa-miR-155-5p |  | hsa-miR-4690 |  |  |
| hsa-miR-200b-3p |  | hsa-miR-891a |  |  |
| hsa-miR-146b-5p |  | hsa-miR-374b |  |  |
| hsa-miR-378a-3p |  | hsa-miR-502 |  |  |
| hsa-miR-200a-5p |  | hsa-miR-4728 |  |  |
| hsa-miR-145-5p |  | hsa-miR-6761 |  |  |
| hsa-miR-140-3p |  | hsa-miR-199a |  |  |
| hsa-miR-451a |  | hsa-miR-30c |  |  |
| hsa-miR-141-5p |  | hsa-miR-4790 |  |  |
| hsa-miR-374a-3p |  | hsa-miR-548g |  |  |
| hsa-miR-141-3p |  | hsa-miR-500a |  |  |
| hsa-miR-181a-2-3p |  | hsa-miR-548f |  |  |
| hsa-let-7e-5p |  | hsa-miR-455 |  |  |
| hsa-miR-379-5p |  | hsa-miR-25 |  |  |
| hsa-miR-150-5p |  | hsa-miR-4633 |  |  |
| hsa-miR-181a-5p |  | hsa-miR-653 |  |  |
| hsa-miR-210-3p |  | hsa-miR-519e |  |  |
| hsa-miR-375 |  | hsa-miR-320a |  |  |
| hsa-miR-23b-3p |  | hsa-miR-3176 |  |  |
| hsa-miR-1307-3p |  | hsa-miR-4432 |  |  |
| hsa-miR-206 |  | hsa-miR-4254 |  |  |
| hsa-miR-29c-3p |  | hsa-miR-3141 |  |  |
| hsa-let-7c-5p |  | hsa-miR-151a |  |  |
| hsa-miR-26a-5p |  | hsa-miR-423 |  |  |
| hsa-miR-30a-3p |  | hsa-miR-182 |  |  |
| hsa-miR-126-3p |  | hsa-miR-371a |  |  |
| hsa-miR-151a-3p |  | hsa-miR-6741 |  |  |
| hsa-miR-199a-3p |  | hsa-miR-30d |  |  |
| hsa-miR-199b-3p |  | hsa-miR-5688 |  |  |
| hsa-miR-27b-3p |  | hsa-miR-6127 |  |  |
| hsa-miR-100-5p |  | hsa-miR-587 |  |  |
| hsa-miR-142-3p |  | hsa-miR-4742 |  |  |
| hsa-miR-27a-3p |  | hsa-miR-4676 |  |  |
| hsa-miR-28-3p |  | hsa-miR-3919 |  |  |
| hsa-miR-24-3p |  | hsa-miR-6740 |  |  |
| hsa-miR-10a-5p |  | hsa-miR-6759 |  |  |
| hsa-miR-30e-3p |  | hsa-miR-424 |  |  |
| hsa-miR-30e-5p |  | hsa-miR-142 |  |  |
| hsa-miR-30d-5p |  | hsa-miR-548h |  |  |
| hsa-miR-23a-3p |  | hsa-miR-4490 |  |  |
| hsa-miR-9-5p |  | hsa-miR-548an |  |  |
| hsa-miR-21-3p |  | hsa-miR-26a |  |  |
| hsa-miR-29a-3p |  | hsa-miR-31 |  |  |
| hsa-miR-93-5p |  | hsa-miR-342 |  |  |
| hsa-miR-30a-5p |  | hsa-miR-449c |  |  |
| hsa-miR-101-3p |  | hsa-miR-6512 |  |  |
| hsa-miR-183-5p |  | hsa-miR-1910 |  |  |
| hsa-let-7f-5p |  | hsa-miR-6511a |  |  |
| hsa-miR-25-3p |  | hsa-miR-4643 |  |  |
| hsa-miR-200c-3p |  | hsa-miR-550a |  |  |
| hsa-miR-92a-3p |  | hsa-miR-4482 |  |  |
| hsa-miR-99b-5p |  | hsa-miR-4500 |  |  |
| hsa-miR-205-5p |  | hsa-miR-485 |  |  |
| hsa-let-7b-5p |  | hsa-miR-3926 |  |  |
| hsa-miR-182-5p |  | hsa-miR-6824 |  |  |
| hsa-miR-103a-3p |  | hsa-miR-3941 |  |  |
| hsa-miR-10b-5p |  | hsa-miR-4531 |  |  |
| hsa-miR-148a-3p |  | hsa-miR-3127 |  |  |
| hsa-let-7a-5p |  | hsa-miR-4743 |  |  |
| hsa-miR-143-3p |  | hsa-miR-548k |  |  |
| hsa-miR-22-3p |  | hsa-miR-4680 |  |  |
| hsa-miR-203a |  | hsa-miR-937 |  |  |
| hsa-miR-21-5p |  | hsa-miR-658 |  |  |
|  |  | hsa-miR-1915 |  |  |
|  |  | hsa-miR-4491 |  |  |
|  |  | hsa-miR-302b |  |  |
|  |  | hsa-miR-2276 |  |  |
|  |  | hsa-miR-3173 |  |  |
|  |  | hsa-miR-378h |  |  |
|  |  | hsa-miR-6873 |  |  |
|  |  | hsa-miR-296 |  |  |
|  |  | hsa-miR-6860 |  |  |
|  |  | hsa-miR-493 |  |  |
|  |  | hsa-miR-6838 |  |  |
|  |  |  |  | hsa-miR-5582 |
|  |  |  |  | hsa-miR-6760 |
|  |  |  |  | hsa-miR-200a |
|  |  |  |  | hsa-miR-4661 |
|  |  |  |  | hsa-miR-6077 |
|  |  |  |  | hsa-miR-211 |
|  |  |  |  | hsa-miR-556 |
|  |  |  |  | hsa-miR-3667 |
|  |  |  |  | hsa-miR-548ab |
|  |  |  |  | hsa-miR-525 |
|  |  |  |  | hsa-miR-5088 |
|  |  |  |  | hsa-miR-4689 |
|  |  |  |  | hsa-miR-640 |
|  |  |  |  | hsa-miR-6792 |
|  |  |  |  | hsa-miR-940 |
|  |  |  |  | hsa-miR-216a |
|  |  |  |  | hsa-miR-33a |
|  |  |  |  | hsa-miR-766 |
|  |  |  |  | hsa-miR-6791 |
|  |  |  |  | hsa-miR-1303 |
|  |  |  |  | hsa-miR-4714 |
|  |  |  |  | hsa-miR-7d |
|  |  |  |  | hsa-miR-6880 |
|  |  |  |  | hsa-miR-3689a |
|  |  |  |  | hsa-miR-6813 |
|  |  |  |  | hsa-miR-6878 |
|  |  |  |  | hsa-miR-448 |
|  |  |  |  | hsa-miR-4465 |
|  |  |  |  | hsa-miR-6793 |
|  |  |  |  | hsa-miR-200b |
|  |  |  |  | hsa-miR-491 |
|  |  |  |  | hsa-miR-3133 |
|  |  |  |  | hsa-miR-6780a |
|  |  |  |  | hsa-miR-4705 |
|  |  |  |  | hsa-miR-761 |
|  |  |  |  | hsa-miR-1233 |
|  |  |  |  | hsa-miR-4659b |
|  |  |  |  | hsa-miR-6514 |
|  |  |  |  | hsa-miR-2682 |
|  |  |  |  | hsa-miR-4473 |
|  |  |  |  | hsa-miR-8069 |
|  |  |  |  | hsa-miR-219a |
|  |  |  |  | hsa-miR-4524b |
|  |  |  |  | hsa-miR-409 |
|  |  |  |  | hsa-miR-3129 |
|  |  |  |  | hsa-miR-429 |
|  |  |  |  | hsa-miR-144 |
|  |  |  |  | hsa-miR-320d |
|  |  |  |  | hsa-miR-3160 |
|  |  |  |  | hsa-miR-3685 |
|  |  |  |  | hsa-miR-548ba |
|  |  |  |  | hsa-miR-4775 |
|  |  |  |  | hsa-miR-3143 |
|  |  |  |  | hsa-miR-4303 |
|  |  |  |  | hsa-miR-3973 |
|  |  |  |  | hsa-miR-544a |
|  |  |  |  | hsa-miR-4463 |
|  |  |  |  | hsa-miR-7113 |
|  |  |  |  | hsa-miR-4727 |

**Table S3** Cluster analyses Results to obtain coclustered miRs with their sequence similar to the miRs obtained from cancer specific database search

| Cluster Analysis Result | |
| --- | --- |
| Selected miRNAs belong/related to: head and neck cancer, head and neck carcinoma, head and neck squamous cell carcinoma, oral cancer, oral carcinoma, oral squamous cell carcinoma, oropharyngeal cancer, oropharyngeal carcinoma, | |
| Analysis configurations: |  |
| Only look at a section (from 1 to end) of miRNA sequences, and sequences with total length longer than 0 shorter than 25. | |
| Expression is both. |  |
| Clustering scores: gap=1 substitution=1 distance= 6 | |
| ------All------(Total 108 clusters) | |
| Clusters with 1 sequences--(67 clusters)-- | |
| Cluster of size 1 (#1) |  |
| hsa-miR-1271 | CUUGGCACCUAGCAAGCACUCA |
| Related to 17 cancers |  |
| Cluster of size 1 (#2) |  |
| hsa-miR-220a | CCACACCGUAUCUGACACUUU |
| Related to 1 cancers |  |
| Cluster of size 1 (#3) |  |
| hsa-miR-137 | UUAUUGCUUAAGAAUACGCGUAG |
| Related to 58 cancers |  |
| Cluster of size 1 (#4) |  |
| hsa-miR-345 | GCUGACUCCUAGUCCAGGGCUC |
| Related to 10 cancers |  |
| Cluster of size 1 (#5) |  |
| hsa-miR-188 | CAUCCCUUGCAUGGUGGAGGGU |
| Related to 7 cancers |  |
| Cluster of size 1 (#6) |  |
| hsa-miR-365 | UAAUGCCCCUAAAAAUCCUUAU |
| Related to 25 cancers |  |
| Cluster of size 1 (#7) |  |
| hsa-miR-877 | GUAGAGGAGAUGGCGCAGGG |
| Related to 4 cancers |  |
| Cluster of size 1 (#8) |  |
| hsa-miR-124 | UAAGGCACGCGGUGAAUGCC |
| Related to 129 cancers |  |
| Cluster of size 1 (#9) |  |
| hsa-miR-876-5p | UGGAUUUCUUUGUGAAUCACCA |
| Related to 2 cancers |  |
| Cluster of size 1 (#10) |  |
| hsa-miR-1250 | ACGGUGCUGGAUGUGGCCUUU |
| Related to 1 cancers |  |
| Cluster of size 1 (#11) |  |
| hsa-miR-216a | UAAUCUCAGCUGGCAACUGUGA |
| Related to 21 cancers |  |
| Cluster of size 1 (#12) |  |
| hsa-miR-186 | CAAAGAAUUCUCCUUUUGGGCU |
| Related to 23 cancers |  |
| Cluster of size 1 (#13) |  |
| hsa-miR-187 | UCGUGUCUUGUGUUGCAGCCGG |
| Related to 15 cancers |  |
| Cluster of size 1 (#14) |  |
| hsa-miR-497 | CAGCAGCACACUGUGGUUUGU |
| Related to 51 cancers |  |
| Cluster of size 1 (#15) |  |
| hsa-miR-874 | CUGCCCUGGCCCGAGGGACCGA |
| Related to 13 cancers |  |
| Cluster of size 1 (#16) |  |
| hsa-miR-632 | GUGUCUGCUUCCUGUGGGA |
| Related to 3 cancers |  |
| Cluster of size 1 (#17) |  |
| hsa-miR-422a | ACUGGACUUAGGGUCAGAAGGC |
| Related to 11 cancers |  |
| Cluster of size 1 (#18) |  |
| hsa-miR-410 | AAUAUAACACAGAUGGCCUGU |
| Related to 10 cancers |  |
| Cluster of size 1 (#19) |  |
| hsa-miR-30a-5p | UGUAAACAUCCUCGACUGGAAG |
| Related to 19 cancers |  |
| Cluster of size 1 (#20) |  |
| hsa-miR-99b-3p | CAAGCUCGUGUCUGUGGGUCCG |
| Related to 2 cancers |  |
| Cluster of size 1 (#21) |  |
| hsa-miR-494 | UGAAACAUACACGGGAAACCUC |
| Related to 32 cancers |  |
| Cluster of size 1 (#22) |  |
| hsa-miR-338 | UCCAGCAUCAGUGAUUUUGUUGA |
| Related to 42 cancers |  |
| Cluster of size 1 (#23) |  |
| hsa-miR-675 | UGGUGCGGAGAGGGCCCACAGUG |
| Related to 13 cancers |  |
| Cluster of size 1 (#24) |  |
| hsa-miR-451 | AAACCGUUACCAUUACUGAGUU |
| Related to 45 cancers |  |
| Cluster of size 1 (#25) |  |
| hsa-miR-646 | AAGCAGCUGCCUCUGAGGC |
| Related to 7 cancers |  |
| Cluster of size 1 (#26) |  |
| hsa-miR-1258 | AGUUAGGAUUAGGUCGUGGAA |
| Related to 7 cancers |  |
| Cluster of size 1 (#27) |  |
| hsa-miR-370 | GCCUGCUGGGGUGGAACCUGGU |
| Related to 21 cancers |  |
| Cluster of size 1 (#28) |  |
| hsa-miR-147 | GUGUGUGGAAAUGCUUCUGC |
| Related to 11 cancers |  |
| Cluster of size 1 (#29) |  |
| hsa-miR-149 | UCUGGCUCCGUGUCUUCACUCCC |
| Related to 19 cancers |  |
| Cluster of size 1 (#30) |  |
| hsa-miR-144 | UACAGUAUAGAUGAUGUACU |
| Related to 45 cancers |  |
| Cluster of size 1 (#31) |  |
| hsa-miR-22 | AAGCUGCCAGUUGAAGAACUGU |
| Related to 322 cancers |  |
| Cluster of size 1 (#32) |  |
| hsa-miR-184 | UGGACGGAGAACUGAUAAGGGU |
| Related to 17 cancers |  |
| Cluster of size 1 (#33) |  |
| hsa-miR-450a | UUUUGCGAUGUGUUCCUAAUAU |
| Related to 2 cancers |  |
| Cluster of size 1 (#34) |  |
| hsa-miR-187* | GGCUACAACACAGGACCCGGGC |
| Related to 1 cancers |  |
| Cluster of size 1 (#35) |  |
| hsa-miR-483-5p | AAGACGGGAGGAAAGAAGGGAG |
| Related to 12 cancers |  |
| Cluster of size 1 (#36) |  |
| hsa-miR-139-5p | UCUACAGUGCACGUGUCUCCAG |
| Related to 25 cancers |  |
| Cluster of size 1 (#37) |  |
| hsa-miR-323-5p | AGGUGGUCCGUGGCGCGUUCGC |
| Related to 1 cancers |  |
| Cluster of size 1 (#38) |  |
| hsa-miR-139-3p | GGAGACGCGGCCCUGUUGGAGU |
| Related to 6 cancers |  |
| Cluster of size 1 (#39) |  |
| hsa-miR-499a-5p | UUAAGACUUGCAGUGAUGUUU |
| Related to 3 cancers |  |
| Cluster of size 1 (#40) |  |
| hsa-miR-134 | UGUGACUGGUUGACCAGAGGGG |
| Related to 17 cancers |  |
| Cluster of size 1 (#41) |  |
| hsa-miR-377 | AUCACACAAAGGCAACUUUUGU |
| Related to 13 cancers |  |
| Cluster of size 1 (#42) |  |
| hsa-miR-503 | UAGCAGCGGGAACAGUUCUGCAG |
| Related to 32 cancers |  |
| Cluster of size 1 (#43) |  |
| hsa-miR-373 | GAAGUGCUUCGAUUUUGGGGUGU |
| Related to 29 cancers |  |
| Cluster of size 1 (#44) |  |
| hsa-miR-610 | UGAGCUAAAUGUGUGCUGGGA |
| Related to 7 cancers |  |
| Cluster of size 1 (#45) |  |
| hsa-miR-342-3p | UCUCACACAGAAAUCGCACCCGU |
| Related to 8 cancers |  |
| Cluster of size 1 (#46) |  |
| hsa-miR-148a | UCAGUGCACUACAGAACUUUGU |
| Related to 52 cancers |  |
| Cluster of size 1 (#47) |  |
| hsa-miR-136 | ACUCCAUUUGUUUUGAUGAUGGA |
| Related to 14 cancers |  |
| Cluster of size 1 (#48) |  |
| hsa-miR-106a* | CUGCAAUGUAAGCACUUCUUAC |
| Related to 3 cancers |  |
| Cluster of size 1 (#49) |  |
| hsa-miR-548d-3p | CAAAAACCACAGUUUCUUUUGC |
| Related to 1 cancers |  |
| Cluster of size 1 (#50) |  |
| hsa-miR-1294 | UGUGAGGUUGGCAUUGUUGUCU |
| Related to 4 cancers |  |
| Cluster of size 1 (#51) |  |
| hsa-miR-340 | UUAUAAAGCAAUGAGACUGAUU |
| Related to 32 cancers |  |
| Cluster of size 1 (#52) |  |
| hsa-miR-668 | UGUCACUCGGCUCGGCCCACUAC |
| Related to 1 cancers |  |
| Cluster of size 1 (#53) |  |
| hsa-miR-506 | UAAGGCACCCUUCUGAGUAGA |
| Related to 31 cancers |  |
| Cluster of size 1 (#54) |  |
| hsa-miR-128 | UCACAGUGAACCGGUCUCUUU |
| Related to 47 cancers |  |
| Cluster of size 1 (#55) |  |
| hsa-miR-329 | AACACACCUGGUUAACCUCUUU |
| Related to 13 cancers |  |
| Cluster of size 1 (#56) |  |
| hsa-miR-142-3p | UGUAGUGUUUCCUACUUUAUGGA |
| Related to 26 cancers |  |
| Cluster of size 1 (#57) |  |
| hsa-miR-297 | AUGUAUGUGUGCAUGUGCAUG |
| Related to 2 cancers |  |
| Cluster of size 1 (#58) |  |
| hsa-miR-150-3p | CUGGUACAGGCCUGGGGGACAG |
| Related to 2 cancers |  |
| Cluster of size 1 (#59) |  |
| hsa-miR-214 | ACAGCAGGCACAGACAGGCAGU |
| Related to 64 cancers |  |
| Cluster of size 1 (#60) |  |
| hsa-miR-423-5p | UGAGGGGCAGAGAGCGAGACUUU |
| Related to 7 cancers |  |
| Cluster of size 1 (#61) |  |
| hsa-miR-495 | AAACAAACAUGGUGCACUUCUU |
| Related to 25 cancers |  |
| Cluster of size 1 (#62) |  |
| hsa-miR-150-5p | UCUCCCAACCCUUGUACCAGUG |
| Related to 13 cancers |  |
| Cluster of size 1 (#63) |  |
| hsa-miR-625-3p | GACUAUAGAACUUUCCCCCUCA |
| Related to 4 cancers |  |
| Cluster of size 1 (#64) |  |
| hsa-miR-372-5p | CCUCAAAUGUGGAGCACUAUUCU |
| Related to 1 cancers |  |
| Cluster of size 1 (#65) |  |
| hsa-miR-650 | AGGAGGCAGCGCUCUCAGGAC |
| Related to 7 cancers |  |
| Cluster of size 1 (#66) |  |
| hsa-miR-96-5p | UUUGGCACUAGCACAUUUUUGCU |
| Related to 4 cancers |  |
| Cluster of size 1 (#67) |  |
| hsa-miR-32 | UAUUGCACAUUACUAAGUUGCA |
| Related to 120 cancers |  |
| Clusters with 2 sequences--(15 clusters)-- | |
| Cluster of size 2 (#1) |  |
| hsa-miR-23a-3p | AUCACAUUGCCAGGGAUUUCC |
| Related to 3 cancers |  |
| hsa-miR-23b | AUCACAUUGCCAGGGAUUACC |
| Related to 25 cancers |  |
| Cluster of size 2 (#2) |  |
| hsa-miR-181a | AACAUUCAACGCUGUCGGUGAGU |
| Related to 61 cancers |  |
| hsa-miR-181b | AACAUUCAUUGCUGUCGGUGGGU |
| Related to 42 cancers |  |
| Cluster of size 2 (#3) |  |
| hsa-miR-363 | AAUUGCACGGUAUCCAUCUGUA |
| Related to 16 cancers |  |
| hsa-miR-363 | AAUUGCACGGUAUCCAUCUGUA |
| Related to 16 cancers |  |
| Cluster of size 2 (#4) |  |
| hsa-miR-205 | UCCUUCAUUCCACCGGAGUCUG |
| Related to 80 cancers |  |
| hsa-miR-205 | UCCUUCAUUCCACCGGAGUCUG |
| Related to 80 cancers |  |
| Cluster of size 2 (#5) |  |
| hsa-miR-542-3p | UGUGACAGAUUGAUAACUGAAA |
| Related to 15 cancers |  |
| hsa-miR-542-3p | UGUGACAGAUUGAUAACUGAAA |
| Related to 15 cancers |  |
| Cluster of size 2 (#6) |  |
| hsa-miR-372 | AAAGUGCUGCGACAUUUGAGCGU |
| Related to 19 cancers |  |
| hsa-miR-372 | AAAGUGCUGCGACAUUUGAGCGU |
| Related to 19 cancers |  |
| Cluster of size 2 (#7) |  |
| hsa-miR-203 | GUGAAAUGUUUAGGACCACUAG |
| Related to 94 cancers |  |
| hsa-miR-203 | GUGAAAUGUUUAGGACCACUAG |
| Related to 94 cancers |  |
| Cluster of size 2 (#8) |  |
| hsa-miR-300 | UAUACAAGGGCAGACUCUCUCU |
| Related to 11 cancers |  |
| hsa-miR-300 | UAUACAAGGGCAGACUCUCUCU |
| Related to 11 cancers |  |
| Cluster of size 2 (#9) |  |
| hsa-miR-27b-5p | AGAGCUUAGCUGAUUGGUGAAC |
| Related to 1 cancers |  |
| hsa-miR-27a* | AGGGCUUAGCUGCUUGUGAGCA |
| Related to 1 cancers |  |
| Cluster of size 2 (#10) |  |
| hsa-miR-107 | AGCAGCAUUGUACAGGGCUAUCA |
| Related to 31 cancers |  |
| hsa-miR-107 | AGCAGCAUUGUACAGGGCUAUCA |
| Related to 31 cancers |  |
| Cluster of size 2 (#11) |  |
| hsa-miR-433 | AUCAUGAUGGGCUCCUCGGUGU |
| Related to 17 cancers |  |
| hsa-miR-433 | AUCAUGAUGGGCUCCUCGGUGU |
| Related to 17 cancers |  |
| Cluster of size 2 (#12) |  |
| hsa-miR-21-3p | CAACACCAGUCGAUGGGCUGU |
| Related to 6 cancers |  |
| hsa-miR-21-3p | CAACACCAGUCGAUGGGCUGU |
| Related to 6 cancers |  |
| Cluster of size 2 (#13) |  |
| hsa-miR-146a | UGAGAACUGAAUUCCAUGGGUU |
| Related to 45 cancers |  |
| hsa-miR-146b | UGAGAACUGAAUUCCAUAGGCU |
| Related to 42 cancers |  |
| Cluster of size 2 (#14) |  |
| hsa-miR-24 | UGGCUCAGUUCAGCAGGAACAG |
| Related to 34 cancers |  |
| hsa-miR-24 | UGGCUCAGUUCAGCAGGAACAG |
| Related to 34 cancers |  |
| Cluster of size 2 (#15) |  |
| hsa-miR-126 | UCGUACCGUGAGUAAUAAUGCG |
| Related to 94 cancers |  |
| hsa-miR-126 | UCGUACCGUGAGUAAUAAUGCG |
| Related to 94 cancers |  |
| Clusters with 3 sequences--(8 clusters)-- | |
| Cluster of size 3 (#1) |  |
| hsa-miR-218 | UUGUGCUUGAUCUAACCAUGU |
| Related to 76 cancers |  |
| hsa-miR-218 | UUGUGCUUGAUCUAACCAUGU |
| Related to 76 cancers |  |
| hsa-miR-218 | UUGUGCUUGAUCUAACCAUGU |
| Related to 76 cancers |  |
| Cluster of size 3 (#2) |  |
| hsa-miR-133a-3p | UUUGGUCCCCUUCAACCAGCUG |
| Related to 5 cancers |  |
| hsa-miR-133a | UUUGGUCCCCUUCAACCAGCUG |
| Related to 52 cancers |  |
| hsa-miR-133a | UUUGGUCCCCUUCAACCAGCUG |
| Related to 52 cancers |  |
| Cluster of size 3 (#3) |  |
| hsa-miR-145 | GUCCAGUUUUCCCAGGAAUCCCU |
| Related to 152 cancers |  |
| hsa-miR-145 | GUCCAGUUUUCCCAGGAAUCCCU |
| Related to 152 cancers |  |
| hsa-miR-145 | GUCCAGUUUUCCCAGGAAUCCCU |
| Related to 152 cancers |  |
| Cluster of size 3 (#4) |  |
| hsa-miR-155 | UUAAUGCUAAUCGUGAUAGGGGU |
| Related to 90 cancers |  |
| hsa-miR-155 | UUAAUGCUAAUCGUGAUAGGGGU |
| Related to 90 cancers |  |
| hsa-miR-155 | UUAAUGCUAAUCGUGAUAGGGGU |
| Related to 90 cancers |  |
| Cluster of size 3 (#5) |  |
| hsa-miR-182-5p | UUUGGCAAUGGUAGAACUCACACU |
| Related to 10 cancers |  |
| hsa-miR-182-5p | UUUGGCAAUGGUAGAACUCACACU |
| Related to 10 cancers |  |
| hsa-miR-182 | UUUGGCAAUGGUAGAACUCACACU |
| Related to 74 cancers |  |
| Cluster of size 3 (#6) |  |
| hsa-miR-25 | CAUUGCACUUGUCUCGGUCUGA |
| Related to 38 cancers |  |
| hsa-miR-92a | UAUUGCACUUGUCCCGGCCUGU |
| Related to 46 cancers |  |
| hsa-miR-92b | UAUUGCACUCGUCCCGGCCUCC |
| Related to 12 cancers |  |
| Cluster of size 3 (#7) |  |
| hsa-miR-9 | UCUUUGGUUAUCUAGCUGUAUGA |
| Related to 336 cancers |  |
| hsa-miR-9 | UCUUUGGUUAUCUAGCUGUAUGA |
| Related to 336 cancers |  |
| hsa-miR-9 | UCUUUGGUUAUCUAGCUGUAUGA |
| Related to 336 cancers |  |
| Cluster of size 3 (#8) |  |
| hsa-miR-26b | UUCAAGUAAUUCAGGAUAGGU |
| Related to 44 cancers |  |
| hsa-miR-26a | UUCAAGUAAUCCAGGAUAGGCU |
| Related to 52 cancers |  |
| hsa-miR-1297 | UUCAAGUAAUUCAGGUG |
| Related to 10 cancers |  |
| Clusters with 4 sequences--(7 clusters)-- | |
| Cluster of size 4 (#1) |  |
| hsa-miR-20a | UAAAGUGCUUAUAGUGCAGGUAG |
| Related to 62 cancers |  |
| hsa-miR-93 | CAAAGUGCUGUUCGUGCAGGUAG |
| Related to 56 cancers |  |
| hsa-miR-93 | CAAAGUGCUGUUCGUGCAGGUAG |
| Related to 56 cancers |  |
| hsa-miR-106b | UAAAGUGCUGACAGUGCAGAU |
| Related to 36 cancers |  |
| Cluster of size 4 (#2) |  |
| hsa-miR-143 | UGAGAUGAAGCACUGUAGCUC |
| Related to 94 cancers |  |
| hsa-miR-143 | UGAGAUGAAGCACUGUAGCUC |
| Related to 94 cancers |  |
| hsa-miR-143 | UGAGAUGAAGCACUGUAGCUC |
| Related to 94 cancers |  |
| hsa-miR-143 | UGAGAUGAAGCACUGUAGCUC |
| Related to 94 cancers |  |
| Cluster of size 4 (#3) |  |
| hsa-miR-211 | UUCCCUUUGUCAUCCUUCGCCU |
| Related to 25 cancers |  |
| hsa-miR-204 | UUCCCUUUGUCAUCCUAUGCCU |
| Related to 58 cancers |  |
| hsa-miR-211 | UUCCCUUUGUCAUCCUUCGCCU |
| Related to 25 cancers |  |
| hsa-miR-204 | UUCCCUUUGUCAUCCUAUGCCU |
| Related to 58 cancers |  |
| Cluster of size 4 (#4) |  |
| hsa-miR-27a-3p | UUCACAGUGGCUAAGUUCCGC |
| Related to 8 cancers |  |
| hsa-miR-27b | UUCACAGUGGCUAAGUUCUGC |
| Related to 26 cancers |  |
| hsa-miR-27b | UUCACAGUGGCUAAGUUCUGC |
| Related to 26 cancers |  |
| hsa-miR-27b | UUCACAGUGGCUAAGUUCUGC |
| Related to 26 cancers |  |
| Cluster of size 4 (#5) |  |
| hsa-miR-34a | UGGCAGUGUCUUAGCUGGUUGU |
| Related to 141 cancers |  |
| hsa-miR-34a | UGGCAGUGUCUUAGCUGGUUGU |
| Related to 141 cancers |  |
| hsa-miR-34a | UGGCAGUGUCUUAGCUGGUUGU |
| Related to 141 cancers |  |
| hsa-miR-34a | UGGCAGUGUCUUAGCUGGUUGU |
| Related to 141 cancers |  |
| Cluster of size 4 (#6) |  |
| hsa-miR-196b | UAGGUAGUUUCCUGUUGUUGGG |
| Related to 18 cancers |  |
| hsa-miR-196b | UAGGUAGUUUCCUGUUGUUGGG |
| Related to 18 cancers |  |
| hsa-miR-196a | UAGGUAGUUUCAUGUUGUUGGG |
| Related to 32 cancers |  |
| hsa-miR-196b | UAGGUAGUUUCCUGUUGUUGGG |
| Related to 18 cancers |  |
| Cluster of size 4 (#7) |  |
| hsa-miR-223 | UGUCAGUUUGUCAAAUACCCCA |
| Related to 69 cancers |  |
| hsa-miR-223 | UGUCAGUUUGUCAAAUACCCCA |
| Related to 69 cancers |  |
| hsa-miR-223 | UGUCAGUUUGUCAAAUACCCCA |
| Related to 69 cancers |  |
| hsa-miR-223-3p | UGUCAGUUUGUCAAAUACCCCA |
| Related to 11 cancers |  |
| Clusters with 5 sequences--(2 clusters)-- | |
| Cluster of size 5 (#1) |  |
| hsa-miR-29a | UAGCACCAUCUGAAAUCGGUUA |
| Related to 50 cancers |  |
| hsa-miR-29a-3p | UAGCACCAUCUGAAAUCGGUUA |
| Related to 4 cancers |  |
| hsa-miR-29a | UAGCACCAUCUGAAAUCGGUUA |
| Related to 50 cancers |  |
| hsa-miR-29b | UAGCACCAUUUGAAAUCAGUGUU |
| Related to 53 cancers |  |
| hsa-miR-29c | UAGCACCAUUUGAAAUCGGUUA |
| Related to 48 cancers |  |
| Cluster of size 5 (#2) |  |
| hsa-miR-31 | AGGCAAGAUGCUGGCAUAGCU |
| Related to 75 cancers |  |
| hsa-miR-31 | AGGCAAGAUGCUGGCAUAGCU |
| Related to 75 cancers |  |
| hsa-miR-31 | AGGCAAGAUGCUGGCAUAGCU |
| Related to 75 cancers |  |
| hsa-miR-31 | AGGCAAGAUGCUGGCAUAGCU |
| Related to 75 cancers |  |
| hsa-miR-31 | AGGCAAGAUGCUGGCAUAGCU |
| Related to 75 cancers |  |
| Clusters with 6 sequences--(1 clusters)-- | |
| Cluster of size 6 (#1) |  |
| hsa-miR-138 | AGCUGGUGUUGUGAAUCAGGCCG |
| Related to 61 cancers |  |
| hsa-miR-138 | AGCUGGUGUUGUGAAUCAGGCCG |
| Related to 61 cancers |  |
| hsa-miR-138 | AGCUGGUGUUGUGAAUCAGGCCG |
| Related to 61 cancers |  |
| hsa-miR-138 | AGCUGGUGUUGUGAAUCAGGCCG |
| Related to 61 cancers |  |
| hsa-miR-138-5p | AGCUGGUGUUGUGAAUCAGGCCG |
| Related to 9 cancers |  |
| hsa-miR-138 | AGCUGGUGUUGUGAAUCAGGCCG |
| Related to 61 cancers |  |
| Clusters with 7 sequences--(4 clusters)-- | |
| Cluster of size 7 (#1) |  |
| hsa-miR-16 | UAGCAGCACGUAAAUAUUGGCG |
| Related to 61 cancers |  |
| hsa-miR-195-5p | UAGCAGCACAGAAAUAUUGGC |
| Related to 10 cancers |  |
| hsa-miR-16 | UAGCAGCACGUAAAUAUUGGCG |
| Related to 61 cancers |  |
| hsa-miR-16 | UAGCAGCACGUAAAUAUUGGCG |
| Related to 61 cancers |  |
| hsa-miR-16 | UAGCAGCACGUAAAUAUUGGCG |
| Related to 61 cancers |  |
| hsa-miR-16 | UAGCAGCACGUAAAUAUUGGCG |
| Related to 61 cancers |  |
| hsa-miR-16 | UAGCAGCACGUAAAUAUUGGCG |
| Related to 61 cancers |  |
| Cluster of size 7 (#2) |  |
| hsa-let-7a | UGAGGUAGUAGGUUGUAUAGUU |
| Related to 81 cancers |  |
| hsa-let-7d | AGAGGUAGUAGGUUGCAUAGUU |
| Related to 22 cancers |  |
| hsa-let-7d | AGAGGUAGUAGGUUGCAUAGUU |
| Related to 22 cancers |  |
| hsa-let-7d | AGAGGUAGUAGGUUGCAUAGUU |
| Related to 22 cancers |  |
| hsa-let-7d | AGAGGUAGUAGGUUGCAUAGUU |
| Related to 22 cancers |  |
| hsa-miR-98 | UGAGGUAGUAAGUUGUAUUGUU |
| Related to 41 cancers |  |
| hsa-let-7f | UGAGGUAGUAGAUUGUAUAGUU |
| Related to 31 cancers |  |
| Cluster of size 7 (#3) |  |
| hsa-miR-125b | UCCCUGAGACCCUAACUUGUGA |
| Related to 69 cancers |  |
| hsa-miR-99a | AACCCGUAGAUCCGAUCUUGUG |
| Related to 32 cancers |  |
| hsa-miR-125a-5p | UCCCUGAGACCCUUUAACCUGUGA |
| Related to 18 cancers |  |
| hsa-miR-100 | AACCCGUAGAUCCGAACUUGUG |
| Related to 37 cancers |  |
| hsa-miR-125b | UCCCUGAGACCCUAACUUGUGA |
| Related to 69 cancers |  |
| hsa-miR-10b | UACCCUGUAGAACCGAAUUUGUG |
| Related to 54 cancers |  |
| hsa-miR-125b | UCCCUGAGACCCUAACUUGUGA |
| Related to 69 cancers |  |
| Cluster of size 7 (#4) |  |
| hsa-miR-221 | AGCUACAUUGUCUGCUGGGUUUC |
| Related to 98 cancers |  |
| hsa-miR-221 | AGCUACAUUGUCUGCUGGGUUUC |
| Related to 98 cancers |  |
| hsa-miR-222-3p | AGCUACAUCUGGCUACUGGGU |
| Related to 4 cancers |  |
| hsa-miR-221 | AGCUACAUUGUCUGCUGGGUUUC |
| Related to 98 cancers |  |
| hsa-miR-222 | AGCUACAUCUGGCUACUGGGU |
| Related to 61 cancers |  |
| hsa-miR-222 | AGCUACAUCUGGCUACUGGGU |
| Related to 61 cancers |  |
| hsa-miR-221 | AGCUACAUUGUCUGCUGGGUUUC |
| Related to 98 cancers |  |
| Clusters with 8 sequences--(1 clusters)-- | |
| Cluster of size 8 (#1) |  |
| hsa-miR-206 | UGGAAUGUAAGGAAGUGUGUGG |
| Related to 48 cancers |  |
| hsa-miR-1 | UGGAAUGUAAAGAAGUAUGUAU |
| Related to 3319 cancers |  |
| hsa-miR-206 | UGGAAUGUAAGGAAGUGUGUGG |
| Related to 48 cancers |  |
| hsa-miR-1 | UGGAAUGUAAAGAAGUAUGUAU |
| Related to 3319 cancers |  |
| hsa-miR-1 | UGGAAUGUAAAGAAGUAUGUAU |
| Related to 3319 cancers |  |
| hsa-miR-1 | UGGAAUGUAAAGAAGUAUGUAU |
| Related to 3319 cancers |  |
| hsa-miR-206 | UGGAAUGUAAGGAAGUGUGUGG |
| Related to 48 cancers |  |
| hsa-miR-206 | UGGAAUGUAAGGAAGUGUGUGG |
| Related to 48 cancers |  |
| Clusters with 9 sequences--(1 clusters)-- | |
| Cluster of size 9 (#1) |  |
| hsa-miR-375 | UUUGUUCGUUCGGCUCGCGUGA |
| Related to 70 cancers |  |
| hsa-miR-375 | UUUGUUCGUUCGGCUCGCGUGA |
| Related to 70 cancers |  |
| hsa-miR-375 | UUUGUUCGUUCGGCUCGCGUGA |
| Related to 70 cancers |  |
| hsa-miR-375 | UUUGUUCGUUCGGCUCGCGUGA |
| Related to 70 cancers |  |
| hsa-miR-375 | UUUGUUCGUUCGGCUCGCGUGA |
| Related to 70 cancers |  |
| hsa-miR-375 | UUUGUUCGUUCGGCUCGCGUGA |
| Related to 70 cancers |  |
| hsa-miR-375 | UUUGUUCGUUCGGCUCGCGUGA |
| Related to 70 cancers |  |
| hsa-miR-375 | UUUGUUCGUUCGGCUCGCGUGA |
| Related to 70 cancers |  |
| hsa-miR-375 | UUUGUUCGUUCGGCUCGCGUGA |
| Related to 70 cancers |  |
| Clusters with 13 sequences--(1 clusters)-- | |
| Cluster of size 13 (#1) |  |
| hsa-miR-21 | UAGCUUAUCAGACUGAUGUUGA |
| Related to 619 cancers |  |
| hsa-miR-21 | UAGCUUAUCAGACUGAUGUUGA |
| Related to 619 cancers |  |
| hsa-miR-21 | UAGCUUAUCAGACUGAUGUUGA |
| Related to 619 cancers |  |
| hsa-miR-21 | UAGCUUAUCAGACUGAUGUUGA |
| Related to 619 cancers |  |
| hsa-miR-21 | UAGCUUAUCAGACUGAUGUUGA |
| Related to 619 cancers |  |
| hsa-miR-21 | UAGCUUAUCAGACUGAUGUUGA |
| Related to 619 cancers |  |
| hsa-miR-21 | UAGCUUAUCAGACUGAUGUUGA |
| Related to 619 cancers |  |
| hsa-miR-21 | UAGCUUAUCAGACUGAUGUUGA |
| Related to 619 cancers |  |
| hsa-miR-21 | UAGCUUAUCAGACUGAUGUUGA |
| Related to 619 cancers |  |
| hsa-miR-21 | UAGCUUAUCAGACUGAUGUUGA |
| Related to 619 cancers |  |
| hsa-miR-21 | UAGCUUAUCAGACUGAUGUUGA |
| Related to 619 cancers |  |
| hsa-miR-21 | UAGCUUAUCAGACUGAUGUUGA |
| Related to 619 cancers |  |
| hsa-miR-21 | UAGCUUAUCAGACUGAUGUUGA |
| Related to 619 cancers |  |
| Clusters with 14 sequences--(1 clusters)-- | |
| Cluster of size 14 (#1) |  |
| hsa-miR-200b | UAAUACUGCCUGGUAAUGAUGA |
| Related to 45 cancers |  |
| hsa-miR-200b | UAAUACUGCCUGGUAAUGAUGA |
| Related to 45 cancers |  |
| hsa-miR-429 | UAAUACUGUCUGGUAAAACCGU |
| Related to 50 cancers |  |
| hsa-miR-200a | UAACACUGUCUGGUAACGAUGU |
| Related to 50 cancers |  |
| hsa-miR-200c | UAAUACUGCCGGGUAAUGAUGGA |
| Related to 64 cancers |  |
| hsa-miR-200b-3p | UAAUACUGCCUGGUAAUGAUGA |
| Related to 3 cancers |  |
| hsa-miR-200a | UAACACUGUCUGGUAACGAUGU |
| Related to 50 cancers |  |
| hsa-miR-200c | UAAUACUGCCGGGUAAUGAUGGA |
| Related to 64 cancers |  |
| hsa-miR-200c | UAAUACUGCCGGGUAAUGAUGGA |
| Related to 64 cancers |  |
| hsa-miR-200c | UAAUACUGCCGGGUAAUGAUGGA |
| Related to 64 cancers |  |
| hsa-miR-141 | UAACACUGUCUGGUAAAGAUGG |
| Related to 65 cancers |  |
| hsa-miR-429 | UAAUACUGUCUGGUAAAACCGU |
| Related to 50 cancers |  |
| hsa-miR-429 | UAAUACUGUCUGGUAAAACCGU |
| Related to 50 cancers |  |
| hsa-miR-141 | UAACACUGUCUGGUAAAGAUGG |
| Related to 65 cancers |  |

**Table S4.** over representation analyses for the three datasets DS1,DS2 and DS3 and their overall representation

| **Dataset** | **Pathway** | **Adj p.** | **Representation** |
| --- | --- | --- | --- |
| **DS-1** | **GO0014843** Skeletal muscle proliferation  **GO0042660** positive regulation of cell fate specification  **GO0060591** chondroblast differentiation  **GO0051541** elastin metabolic process  **GO0010764** negative regulation of fibroblast migration | 1.27e-8  1.27e-8  1.51e-8  9.46e-8  9.46e-8 | 0R  OR  OR  OR  OR |
| **DS-2** | **GO0014843** growth factor dependent regulation of skeletal muscle satellite cell  **GO0042660** proliferation positive regulation of cell fate specification  **GO0060591** chondroblast differentiation  **GO0010764** negative regulation of fibroblast migration  **GO2000546** positive regulation of endothelial cell chemotaxis to fibroblast growth factor | 1.04e-6  1.04e-6  1.06e-6  3.25e-6  3.25e-6 | 0R  OR  OR  OR  OR |
| **DS-3** | **GO0032274** gonadotropin secretion  **GO0044029** hypomethylation of CpG island  **GO0071072** negative regulation of phospholipid biosynthetic process  **GO0030948** negative regulation of vascular endothelial growth factor receptor  **GO0048743** signaling pathway positive regulation of skeletal muscle fiber development | 5.89e-10  5.89e-10  1.06e-7  8.07e-8  8.99e-8 | 0R  OR  OR  OR  OR |

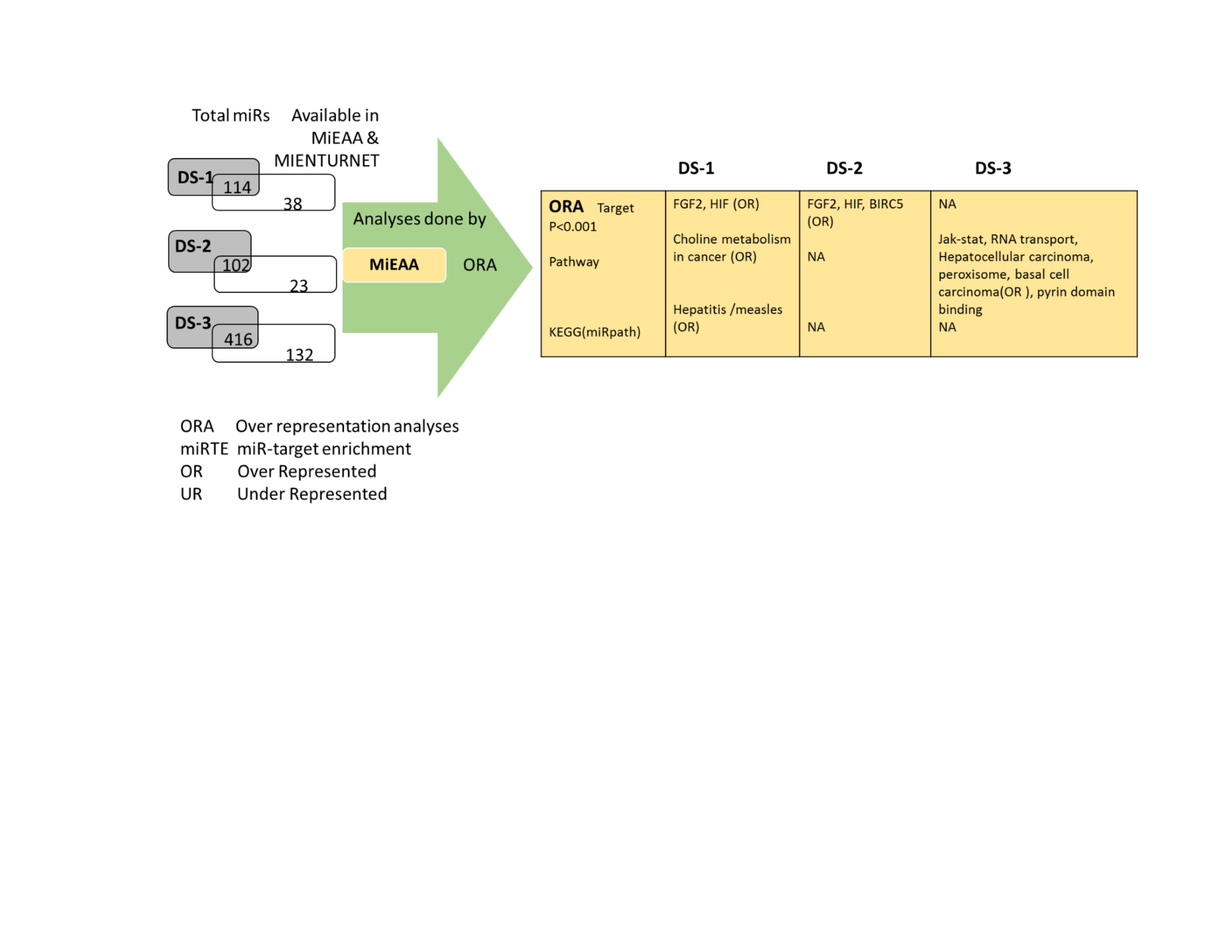

**Figure S1.** Brief results of miR over representation analyses

| **hsa-miR-184** | |
| --- | --- |
| **EPB41L5** | **-0.93** |
| **SF1** | **-0.91** |
| **AGO2** | **-0.83** |
| **CRTC1** | **-0.75** |
| **ALDH4A1** | **-0.72** |
| **FBXO28** | **-0.69** |
| **SIDT2** | **-0.68** |
| **NUS1** | **-0.67** |
| **ZNF740** | **-0.61** |
| **ZNF865** | **-0.6** |
| **HAND2** | **-0.6** |
| **SLC25A22** | **-0.6** |
| **C20orf112** | **-0.56** |

| **hsa-miR-107** | | |
| --- | --- | --- |
| **NEK10** | | **-1.03** |
| **AGFG1** | | **-0.76** |
| **RRAGC** | | **-0.62** |
| **KB-1507C5.2** | | **-0.58** |
| **AMMECR1** | | **-0.55** |
| **USP14** | | **-0.55** |
| **hsa-miR-200** | | |
| **AGO2** | **-1.12** | |
| **CDC73** | **-1.04** | |
| **RAP1B** | **-0.96** | |
| **CFL2** | **-0.89** | |
| **KLHL14** | **-0.75** | |
| **FEZ2** | **-0.72** | |
| **IMMP2L** | **-0.72** | |
| **WIPF1** | **-0.71** | |
| **KIAA0101** | **-0.7** | |
| **SEC23A** | **-0.67** | |
| **DZIP1** | **-0.66** | |
| **GPM6A** | **-0.65** | |
| **PAPD5** | **-0.65** | |
| **INTS8** | **-0.64** | |
| **AP1S2** | **-0.64** | |
| **CACUL1** | **-0.62** | |
| **IAH1** | **-0.62** | |
| **FBXO22** | **-0.62** | |
| **ERRFI1** | **-0.61** | |
| **CNEP1R1** | **-0.6** | |
| **RP6-24A23.6** | **-0.6** | |
| **HMGB3** | **-0.59** | |
| **ARL5A** | **-0.59** | |
| **CCNJ** | **-0.58** | |
| **APOO** | **-0.58** | |
| **LOX** | **-0.58** | |
| **SERPINI1** | **-0.57** | |
| **PTPN21** | **-0.57** | |
| **MFAP5** | **-0.57** | |
| **NUP153** | **-0.55** | |
| **PTHLH** | **-0.53** | |
| **RECK** | **-0.52** | |
| **TMEFF2** | **-0.51** | |
| **YWHAG** | **-0.51** | |
| **RAP2C** | **-0.51** | |
| **TMOD3** | **-0.51** | |
| **CEP41** | **-0.5** | |
| **B3GNT1** | **-0.5** | |
| **RNF2** | **-0.5** | |
| **PKIA** | **-0.5** | |

| **hsa-miR-133a-3p** | | | |
| --- | --- | --- | --- |
| **LHFP** | | | **-1.04** |
| **SEC61B** | | | **-0.98** |
| **CETN3** | | | **-0.87** |
| **FTL** | | | **-0.87** |
| **PTBP1** | | | **-0.85** |
| **LDLRAP1** | | | **-0.83** |
| **CLTA** | | | **-0.78** |
| **ACAT2** | | | **-0.78** |
| **SUMO1** | | | **-0.77** |
| **TFAP2D** | | | **-0.76** |
| **SLC50A1** | | | **-0.75** |
| **SYT2** | | | **-0.75** |
| **TMEM200B** | | | **-0.75** |
| **TIMM17A** | | | **-0.74** |
| **PPP2R2D** | | | **-0.74** |
| **BICC1** | | | **-0.72** |
| **XXYLT1** | | | **-0.71** |
| **VKORC1** | | | **-0.7** |
| **ZNF354A** | | | **-0.7** |
| **SAMD5** | | | **-0.7** |
| **LASP1** | | | **-0.68** |
| **PRRT2** | | | **-0.67** |
| **FOSL2** | | | **-0.67** |
| **EIF4A1** | | | **-0.66** |
| **AL117190.3** | | | **-0.66** |
| **TTPAL** | | | **-0.66** |
| **STXBP6** | | | **-0.66** |
| **ZIC3** | | | **-0.65** |
| **PPP2CA** | | | **-0.65** |
| **ENPP5** | | | **-0.65** |
| **CUL4B** | | | **-0.65** |
| **TMEM170B** | | | **-0.64** |
| **UBXN7** | | | **-0.64** |
| **ZC3H14** | | | **-0.64** |
| **PAX7** | | | **-0.63** |
| **C12orf43** | | | **-0.63** |
| **SMARCD1** | | | **-0.63** |
| **COL25A1** | | | **-0.63** |
| **SLC30A7** | | | **-0.62** |
| **SGMS2** | | | **-0.62** |
| **GDNF** | | | **-0.62** |
| **TPM4** | | | **-0.62** |
| **TMOD3** | | | **-0.61** |
| **RLN2** | | | **-0.61** |
| **SIMC1** | | | **-0.6** |
| **GPM6A** | | | **-0.6** |
| **DOLPP1** | | | **-0.6** |
| **PTPRZ1** | | | **-0.6** |
| **TMEM167A** | | | **-0.6** |
| **CMPK1** | | | **-0.6** |
| **FGF1** | | | **-0.6** |
| **LHX9** | | | **-0.59** |
| **VPS54** | | | **-0.59** |
| **SLC6A1** | | | **-0.58** |
| **CNN2** | | | **-0.58** |
| **KIRREL** | | | **-0.58** |
| **LANCL2** | | | **-0.58** |
| **RBMX** | | | **-0.58** |
| **RBM23** | | | **-0.58** |
| **QKI** | | | **-0.57** |
| **NAA40** | | | **-0.57** |
| **CMTM6** | | | **-0.57** |
| **DDX19B** | | | **-0.57** |
| **AGRP** | | | **-0.56** |
| **SGPP1** | | | **-0.56** |
| **NRIP3** | | | **-0.56** |
| **ELP5** | | | **-0.56** |
| **FAM57A** | | | **-0.55** |
| **EMP2** | | | **-0.55** |
| **RAP2C** | | | **-0.55** |
| **VEGFC** | | | **-0.55** |
| **DBNL** | | | **-0.55** |
| **CCNDBP1** | | | **-0.55** |
| **FOXL2** | | | **-0.54** |
| **CAPN15** | | | **-0.54** |
| **UBA2** | | | **-0.54** |
| **KIF3C** | | | **-0.54** |
| **AFTPH** | | | **-0.54** |
| **B3GALNT1** | | | **-0.54** |
| **TXLNA** | | | **-0.54** |
| **GABARAPL1** | | | **-0.53** |
| **TRIM44** | | | **-0.53** |
| **SFXN2** | | | **-0.53** |
| **SOBP** | | | **-0.53** |
| **CCDC30** | | | **-0.53** |
| **GABPB2** | | | **-0.52** |
| **SLC6A6** | | | **-0.52** |
| **RAD51L3-RFFL** | | | **-0.52** |
| **ELFN1** | | | **-0.51** |
| **STOM** | | | **-0.51** |
| **NAGS** | | | **-0.51** |
| **ANKRD28** | | | **-0.51** |
| **DCBLD1** | | | **-0.51** |
| **MAP3K2** | | | **-0.51** |
| **TM9SF3** | | | **-0.51** |
| **TNFRSF10B** | | | **-0.51** |
| **PTBP3** | | | **-0.51** |
| **GNB4** | | | **-0.51** |
| **CRTAM** | | | **-0.5** |
| **ZC3H11A** | | | **-0.5** |
| **PLEKHA3** | | | **-0.5** |
| **DCLRE1A** | | | **-0.5** |
| **GCH1** | | | **-0.5** |
| **PTPRK** | | | **-0.5** |
| **COL8A1** | | | **-0.5** |
| **FOXP2** | | | **-0.5** |
| **hsa-miR-221-3p** | | | |
| **TFG** | | **-0.98** | |
| **NGRN** | | **-0.94** | |
| **VAPB** | | **-0.93** | |
| **GABRA1** | | **-0.88** | |
| **EIF5A2** | | **-0.88** | |
| **TMSB15B** | | **-0.83** | |
| **SNX4** | | **-0.83** | |
| **NAP1L5** | | **-0.81** | |
| **WSB2** | | **-0.76** | |
| **SEC62** | | **-0.74** | |
| **BEAN1** | | **-0.73** | |
| **ATAD2B** | | **-0.73** | |
| **WEE1** | | **-0.71** | |
| **IRX5** | | **-0.7** | |
| **VGLL4** | | **-0.7** | |
| **NTF3** | | **-0.69** | |
| **CREBZF** | | **-0.69** | |
| **RIMS3** | | **-0.67** | |
| **OSTM1** | | **-0.66** | |
| **TCF12** | | **-0.66** | |
| **ZNF652** | | **-0.66** | |
| **HMBOX1** | | **-0.65** | |
| **FNIP2** | | **-0.65** | |
| **HECTD2** | | **-0.64** | |
| **STMN1** | | **-0.63** | |
| **DCUN1D1** | | **-0.62** | |
| **GGA2** | | **-0.62** | |
| **TUBA1A** | | **-0.61** | |
| **CLVS2** | | **-0.61** | |
| **KIT** | | **-0.61** | |
| **MBD2** | | **-0.6** | |
| **CCT4** | | **-0.6** | |
| **RFX7** | | **-0.59** | |
| **PAIP1** | | **-0.59** | |
| **DMRT3** | | **-0.58** | |
| **PDE3A** | | **-0.58** | |
| **ARF4** | | **-0.58** | |
| **SMARCA5** | | **-0.57** | |
| **SRSF10** | | **-0.57** | |
| **MARK1** | | **-0.57** | |
| **NOVA1** | | **-0.57** | |
| **MIDN** | | **-0.56** | |
| **AGFG1** | | **-0.56** | |
| **STYX** | | **-0.56** | |
| **TP53BP2** | | **-0.54** | |
| **HNRNPA3** | | **-0.54** | |
| **ZFAND5** | | **-0.54** | |
| **DCAF12** | | **-0.53** | |
| **FRS2** | | **-0.53** | |
| **PAIP2** | | **-0.52** | |
| **PCMTD1** | | **-0.52** | |
| **MYBL1** | | **-0.52** | |
| **MYLIP** | | **-0.52** | |
| **ERBB4** | | **-0.52** | |
| **CACNB4** | | **-0.52** | |
| **NSUN4** | | **-0.52** | |
| **RNF4** | | **-0.51** | |
| **FRK** | | **-0.51** | |
| **FAM214A** | | **-0.5** | |
| **ZBTB37** | | **-0.5** | |
| **IRF2** | | **-0.5** | |
| **hsa-miR-451a** | | | |
| **PSMB8** | | **-1.09** | |
| **OSR1** | | **-0.93** | |
| **ATF2** | | **-0.91** | |
| **MIF** | | **-0.6** | |
| **hsa-miR-133-b** | | | |
| **TAGLN2** | | **-0.93** | |
| **PPP2CB** | | **-0.63** | |
| **C11orf58** | | **-0.58** | |
| **ZNF131** | | **-0.57** | |
| **SGTB** | | **-0.51** | |

| **hsa-miR-1-3p** | | | | |
| --- | --- | --- | --- | --- |
| **ARPC3** | | | **-1.3** | |
| **BDNF** | | | **-0.95** | |
| **SLC10A7** | | | **-0.9** | |
| **SEC23B** | | | **-0.88** | |
| **TAGLN2** | | | **-0.85** | |
| **POGK** | | | **-0.82** | |
| **ERMP1** | | | **-0.81** | |
| **SERP1** | | | **-0.8** | |
| **IGF1** | | | **-0.8** | |
| **UST** | | | **-0.75** | |
| **MMD2** | | | **-0.69** | |
| **G6PD** | | | **-0.68** | |
| **CDK14** | | | **-0.68** | |
| **VAMP4** | | | **-0.68** | |
| **PHAX** | | | **-0.67** | |
| **TNKS2** | | | **-0.66** | |
| **ALX1** | | | **-0.66** | |
| **SRI** | | | **-0.65** | |
| **C5orf51** | | | **-0.65** | |
| **RIT2** | | | **-0.64** | |
| **SLC44A1** | | | **-0.64** | |
| **RAB43** | | | **-0.64** | |
| **ARCN1** | | | **-0.63** | |
| **MMD** | | | **-0.61** | |
| **SRSF9** | | | **-0.61** | |
| **ANKRD29** | | | **-0.61** | |
| **EIF1AX** | | | **-0.6** | |
| **NDRG3** | | | **-0.59** | |
| **TIMP3** | | | **-0.58** | |
| **CREBL2** | | | **-0.58** | |
| **FOXP1** | | | **-0.58** | |
| **CNKSR3** | | | **-0.58** | |
| **NANP** | | | **-0.57** | |
| **SLC8A1** | | | **-0.57** | |
| **MAB21L1** | | | **-0.56** | |
| **NXT2** | | | **-0.55** | |
| **44076** | | | **-0.55** | |
| **GPR137C** | | | **-0.55** | |
| **HNRNPU** | | | **-0.55** | |
| **C9orf152** | | | **-0.54** | |
| **RABGAP1L** | | | **-0.54** | |
| **ANXA4** | | | **-0.54** | |
| **RSBN1L** | | | **-0.54** | |
| **SLC35B4** | | | **-0.54** | |
| **RAB5A** | | | **-0.53** | |
| **C2orf69** | | | **-0.53** | |
| **HNRNPA3** | | | **-0.53** | |
| **SLC30A9** | | | **-0.53** | |
| **TMEM178A** | | | **-0.52** | |
| **ZNF281** | | | **-0.52** | |
| **ZFP36L1** | | | **-0.52** | |
| **YWHAZ** | | | **-0.52** | |
| **BMPR1B** | | | **-0.52** | |
| **E2F5** | | | **-0.51** | |
| **LASP1** | | | **-0.51** | |
| **CCSAP** | | | **-0.51** | |
| **TWF1** | | | **-0.5** | |
| **SPRED1** | | | **-0.5** | |
| **GCH1** | | | **-0.5** | |
| **CITED2** | | | **-0.5** | |
| **SMIM2** | | | **-0.5** | |
| **hsa-miR-429** | | | | |
| **GMFB** | **-1** | | | |
| **ZEB1** | **-0.93** | | | |
| **FAM8A1** | **-0.78** | | | |
| **TCEB1** | **-0.73** | | | |
| **C16orf52** | **-0.71** | | | |
| **LHFP** | **-0.68** | | | |
| **PMAIP1** | **-0.68** | | | |
| **NR5A2** | **-0.64** | | | |
| **HIPK3** | **-0.63** | | | |
| **ZFPM2** | **-0.61** | | | |
| **VASH2** | **-0.6** | | | |
| **PPP4R2** | **-0.59** | | | |
| **NTF3** | **-0.58** | | | |
| **SUGT1** | **-0.58** | | | |
| **TIMP2** | **-0.58** | | | |
| **C16orf72** | **-0.57** | | | |
| **MCFD2** | **-0.56** | | | |
| **MARCKS** | **-0.56** | | | |
| **C6orf120** | **-0.56** | | | |
| **ASF1A** | **-0.56** | | | |
| **TFAP2A** | **-0.56** | | | |
| **PSAT1** | **-0.55** | | | |
| **THAP1** | **-0.54** | | | |
| **GTF2E1** | **-0.53** | | | |
| **ANGEL2** | **-0.52** | | | |
| **RASSF8** | **-0.51** | | | |
| **ELAVL2** | **-0.51** | | | |
| **ZFAND6** | **-0.5** | | | |
| **PRKACB** | **-0.5** | | | |
| **HNRNPD** | **-0.5** | | | |
| **hsa-miR-9-5p** | | | | |
| **ONECUT2** | **-5.98** | | | |
| **ONECUT1** | **-4.69** | | | |
| **POU2F2** | | **-1.44** | | |
| **TRPM7** | | **-1.36** | | |
| **YBX3** | | **-1.25** | | |
| **POU2F1** | | **-1.2** | | |
| **LYVE1** | | **-1.15** | | |
| **SLC50A1** | | **-0.95** | | |
| **ZBTB20** | | **-0.82** | | |
| **POU6F2** | | **-0.78** | | |
| **MTHFD2** | | **-0.77** | | |
| **C2ORF15** | | **-0.76** | | |
| **AP1S2** | | **-0.71** | | |
| **CTNNA1** | | **-0.68** | | |
| **RNF146** | | **-0.67** | | |
| **ANP32B** | | **-0.67** | | |
| **PDK4** | | **-0.66** | | |
| **ID4** | | **-0.66** | | |
| **LDLRAP1** | | **-0.65** | | |
| **SGMS2** | | **-0.65** | | |
| **SLC8A1** | | **-0.65** | | |
| **RPAP2** | | **-0.65** | | |
| **BRAF** | | **-0.63** | | |
| **SPTLC2** | | **-0.63** | | |
| **KCNJ2** | | **-0.62** | | |
| **FSTL1** | | **-0.62** | | |
| **SDC2** | | **-0.62** | | |
| **FRMD6** | | **-0.61** | | |
| **SLC31A2** | | **-0.6** | | |
| **TNFAIP8** | | **-0.6** | | |
| **CHRM3** | | **-0.59** | | |
| **RIC3** | | **-0.58** | | |
| **LIN28B** | | **-0.58** | | |
| **PRDM6** | | **-0.58** | | |
| **RAB34** | | **-0.58** | | |
| **SOCS5** | | **-0.58** | | |
| **TGFBI** | | **-0.58** | | |
| **GOT1** | | **-0.58** | | |
| **ANK2** | | **-0.57** | | |
| **MYOCD** | | **-0.57** | | |
| **SFXN2** | | **-0.56** | | |
| **C9orf89** | | **-0.56** | | |
| **C2orf88** | | **-0.56** | | |
| **MAGT1** | | **-0.56** | | |
| **RNF150** | | **-0.56** | | |
| **SNX25** | | **-0.56** | | |
| **CLDN14** | | **-0.55** | | |
| **VAV3** | | **-0.55** | | |
| **CCNDBP1** | | **-0.55** | | |
| **C17orf85** | | **-0.55** | | |
| **CXCL11** | | **-0.54** | | |
| **FAM83B** | | **-0.54** | | |
| **ENPEP** | | **-0.54** | | |
| **PRDM1** | | **-0.54** | | |
| **RANBP17** | | **-0.53** | | |
| **MESDC1** | | **-0.53** | | |
| **SHC1** | | **-0.53** | | |
| **UHMK1** | | **-0.53** | | |
| **FKBP7** | | **-0.53** | | |
| **C4orf46** | | **-0.52** | | |
| **TAL2** | | **-0.52** | | |
| **CREG2** | | **-0.52** | | |
| **FOXP1** | | **-0.52** | | |
| **ITM2B** | | **-0.52** | | |
| **GALNT3** | | **-0.51** | | |
| **TNC** | | **-0.51** | | |
| **NXPE3** | | **-0.51** | | |
| **ARMCX2** | | **-0.51** | | |
| **MTMR2** | | **-0.51** | | |
| **RHOQ** | | **-0.51** | | |
| **DNAJC14** | | **-0.51** | | |
| **GDE1** | | **-0.51** | | |
| **SLC16A10** | | **-0.51** | | |
| **FAM46D** | | **-0.5** | | |
| **GABRB2** | | **-0.5** | | |
| **PPCS** | | **-0.5** | | |
| **CCNG1** | | **-0.5** | | |
| **NID2** | | **-0.5** | | |
| **SUDS3** | | **-0.5** | | |
| **hsa-miR-9-3p** | | | | |
| **ZNF99** | | | | **-1.04** |
| **C10orf11** | | | | **-0.97** |
| **C12orf36** | | | | **-0.94** |
| **AC090186.1** | | | | **-0.7** |
| **ACTL6A** | | | | **-0.7** |
| **DSCC1** | | | | **-0.72** |
| **MZT1** | | | | **-0.67** |
| **NAA20** | | | | **-0.67** |
| **GTF2H5** | | | | **-0.68** |
| **RP11-542P2.1** | | | | **-0.66** |
| **GTPBP8** | | | | **-0.92** |
| **MTRNR2L8** | | | | **-0.65** |
| **GPR19** | | | | **-0.64** |
| **MPPED2** | | | | **-0.64** |
| **GABRA1** | | | | **-0.62** |
| **MTRNR2L13** | | | | **-0.61** |
| **HIST1H2BG** | | | | **-0.8** |
| **NXT2** | | | | **-0.59** |
| **KITLG** | | | | **-0.75** |
| **KLLN** | | | | **-0.59** |
| **MTRNR2L6** | | | | **-0.59** |
| **ITGB1** | | | | **-0.58** |
| **STARD4** | | | | **-0.72** |
| **MTRNR2L1** | | | | **-0.58** |
| **MTRNR2L10** | | | | **-0.58** |
| **ACN9** | | | | **-0.59** |
| **SMIM19** | | | | **-0.57** |
| **YIPF5** | | | | **-0.61** |
| **MTRNR2L3** | | | | **-0.55** |
| **MTRNR2L11** | | | | **-0.55** |
| **PPA1** | | | | **-0.55** |
| **USP51** | | | | **-0.55** |
| **RBM48** | | | | **-0.64** |
| **MTRNR2L7** | | | | **-0.54** |
| **TMEM255A** | | | | **-0.53** |
| **MDK** | | | | **-0.53** |
| **ZFHX3** | | | | **-0.55** |
| **TTC8** | | | | **-0.52** |
| **ARL8B** | | | | **-0.56** |
| **CPA3** | | | | **-0.52** |
| **OR7A17** | | | | **-0.52** |
| **XAGE5** | | | | **-0.51** |
| **ONECUT1** | | | | **-0.55** |
| **POLR1C** | | | | **-0.53** |
| **GCG** | | | | **-0.5** |
| **hsa-miR-141-3p** | | | | |
| **ZNF385D** | | **-0.95** | | |
| **C3orf14** | | **-0.78** | | |
| **ZEB1** | | **-0.76** | | |
| **ZFR** | | **-0.75** | | |
| **ACOT7** | | **-0.74** | | |
| **LMO3** | | **-0.72** | | |
| **PHB2** | | **-0.72** | | |
| **DR1** | | **-0.69** | | |
| **PGRMC2** | | **-0.65** | | |
| **ZEB2** | | **-0.65** | | |
| **C1orf21** | | **-0.64** | | |
| **ANP32E** | | **-0.63** | | |
| **FUS** | | **-0.62** | | |
| **KCTD20** | | **-0.62** | | |
| **GSG1** | | **-0.61** | | |
| **P2RY1** | | **-0.61** | | |
| **C4orf29** | | **-0.6** | | |
| **SDC2** | | **-0.59** | | |
| **DEK** | | **-0.58** | | |
| **RNF145** | | **-0.56** | | |
| **TADA1** | | **-0.56** | | |
| **YAF2** | | **-0.56** | | |
| **DMWD** | | **-0.55** | | |
| **MSANTD4** | | **-0.54** | | |
| **SLC35D1** | | **-0.54** | | |
| **YWHAG** | | **-0.54** | | |
| **MAGED4** | | **-0.53** | | |
| **SCHIP1** | | **-0.53** | | |
| **UGT1A1** | | **-0.53** | | |
| **UGT1A10** | | **-0.53** | | |

| **UGT1A9** \| **-0.53**; **UGT1A3** \| **-0.53**; **UGT1A4** \| **-0.53**; **UGT1A5** \| **-0.53**; **UGT1A6** \| **-0.53**; **UGT1A3** \| **-0.53**; **UGT1A4** \| **-0.53**; **UGT1A7** \| **-0.53**; **UGT1A8** \| **-0.53**; **MAGED4B** \| **-0.52**; **RBM33** \| **-0.52**; **TGFB2** \| **-0.52**; **KATNAL1** \| **-0.51**; **LYSMD3** \| **-0.51**; **SOBP** \| **-0.51**; **STAT4** \| **-0.51**; **STRN** \| **-0.51**; **DCTN6** \| **-0.5**; **DLC1** \| **-0.5**; **DUSP3** \| **-0.5** | | | | | |
| --- | --- | --- | --- | --- | --- |
| **hsa-miR-206** | | | | |  |
| **SMIM14** | | | | | **-1.38** |
| **PTPLAD1** | | | | | **-1.11** |
| **CORO1C** | | | | | **-1.09** |
| **TTR -1.07** | | | | | |
| **GJA1** | | **-0.84** | | | |
| **TMSB4X** | | **-0.74** | | | |
| **GLCCI1** | | **-0.73** | | | |
| **RABEPK** | | **-0.65** | | | |
| **KCNIP3** | | **-0.63** | | | |
| **TMEM243** | | **-0.62** | | | |
| **EBPL** | | **-0.61** | | | |
| **TPM4** | | **-0.6** | | | |
| **ANXA2** | | **-0.59** | | | |
| **CDC42SE1** | | **-0.56** | | | |
| **GPD2** | | **-0.55** | | | |
| **UHMK1** | | **-0.55** | | | |
| **PAX3** | | **-0.54** | | | |
| **TIGD6** | | **-0.54** | | | |
| **SLC15A2** | | **-0.54** | | | |
| **MAL2** | | **-0.53** | | | |
| **MXD1** | | **-0.53** | | | |
| **ABHD2** | | **-0.53** | | | |
| **CCND2** | | **-0.53** | | | |
| **RASA1** | | **-0.53** | | | |
| **PPIB** | | **-0.52** | | | |
| **CDON** | | **-0.51** | | | |
| **SLC29A3** | | **-0.5** | | | |
| **hsa-miR-221-5p** | | | | | |
| **VPS53** | | **-3.45** | | | |
| **EVC** | | **-2.65** | | | |
| **LSM5** | | **-1.17** | | | |
| **ZNF526** | | | **-1.11** | | |
| **TRAF1** | | | | **-1.06** | |
| **MRGPRF** | | | | **-0.96** | |
| **PPIL4** | | | | **-0.95** | |
| **NRIP2** | | | | **-0.94** | |
| **RRP15** | | | | **-0.94** | |
| **GLA** | | | | **-0.9** | |
| **WDR31** | | | | **-0.87** | |
| **C17orf103** | | | | **-0.87** | |
| **AGBL5** | | | | **-0.86** | |
| **ALDH1A2** | | | | **-0.85** | |
| **BSN** | | | | **-0.85** | |
| **G6PC** | | | | **-0.85** | |
| **CLPB** | | | | **-0.85** | |
| **COL20A1** | | | | **-0.84** | |
| **SCUBE1** | | | | **-0.84** | |
| **RPL37A** | | | | **-0.83** | |
| **RAD51** | | | | **-0.82** | |
| **KIAA1644** | | | | **-0.81** | |
| **RBM23** | | | | **-0.81** | |
| **HEBP2** | | | | **-0.81** | |
| **C19orf73** | | | | **-0.8** | |
| **RRP7A** | | | | **-0.8** | |
| **TTC39A** | | | | **-0.8** | |
| **DPH3** | | | | **-0.8** | |
| **PRR13** | | | | **-0.8** | |
| **NDUFAF3** | | | | **-0.79** | |
| **KIF1C** | | | | **-0.78** | |
| **TMED1** | | | | **-0.78** | |
| **PNPO** | | | | **-0.77** | |
| **RP11-114H20.1** | | | | **-0.76** | |
| **CBX5** | | | | **-0.76** | |
| **GOLGA7B** | | | | **-0.76** | |
| **CAMK2N1** | | | | **-0.76** | |
| **RP11-363G10.2** | | | | **-0.75** | |
| **F2RL3** | | | | **-0.74** | |
| **C11orf83** | | | | **-0.73** | |
| **NRIP3** | | | | **-0.72** | |
| **PRIM1** | | | | **-0.72** | |
| **ZFP69B** | | | | **-0.71** | |
| **RPS12** | | | | **-0.71** | |
| **MAVS** | | | | **-0.7** | |
| **ACOT2** | | | | **-0.7** | |
| **CAPN5** | | | | **-0.69** | |
| **GGCX** | | | | **-0.69** | |
| **HES3** | | | | **-0.68** | |
| **SDHAF1** | | | | **-0.68** | |
| **POC1A** | | | | **-0.67** | |
| **NCAPH** | | | | **-0.67** | |
| **C14orf23** | | | | **-0.66** | |
| **C1QTNF2** | | | | **-0.66** | |
| **TIMM10** | | | | **-0.66** | |
| **ZSCAN5A** | | | | **-0.65** | |
| **TMEM92** | | | | **-0.65** | |
| **UBL4A** | | | | **-0.64** | |
| **HN1L** | | | | **-0.64** | |
| **TYRO3** | | | | **-0.64** | |
| **LAX1** | | | | **-0.64** | |
| **RAB3B** | | | | **-0.64** | |
| **PSD2** | | | | **-0.63** | |
| **C11orf48** | | | | **-0.63** | |
| **FAM118A** | | | | **-0.63** | |
| **NOA1** | | | | **-0.63** | |
| **ATG13** | | | | **-0.62** | |
| **YME1L1** | | | | **-0.62** | |
| **ACTR1A** | | | | **-0.62** | |
| **LIPG** | | | | **-0.62** | |
| **CLEC4C** | | | | **-0.62** | |
| **IL12B** | | | | **-0.61** | |
| **IRAK4** | | | | **-0.61** | |
| **TTC39C** | | | | **-0.61** | |
| **MRTO4** | | | | **-0.61** | |
| **ELAC2** | | | | **-0.61** | |
| **C12orf79** | | | | **-0.6** | |
| **GMEB1** | | | | **-0.6** | |
| **FOSL2** | | | | **-0.6** | |
| **ARMCX3** | | | | **-0.6** | |
| **MSRB1** | | | | **-0.6** | |
| **AC106017.1** | | | | **-0.6** | |
| **HIST1H2AC** | | | | **-0.59** | |
| **KMT2A** | | | | **-0.59** | |
| **ZNF589** | | | | **-0.59** | |
| **ANXA11** | | | | **-0.59** | |
| **DIS3** | | | | **-0.59** | |
| **PELI3** | | | | **-0.59** | |
| **UPP1** | | | | **-0.59** | |
| **ADORA2B** | | | | **-0.59** | |
| **COX6B1** | | | | **-0.59** | |
| **QDPR** | | | | **-0.59** | |
| **NDUFB10** | | | | **-0.58** | |
| **HPCAL4** | | | | **-0.58** | |
| **IFRG15** | | | | **-0.58** | |
| **WSCD1** | | | | **-0.58** | |
| **SLC30A7** | | | | **-0.58** | |
| **ZNF445** | | | | **-0.58** | |
| **KNOP1** | | | | **-0.58** | |
| **IFNLR1** | | | | **-0.58** | |
| **FGG** | | | | **-0.58** | |
| **RWDD3** | | | | **-0.57** | |
| **CABP5** | | | | **-0.57** | |
| **WIPF3** | | | | **-0.57** | |
| **SNTN** | | | | **-0.57** | |
| **ZNF321P** | | | | **-0.57** | |
| **GFER** | | | | **-0.57** | |
| **NT5C1A** | | | | **-0.56** | |
| **TMEM154** | | | | **-0.56** | |
| **DMBX1** | | | | **-0.56** | |
| **CCDC115** | | | | **-0.56** | |
| **RAP2B** | | | | **-0.56** | |
| **RBCK1** | | | | **-0.56** | |
| **FDX1L** | | | | **-0.56** | |
| **CRCP** | | | | **-0.56** | |
| **SLC25A16** | | | | **-0.56** | |
| **NAA38** | | | | **-0.56** | |
| **EDN2** | | | | **-0.55** | |
| **PPP1R1B** | | | | **-0.55** | |
| **MEX3C** | | | | **-0.55** | |
| **REC8** | | | | **-0.55** | |
| **AP1S2** | | | | **-0.55** | |
| **SAMD14** | | | | **-0.55** | |
| **RNMT** | | | | **-0.55** | |
| **STX17** | | | | **-0.55** | |
| **MRGBP** | | | | **-0.55** | |
| **ZNF527** | | | | **-0.55** | |
| **TTLL6** | | | | **-0.55** | |
| **PPBP** | | | | **-0.55** | |
| **INSIG2** | | | | **-0.54** | |
| **DNAJB6** | | | | **-0.54** | |
| **ZNF791** | | | | **-0.54** | |
| **MED22** | | | | **-0.54** | |
| **TRAPPC3** | | | | **-0.54** | |
| **NHP2L1** | | | | **-0.54** | |
| **SF3A1** | | | | **-0.54** | |
| **MEX3B** | | | | **-0.53** | |
| **LDLRAD2** | | | | **-0.53** | |
| **COL4A2** | | | | **-0.53** | |
| **COPE** | | | | **-0.53** | |
| **SKA1** | | | | **-0.53** | |
| **ZIK1** | | | | **-0.53** | |
| **CEP41** | | | | **-0.53** | |
| **XRCC5** | | | | **-0.53** | |
| **NKX6-3** | | | | **-0.52** | |
| **SMAGP** | | | | **-0.52** | |
| **NADK** | | | | **-0.52** | |
| **ZNF708** | | | | **-0.52** | |
| **CD68** | | | | **-0.52** | |
| **ZNF207** | | | | **-0.52** | |
| **SLC5A3** | | | | **-0.52** | |
| **LRRD1** | | | | **-0.52** | |
| **ZNF394** | | | | **-0.52** | |
| **TIRAP** | | | | **-0.52** | |
| **CD84** | | | | **-0.52** | |
| **RBM48** | | | | **-0.52** | |
| **ATP1B1** | | | | **-0.52** | |
| **LLPH** | | | | **-0.52** | |
| **CRP** | | | | **-0.52** | |
| **CYTL1** | | | | **-0.51** | |
| **EPO** | | | | **-0.51** | |
| **RNF222** | | | | **-0.51** | |
| **TXLNA** | | | | **-0.51** | |
| **ZNF587** | | | | **-0.51** | |
| **DNAJC22** | | | | **-0.51** | |
| **SMUG1** | | | | **-0.51** | |
| **BAK1** | | | | **-0.51** | |
| **ZBTB3** | | | | **-0.51** | |
| **NPR1** | | | | **-0.51** | |
| **TUBD1** | | | | **-0.51** | |
| **TMEM120B** | | | | **-0.51** | |
| **IPO9** | | | | **-0.51** | |
| **UQCRQ** | | | | **-0.51** | |
| **FBXL7** | | | | **-0.5** | |
| **CRX** | | | | **-0.5** | |
| **CHRNA1** | | | | **-0.5** | |
| **C10orf25** | | | | **-0.5** | |
| **DAPK2** | | | | **-0.5** | |
| **LEPREL2** | | | | **-0.5** | |
| **POLH** | | | | **-0.5** | |
| **BMF** | | | | **-0.5** | |
| **DNAJC28** | | | | **-0.5** | |
| **LIG3** | | | | **-0.5** | |
| **C5orf49** | | | | **-0.5** | |
| **TMEM170A** | | | | **-0.5** | |
| **GRM4** | | | | **-0.5** | |
| **FAM118B** | | | | **-0.5** | |
| **C3orf38** |  |  |  |  |  |
| **ELAVL4** |  |  |  |  |  |

**Table S5**. Targets obtained from Target scan with their cutoff values

**Figure S2.** PPI analyses using STRING and interpreted using CYTOSCAPE for all selected miR’s

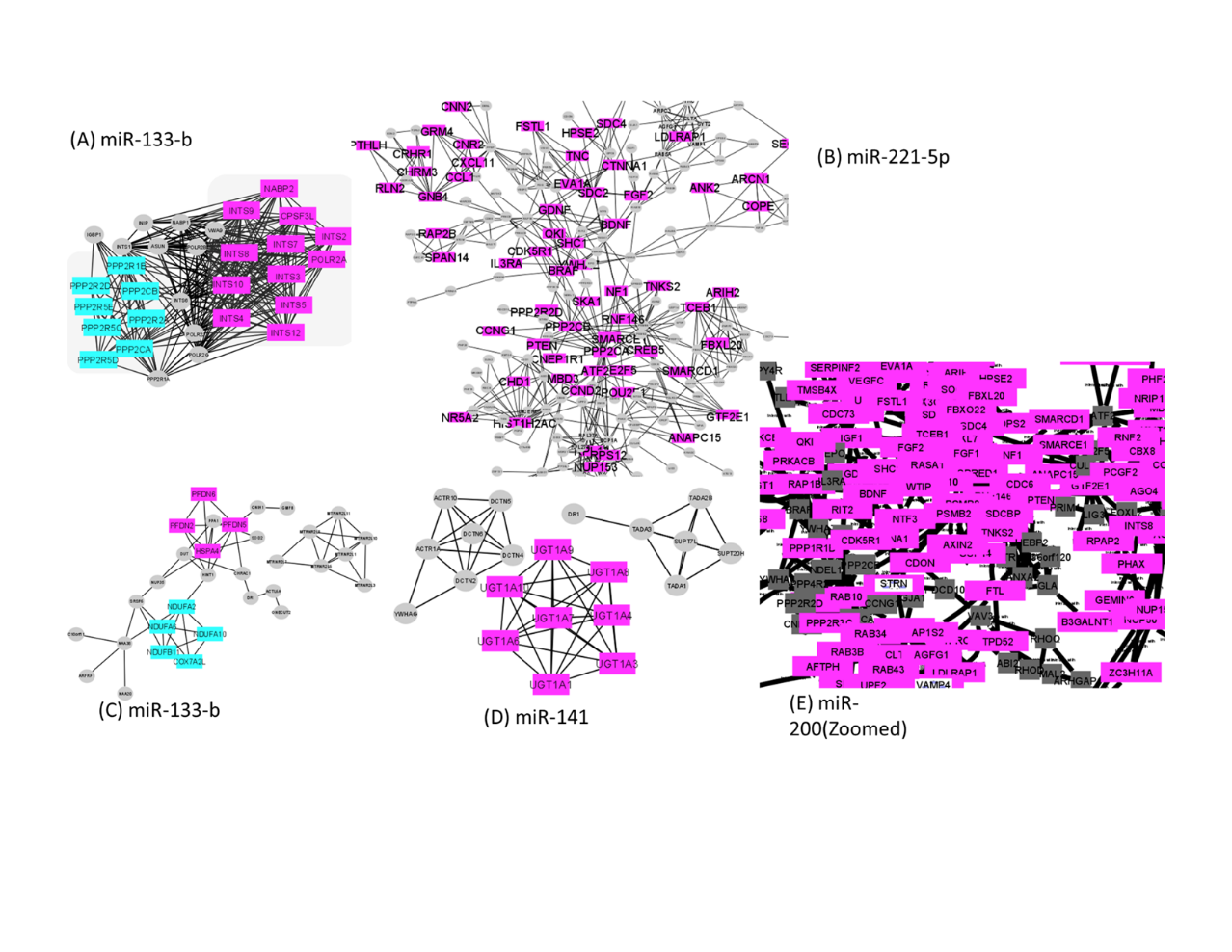

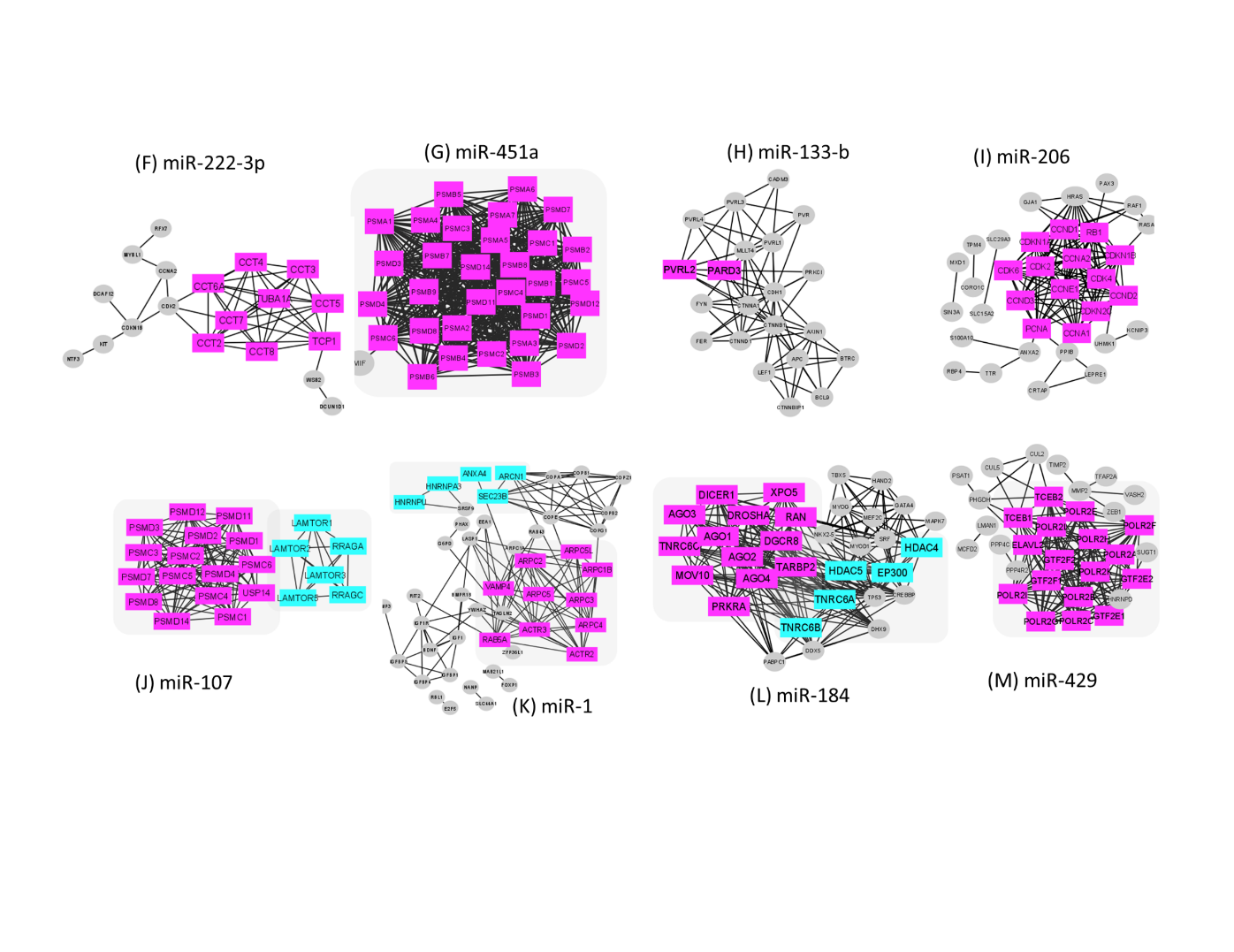

**Figure S3.** ∆Ct values of blood and tissue for each miR in controls , OSCC and OSF (A) Controls (B) OSCC (C) OSF

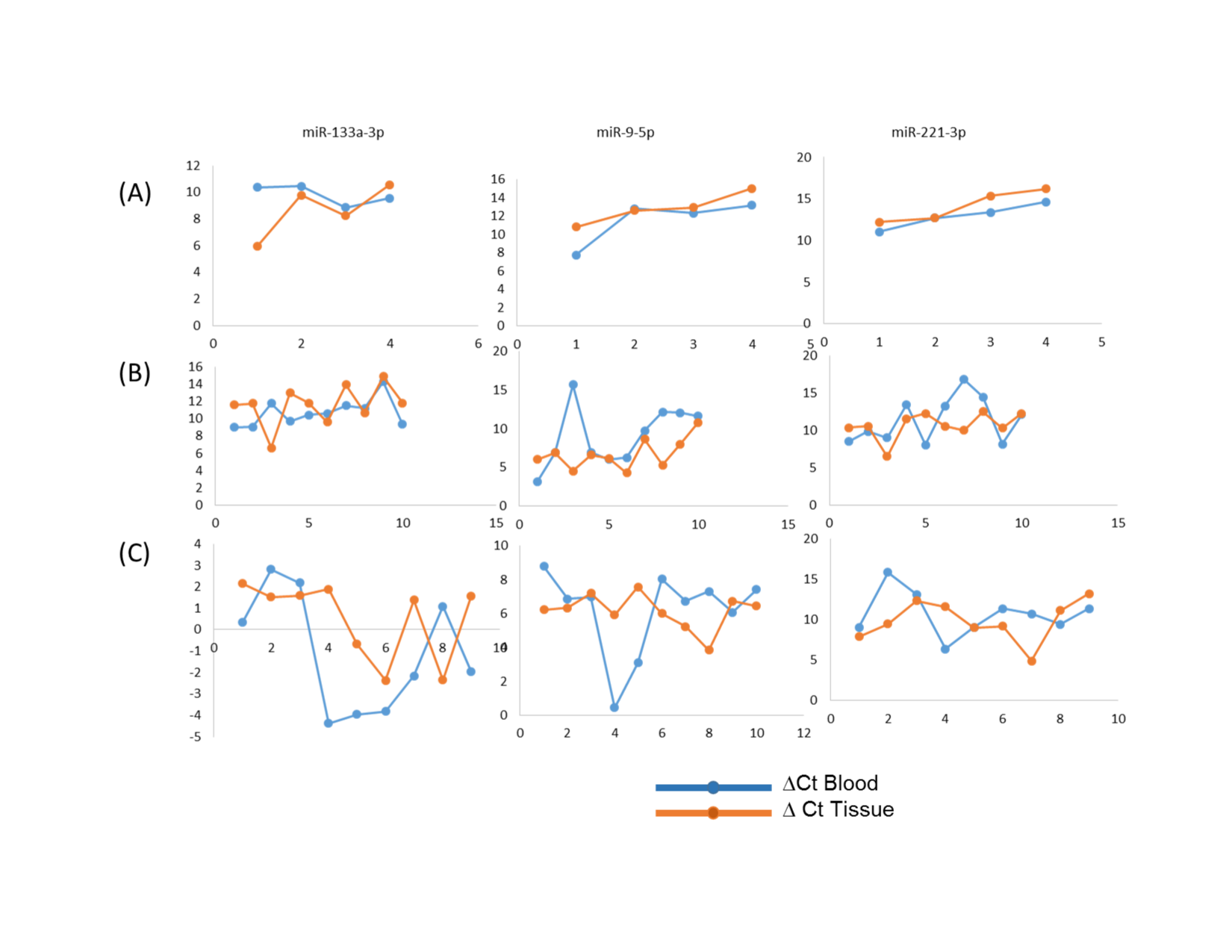
